# Monensin as potential drug for treatment of SLeX-positive tumors

**DOI:** 10.1101/2024.03.11.24304048

**Authors:** Ana F. Costa, Emanuel Senra, Diana Campos, Isabel Faria-Ramos, Liliana Santos-Ferreira, Sofia Lamas, Joana Gomes, Filipe Pinto, Andreia Teixeira, Rafaela Abrantes, Henrique O. Duarte, Mariana Pacheco, Marta T. Pinto, André F. Maia, António Pombinho, Rita Barros, Verónica Fernandes, Frederica Casanova-Gonçalves, Fabiana Sousa, José Barbosa, Luísa Pereira, Fátima Carneiro, Celso A. Reis, Catarina Gomes

## Abstract

Colorectal (CRC) and gastric (GC) cancers remain the top lethal cancers and targeted therapies in this setting are still very limited. Sialyl Lewis X (SLeX), a cancer-associated glycan highly expressed in both CRC and GC, plays a crucial role in cancer cell dissemination and metastasis. Thus, presenting a promising but still underexplored therapeutic target. In this work, we performed a high-throughput screening (HTS) approach to identify potential inhibitors of SLeX expression on cancer cells. Two libraries including a total of 7836 compounds were screened and monensin emerged as a promising SLeX inhibitor. Monensin promoted structural alterations in the secretory pathway, particularly at the Golgi apparatus, impacting protein *O*-glycosylation and secretion. RNAseq transcriptomic analysis uncovered significant alterations in Gene Ontology (GO) terms associated with protein misfolding, target to the membrane, as well as, epithelial cell-cell adhesion protein. *In vitro* studies showed that, upon treatment with monensin, SLeX-positive cancer cells showed reduced viability, concomitant with decreased motility and invasive capacities. Using *in vivo* xenograft models of chick embryo chorioallantoic membrane (CAM) and nude mice, revealed that monensin reduced tumor formation and invasion. Pre-clinical validation using gastric cancer patient-derived organoids (PDOs) and organoid xenotransplants in mice further underscored the clinical potential of monensin in suppressing the growth of SLeX- positive tumors. Overall, our findings set the ground for further evaluation of monensin as a novel therapeutic agent in GC and CRC in the clinical setting.

## Introduction

The limitations of currently available cancer therapies frequently lead to tumor recurrence and metastasis, with the latter being, in fact, the major cause of mortality in cancer patients, accounting for approximately 90% of cancer-related deaths [1]. Gastrointestinal (GI) tract tumors, including gastric (GC) and colorectal (CRC) cancer, are major contributors to the current global landscape of cancer-associated mortality [2]. Therefore, it is of the utmost importance to search and identify novel molecular targets novel anti-cancer agents with superior therapeutic efficacy.

A deeper understanding of the molecular changes that occur in neoplastic cells is key for drug discovery. Alterations in the glycosylation process are well-established features in cancer [3] and have been associated to tumor development, progression, therapy resistance, as well as cancer cell communication [4]. Particularly, increased expression of the Sialyl Lewis X (SLeX; Neu5Acα2-3Galβ1-4[Fucα1-3]GlcNAc-R) antigen is commonly found in epithelial-derived tumors and has been associated with the malignant behavior of cancer cells and with more aggressive tumors [5–7]. Indeed, SLeX expression in numerous cancer types sustains tumor cell invasion and metastization, by facilitating tumor cell motility, adhesion to endothelial cells and interaction with platelets within blood vessels, promoting immune evasion and extravasation to distant sites [6, 8, 9]. Indeed, we have shown that the motility of glycoengineered GC and CRC cells depends on SLeX expression [10]. Additionally, the increased invasive capacity of GC cells, due to SLeX expression, was associated to the oncogenic activation of the c-Met signaling axis [7]. Therefore, SLeX holds promise as a therapeutic target, since its inhibition will lead to reduced tumor growth and metastasis dissemination.

The emergence of high-throughput screening (HTS) methodologies have provided a useful tool in the search for new therapeutic drugs. The application of such approaches on the discovery of glyco-specific drugs has been limited due to the complexity and spatiotemporal heterogeneity associated with the glycosylation process [11]. Nevertheless, recent advancements in methodologies have opened new possibilities [12, 13]. For instance, an HTS assay using reporters to specifically evaluate GalNAcT inhibitors, identified T3lnh-1 as a specific inhibitor of ppGalNACT3. Moreover, a general inhibitor of glycosylation was identified by HTS using glycoengineered cells expressing tailored glycoproteins [14].

In this work, we aimed to develop an HTS approach specifically targeting the SLeX glycan antigen. We identified a lead compound, monensin, a monovalent cation ionophore, as a potent inhibitor of SLeX expression and of its associated malignant properties. Monensin, recognized as a promising repurposing cancer drug, has been garnered attention in previous HTS based studies and it has been extensively explored across various tumor types [15–27].

In this study, we further shown the potential of monensin as an anti-cancer drug was further substantiated through *in vivo* xenografted models (CAM and mice) and the pre-clinical model of GC patients-derived organoids (PODs). Overall, our findings set the ground for the use of monensin as a novel potential therapeutic strategy in the clinical setting.

## Results

### Fluorescent High-Throughput Screening (HTS) for SLeX inhibitors identifies monensin as the lead compound

The SLeX-positive CRC cell line COLO205, known for its 100% positivity for SLeX expression and monolayer growth pattern [10], was used to HTS screening, facilitating fluorescent image acquisition and accurate data analysis. We screened 7836 compounds comprised two distinct libraries, the Prestwick Chemical library® and the DIVERSet® library (Figure 1a, Supplementary Figure 1). As our inhibitory readout relies on glycosylation turnover at the cell membrane, an optimized 72h incubation period was applied. We developed a custom pipeline in CellProfiler^TM^ to quantify the mean fluorescence intensity per cell obtained from the Texas Red (Alexa 594) channel. Given the heterogeneity in SLeX expression within the selected cell line (Supplementary Figure 1), mean intensities within the four fields of view (fov) exhibited considerable standard deviations, resulting in a Z’-factor of 0.539 (Figure 1b). Compounds that induced a significant decrease in fluorescent signal without causing a substantial loss of cell number (indicative of toxicity) were considered lead HIT compounds.

**Figure 1.**
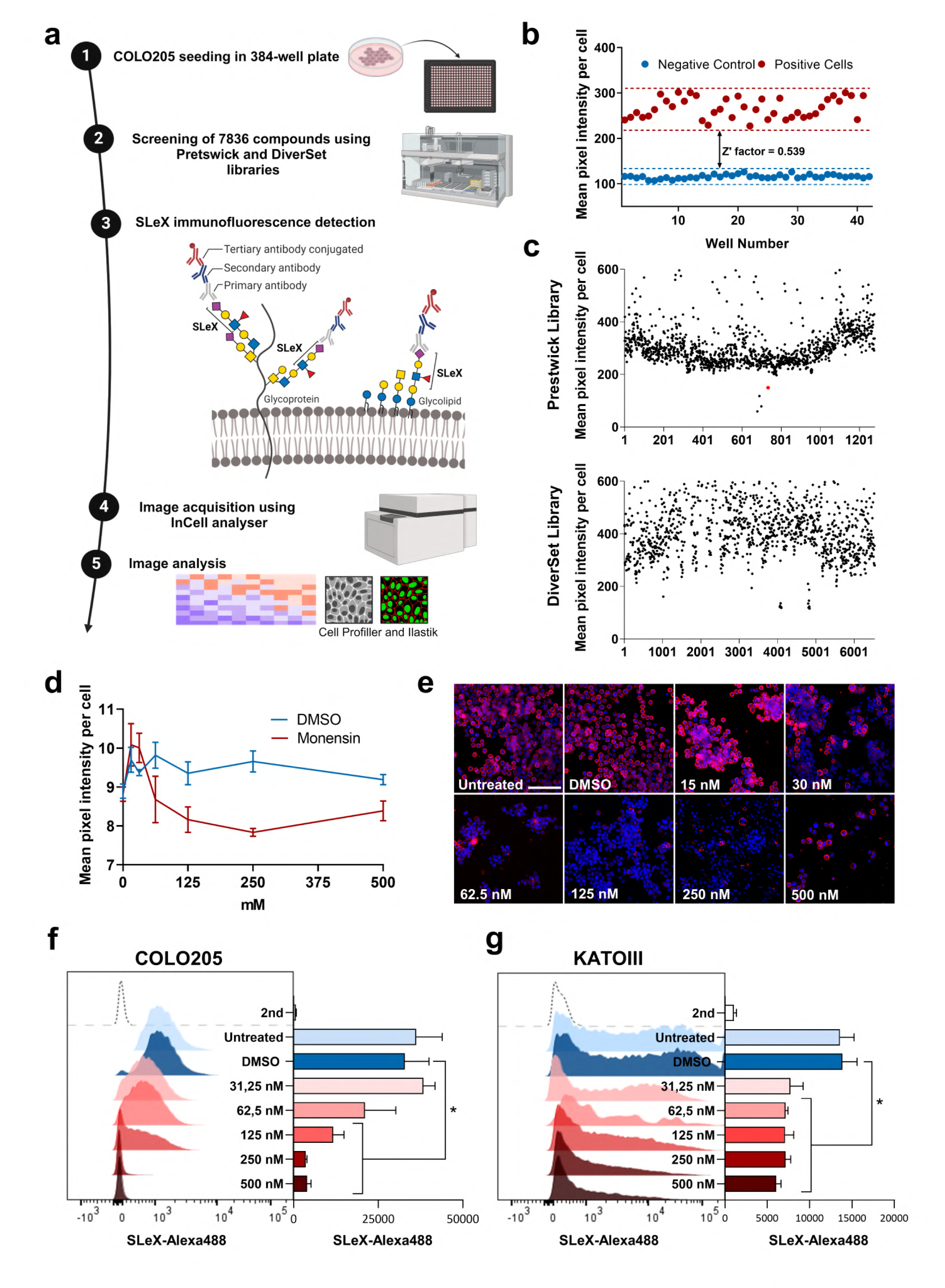
High-throughput screening (HTS) approach identifies monensin as a promising SLeX inhibitor. **(a)** Schematic representation of the adopted methodology involved on the HTS setup. **b)** Z’-factor determination of HTS assay in 384-well plates considering the detection of SLeX in COLO205 WT cells as the positive control and COLO205 ST3GalIV as the negative control. Values of Z’ higher than 0,5 allow us to proceed with confidence with the HTS assay **(c)** Graphic representation of SLeX detection in each well containing the different screened compounds from two distinct libraries (Prestwick® and DIVERSet®). Monensin (red circle) was one of the identified hits, according to the selection criteria **(d)** Graphic representation of SLeX irnmunodetection at the cell surface of COLO205 cells treated with DMSO (blue) or with different monensin concentrations ranging from O to 500 nM (red), and **(e)** representative images of irnmunotluorescence staining showing the reduction in SLeX irnmunodetection in the different tested monensin concentrations in a dose dependent manner. The scale bar corresponds to I00 µm. **(f, g)** SLeX irnmunodetection by flow cytometry in COLO205 cells (f) and KATOIII cells **(g)** upon treatment with DMSO (blue) or monensin (red), validated the inhibitory capacity ofmonensin in a dose-response manner. *p<0.01 vs. the DMSO-treated group.

Most of the tested compounds from our screening assay showed no significant impact on SLeX expression (Figure 1c). Interestingly, while some compounds resulted in increased SLeX levels, a few led to decreased SLeX expression. Manual image analysis revealed that, although some compounds seem to induce a higher SLeX expression level, the observed intensity signal was due to cell debris and, therefore, considered a false result. high throughput screening approach, the Prestwick Chemical library® yielded better results, rendering 4 HIT candidates: glimepiride, auranofin, lanatoside C, and monensin. According to our HIT selection criteria, which considered HITs with less than 200 cells per well as false positives, monensin fulfilled all the defined criteria (Figure 1c) and therefore, was selected for the subsequent validation assays.

### Monensin decreases SLeX expression on the surface of cancer cells

The inhibitory capacity of monensin towards SLeX expression was validated in COLO205 through a dose–response manner, using concentrations ranging from 0 to 500 nM and the same fluorescent detection assay employed in the HTS. The results demonstrated that monensin effectively inhibits SLeX expression at the cell surface, with concentrations of 100 and 200 nM significative reducing SLeX expression without compromising cell viability (Figure 1d and e). To further validate monensin’s inhibitory capacity, we assessed SLeX expression on COLO205 by flow cytometry in a dose-dependent manner. COLO205 cells exhibited decreased SLeX expression at the cell surface when treated with monensin concentrations above 100 nM, highlighting its promising inhibitory effect (Figure 1f). This inhibitory effect was also validated in the KATOIII GC cell line, with flow cytometry results revealing a dose-dependent inhibition of SLeX expression (Figure 1g). Overall, these findings underscore monensin’s potential as a promising drug to inhibit SLeX-associated malignant features of CRC and GC cells.

### Monensin impacts Golgi morphology and function, affecting protein glycosylation and processing

Treatment of CRC and GC cells with monensin lead to marked alterations in the cellular morphology (Supplementary Figure 2a). Monensin-treated cells exhibited larger vacuoles and increased cellular aggregation, indicative of a phenotype associated with cytosolic stress. Transmission electron microscopy (TEM) analysis was employed to investigate potential alterations induced by monensin treatment regarding cytoarchitecture and organelle morphology. TEM analysis of monensin-treated cells revealed the increased presence of autophagic and Golgi-associated vacuoles indicative of Golgi stress in both COLO205 and KATOIII cells (Figure 2a). Additionally, we identified altered mitochondrial morphology in monensin-treated GC cells (Figure 2a).

**Figure 2:**
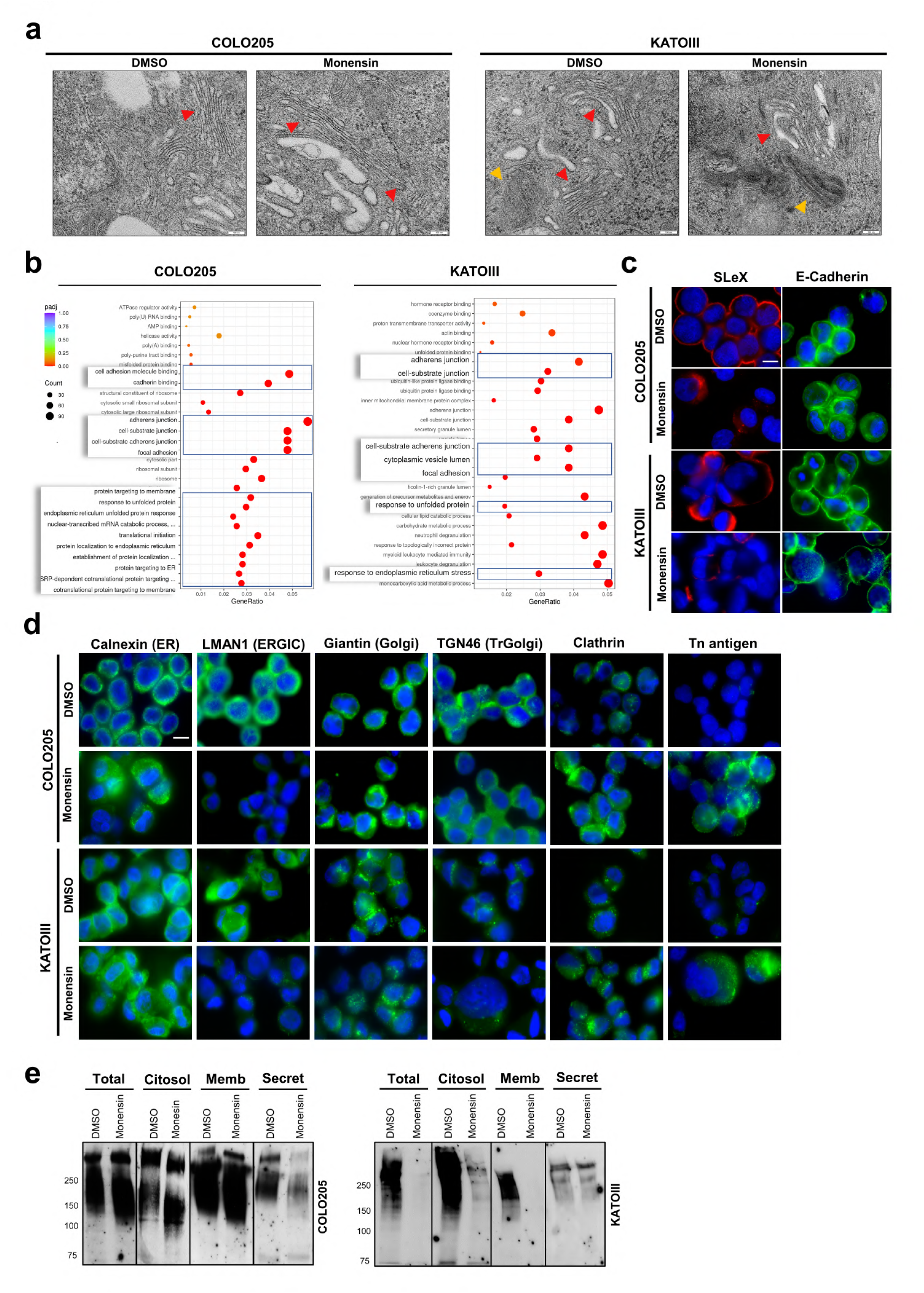
Molecular impact of monensin in COLO205 and KATOIII cell lines. **(a)** Transmission electron microscopy (TEM) analysis, showing morphological alterations mainly at the Golgi apparatus (red arrow) and mitochondria (yellow arrow) upon monensin treatment. Images were acquired at a 12000-20000X magnification. **(b)** Gene ontology enrichment analysis from RNA sequencing (RNA-seq) revealing that monensin treatment led to transcriptomic alterations associated with endoplasmatic reticulum (ER) stress, protein folding and targeting to the cell membrane and adhesion. **(c)** Immunodetection of SLeX (left panel), and E-cadherin (right panel) in COLO205 and KATOIII cell lines upon treatment with DMSO or monensin displaying a reduction and mislocalization in SLeX and E-cadherin expression. **(d)** Immunotluorescence analysis of molecular makers of protein processing at cellular compartments showing alterations in the expression and localization of calnexin (ER), LMAN1 (ERGIC), Giantin (Golgi), TGN46 (TransGolgi), Tn antigen and Clathrin (endocytosis). Images were acquired at 630X magnification, and the scale bar corresponds to 10 µm. (e) Western blot analysis of SLeX expression in different cellular compartments (total, cytosolic, membrane, and secreted) in COLO205 (right) and KATOIII (left) cells treated with DMSO or monensin, demonstrating a reduction in cell membrane and secretion of SLeX-positive proteins.

We further explored the molecular mechanisms associated to monensin treatment in COLO205 and KATOIII cancer cells using RNA sequencing transcriptomics. Gene ontology (GO) enrichment analysis of DMSO- and monensin-treated cells uncovered significant alterations in pathways primarily associated with i) endoplasmic reticulum (ER) unfolded protein response (UPR), ii) response to misfolded proteins, iii) protein targeting to the membrane, and iv) cell adhesion molecule binding (Figure 2b). Indeed, the epithelial cell-cell adhesion protein E-cadherin was shown to have altered levels of expression (Supplementary Figure 2b) and mislocalization to the cytosol (Figure 2c), particularly in COLO205 cancer cells, validating monensin’s impact on the expression of adhesion molecules. Moreover, although some SLeX immunodetection persisted in monensin-treated cancer cells, it was mainly confined to a Golgi-like pattern of expression (Figure 2c), indicative of unfolded protein retention within cells. Considering that protein glycosylation, mainly O-glycosylation, occurs at the Golgi apparatus and is essential for proper protein folding, secretion and function, we explored the impact of monensin on the secretory pathway and SLeX biosynthesis. Our findings revealed differences in the pattern and levels of expression of ER, ER-Golgi intermediate compartment (ERGIC) and Golgi markers, highlighting significant reduction and diffused expression in ERGIC and trans-Golgi (Figure 2d). Additionally, treatment with monensin lead to the increased expression of clathrin endocytosis marker. Moreover, monensin disrupted the proper O-glycan biosynthesis, leading to the expression of Tn glycan, the first glycan structure on the O-glycan synthesis (Figure 2d). This shows that the O-glycan biosynthesis is disrupted in the early elongation steps, hampering the synthesis of more complex structures such as SLeX. As an outcome, we observed a reduction in cell membrane and secretion of SLeX-positive proteins (Figure 2e).

### Monensin undermines SLeX-induced malignant features in CRC and GC cells

Given monensin’s ability to reduce SLeX expression on the cancer cell surface, we comprehensively assess its biological impact and therapeutic potential. We observed that monensin treatment affected the metabolism of SLeX-positive cancer cells COLO205 and KATOIII (Figure 3a). Notably, COLO205 cells showed a statistically significant increase in cellular metabolism at 24h and 48h when subjected to both 100 and 200 nM monensin concentrations. However, this effect was not evident at 72h although higher concentration (500 nM) of monensin negatively impacted cellular metabolism. Conversely, in KATOIII cells, concentrations superior to 100 nM negatively affected cellular metabolism after 48h of treatment. Remarkably, no significant effects were observed in SLeX-negative CRC and GC cells (Figure 3a).

**Figure 3:**
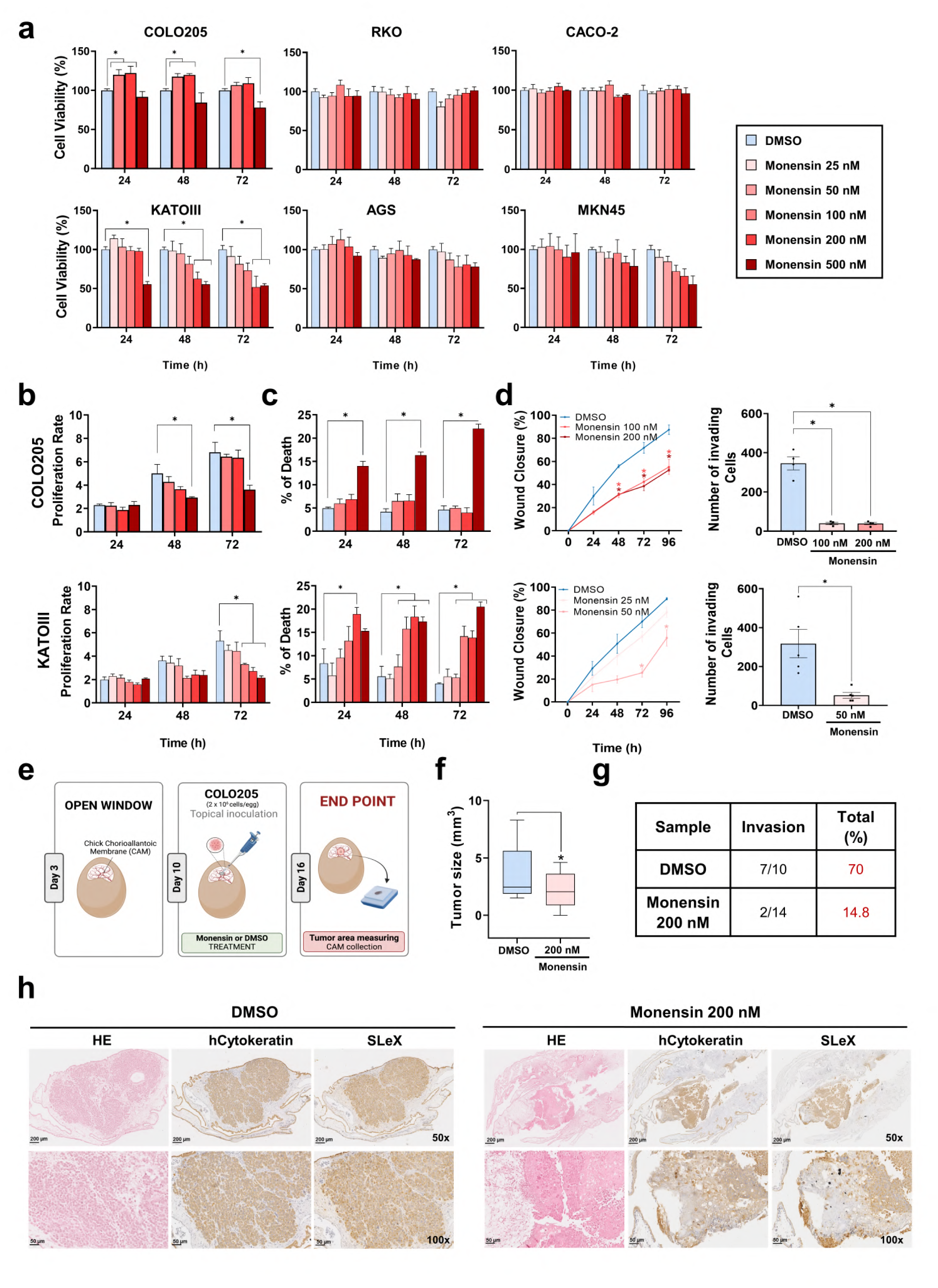
Effect of monensin on cancer cell metabolism, death, proliferation and invasion. **(a)** Evaluation of cell metabolism through the MTT assay in SLeX-positive (COLO205 and KATOIII) and SLeX-negative cancer cell lines (RKO, CACO-2, AGS and MKN45), showing that Monensin can significantly impact cell viability only in SLeX-positive cells **(b)** Proliferation rate of COLO205 and KATOIII cancer cells measured along 72h of treatment, demonstrating monensin’s inhibitory activity at higher concentrations **(c)** Cell death of COLO205 and KATOIII cancer cells treated with monensin for 72h, showing increased cell death mostly in KATOIII cells. **(d)** Wound healing (Left) and invasion (Right) assays, showing monensin’s ability to reduce COLO205 and KATOill migratory and invasive properties. **(e)** Schematic representation of the workflow adopted for the *in vivo* study of monensin impact on tumor formation and invasion using the chick chorioallantoic membrane (CAM) model. **(f)** Measurement of COLO205 derived xenografted tumor area, showing the development of smaller tumors by monensin-treated cancer cells (Righ) when compared to DMSO (Left). All results are presented as mean ± SEM of at least three independent experiments. *p<0.01 vs. the DMSO-treated group. **(g)** Number of invading xenografted COLO205 derived tumors, showing monensin’s ability to efficiently reduce tumor cell invasion when compared to DMSO-treated cells. **(h)** Representative images of immunohistochemical analysis of CAMs, including Hematoxilin & Eosin (H&E) staining, and human cytokeratin and SLeX immunodetection. Cytokeratin immunostaining dispicting the reduced invasive capacity of Monensin-treated cells in the chicken membrane, concomitant with reduced SLeX expression.

Subsequently, we investigated whether the observed monensin-induced metabolic alterations resulted in variations in cell proliferation and cell death. We observed that in COLO205 cells, concentrations ranging from 25 to 200 nM did not have an impact on cell proliferation and cell death (Figure 3b and c). An exception was noted at 500 nM concentration, which induced a substantial reduction in proliferation and an increase in cell death, indicative of cellular toxicity. Regarding KATOIII cells, concentrations higher than 50 nM resulted in a significant reduction in proliferation rates at 72h (Figure 3b). Similarly, concentrations above 50 nM led to a significant increase in cell death after 24h or 48h, in particular with the 500 nM concentration (Figure 3c), suggesting its potential as a killing agent.

Given the known association of SLeX expression with cancer cell invasion and the presence of metastasis in CRC and GC [6, 9], we assessed monensin’s ability to impair cell motility and invasion. To this end, COLO205 and KATOIII cancer cells were treated with monensin for 72h and a wound healing and Matrigel-chamber invasion assay were performed. Monensin concentrations that showed significant differences in cell proliferation and cell death were not considered in these assays. Our results revealed that both COLO205 and KATOIII cells treated with monensin displayed a decreased capacity to migrate and invade when compared to DMSO-treated cells (Figure 3d). These results, underscore the potential of monensin to impair SLeX-associated motility and invasive capacities *in vitro*.

To further elucidate the *in vivo* effect of monensin, we employed the chick chorioallantoic membrane (CAM) model to evaluate the growth and metastatic potential of both KATOIII and COLO205 cells. KATOIII cells cells present a diffuse growth pattern, thus an accurate measure of the CAM xenographed size is not possible (Supplementary Figure 3a). Given that only COLO205 cells formed measurable tumors, this was the cell model analysed. Tumor formation was measured 6 days post-inoculation of the cells on the top of the CAMs (Figure 3e). Results showed that monensin-treated cells formed significantly smaller tumors (Figure 3f). In addition, DMSO-treated cells exhibited high invasive capacity (70%), significantly higher that monensin-treated cell (14,8%). as evidenced by H&E staining and IHCs stainings monensin-treated cells also showed reduced SLeX expression (Figure 3g and h and Supplementary Figure 3b). Collectively, these results underscore the ability of monensin to reduce SLeX expression on cancer cells and mitigate their associated malignant properties *in vivo*.

### Monensin impacts tumor growth in mice

To further substantiate the *in vivo* potential of monensin in inhibiting SLeX-driven malignancy, we then assessed monensin’s therapeutic efficacy in a COLO205 xenograft mouse. 1 and 5×10^6^ COLO205 cells were subcutaneously inoculated into nude mice, and mice were intraperitonially (IP) treated after day 7 with monensin (10 mg/kg) or PBS. Tumor burden and SLeX expression were then analyzed (Figure 4a). The results show that monensin significantly inhibited tumor growth, irrespective of cellular inoculum (Figure 4b and c, Supplementary Figure 4a). In addition, SLeX immunodetection in collected monensin-treated tumors was significantly decreased (Figure 4d and Supplementary Figure 4b and c). Notably, COLO205 xenografts exhibited rapid tumor growth, leading to ulceration within a few days (around day 10 after starting treatment). Quantification of tumor ulceration revealed positive outcomes in monensin-treated mice comparing to untreated tumors, that exhibited a higher percentage of ulceration (Figure 4e). Tissue analysis of collected organs did not show any metastatic lesion in any group of mice (Supplementary Figure 4d), which can be attributed to the relatively short duration of this experiment. Taken altogether, monensin is shown to effectively control tumor growth *in vivo*.

**Figure 4:**
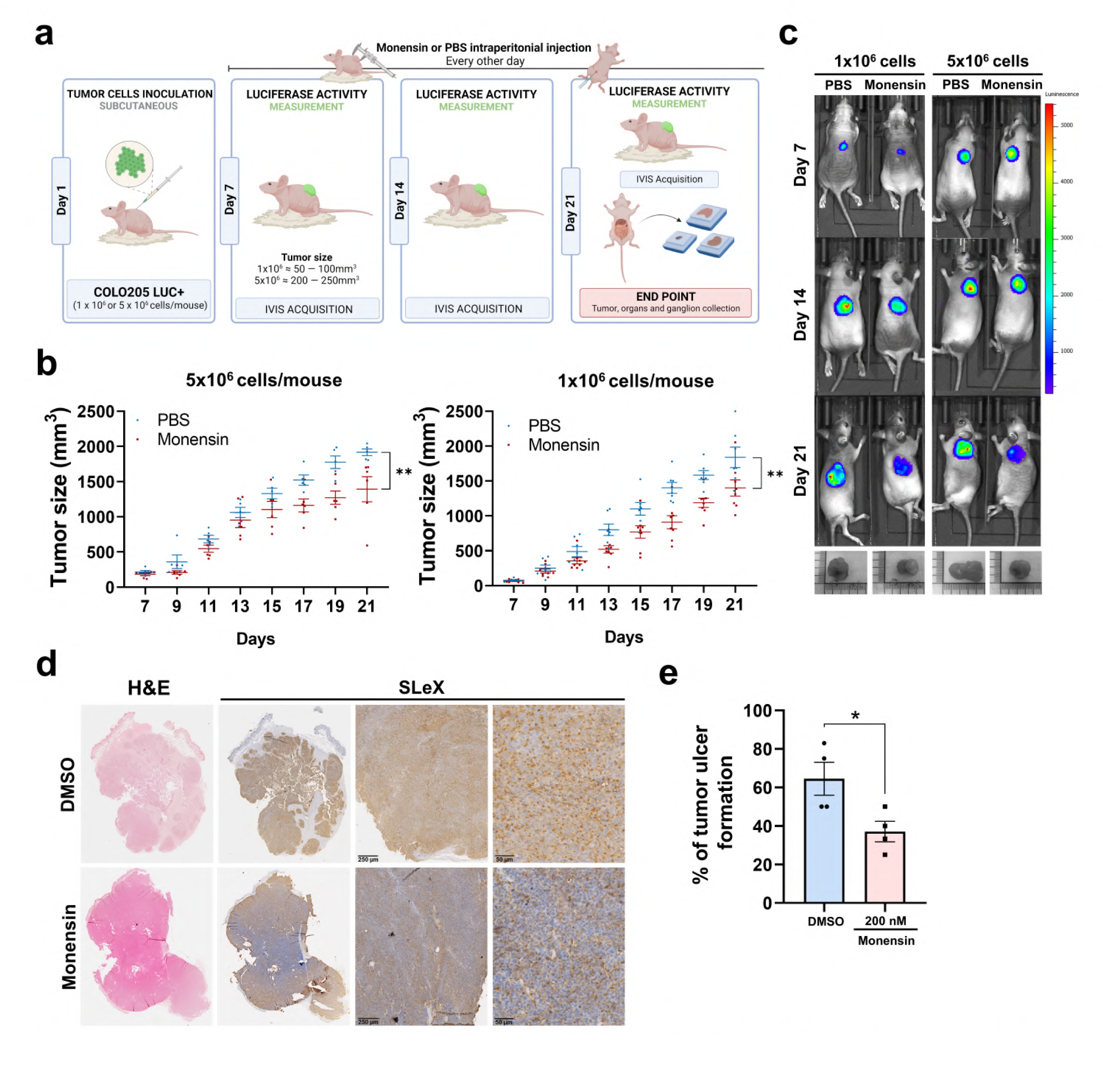
*In vivo* biological effect of monensin using a xenograft mouse model. **(a)** Schematic representation of the tnice experiment. **(b)** Tumor growth rate along time of the experiment showing the reduced tumor burden in monensin treated mice. **(c)** Representative images of *in vivo* tumor growth by measuring luciferase activity throughout time, showing the reduced tumor growth in monensin-treated mice. **(d)** Tumor images of H&E staining and immunohistochemistry of SLeX expression in representative tnice, demonstrating a reduction in SLeX immunostaining in tumors from monensin­ treated mice. **(e)** Quantification of tumor ulceration in mice, showing increased number of ulcerative tumors in PBS­ treated mice. Results are presented as mean ± SEM. *P<0.0 I vs. DMSO-treated group.

### Monensin inhibits tumor cell proliferation in SLeX-expressing patient-derived organoids

Patient-derived organoids (PDOs) are recognized as important models to study molecular pathways underlying carcinogenesis and therapeutic resistance, but also for conducting drug screening experiments [28–30]. We assessed monensin’s therapeutic efficacy in four distinct GC-derived SLeX-positive PDOs representing papillary/ intestinal, poorly cohesive and mixed carcinoma subtypes, as well as in two organoids derived from normal mucosa adjacent to tumor (models established in the group, unpublished data) (Figure 5a). Monensin’s IC50 in PDOs was determined in a dose-response manner, using concentrations ranging from 0 to 400 nM. The results demonstrated a heterogeneous *in vitro* response to monensin, with normal derived organoids presenting an IC 50 of 140 nM, and GC-derived PDOs an IC 50 of 50 nM or resistant (Fig 5b and Supplementary Figure 5a). Additionally, immunofluorescence analysis revealed the capacity of monensin to impair SLeX biosynthesis, as shown by its reduced expression at cell surface of treated PDOs (Figure 5c).

**Figure 5:**
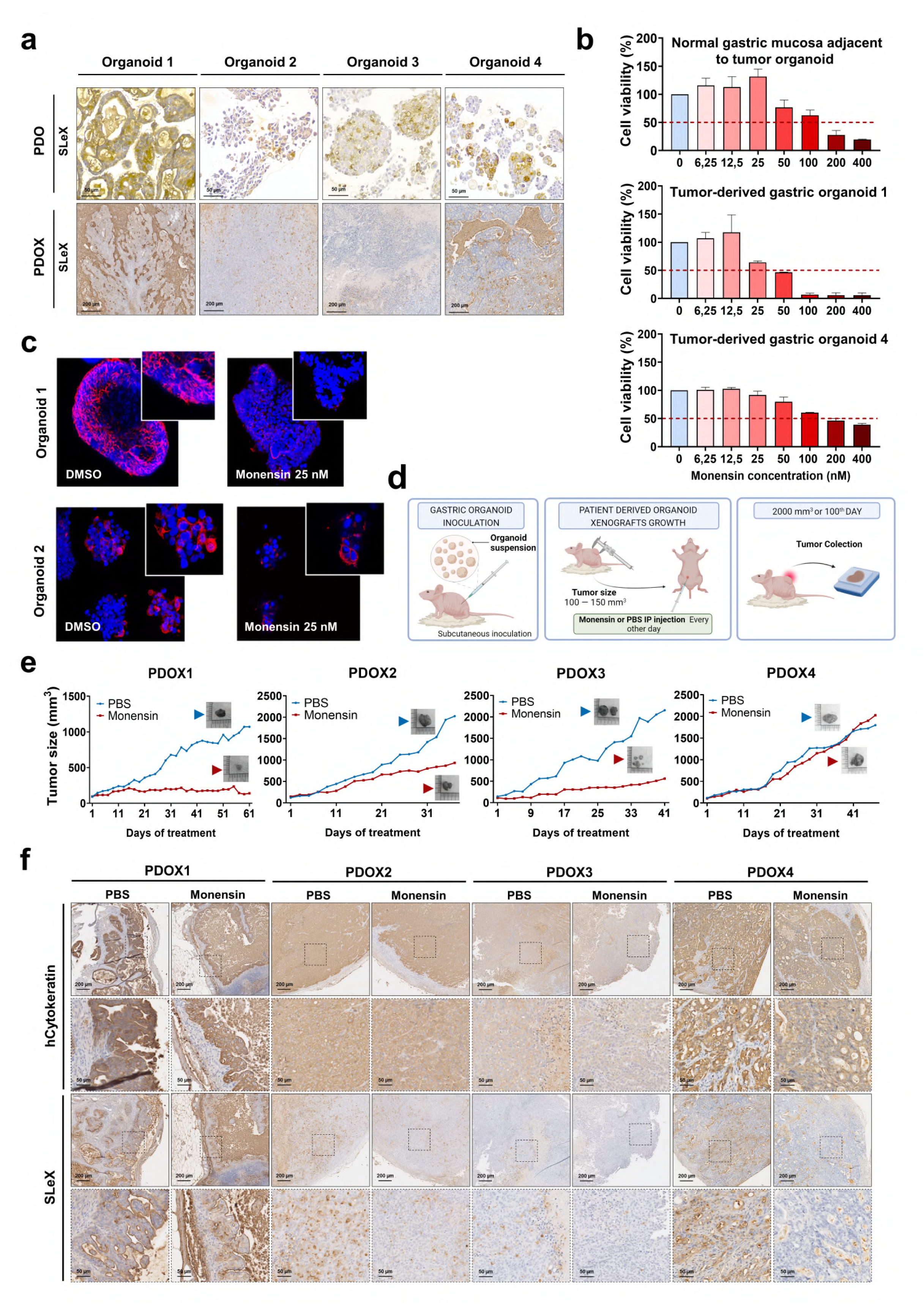
Monensin biological effect in a pre-clinical organoid model. **(a)** Histology (H&E) and SLeX expression of GC tumor patient-derived organoids (PDOs) and its respective patient-derived organoid xenograft (PDOX) tumor tissue collected from mice. **b)** Viability assay of normal mucosa adjacent to tumor (top) and two tumor patient (botton) -derived gastric organoids after monensin treatment showing differences in sensitivity in the different models. **(c)** Immunofluorescence analysis of SLeX expression in SLeX-positive PDOs after monensin treatment showing the reduction of SLeX. 400 and 630x magnification. **(d)** Schematic representation of the experimental design followed for the establishment of PDOX mouse model. **(e)** Tumor growth rate of GC PDOs in mice showing the reduction of tumor size upon treatment with monensin in comparison to PBS-treated mice. **(f)** Histological characterization of GC PDOXs tumors, showing a preserved tumor histoarchitecture (human cytokeratin staining) and a reduction of SLeX immunostaining in tumors from monensin-treated mice.

To further validate monensin therapeutic activity under physiological conditions, we used a Patient-Derived Organoid Xenograft (PDOX) mice model. PDOs were subcutaneously implanted in mice dorsal area, and when tumors reached approximately 100mm^3^ in size, were treated with monensin via IP administration (Figure 5d). Importantly, PDOX models preserved the histoarchitecture and SLeX expression pattern of PDOs (Figure 5a). Notably, none of the normal mucosa adjacent to tumor-derived organoids successfully developed in mice, limiting monensin testing to GC-PDOXs. Results showed a significant impact of monensin on tumor growth in 3 out of the 4 tested PDOXs, with a more pronounced effect observed in organoids expressing high levels of SLeX compared to those with moderate expression (Figure 5e). In the case of PDOX4, although no statistically significant difference in tumor size over time was observed, gross histological examination revealed a substantial necrotic area within monensin-treated PDOX4 in relation to PBS control PDOX (Supplementary Figure 5b). Neither monensin-treated mice nor control mice displayed signs of physiological or behavioral distress, demonstrating no significant side effects of monensin and therefore human endpoints were not considered along the experiment.

Histological examination of the tumors collected from the PDOX mice revealed a reduction in papillary-like structures in monensin-treated cases derived from papillary carcinomas (Figure 5f and Supplementary Figure 5b and c). Additionally, the observed tumor bed of monensin-treated PDOXs is rich in necrotic areas and infiltrated immune cells, characteristic of tissue death.

Furthermore, an impact on the protein secretory pathway was evident, with tumor cells exhibiting a cytoplasmic diffusely stained for SLeX in monensin-treated mice, in contrast to the clear membrane staining in PBS-treated mice (Figure5f).

## Discussion

CRC and GC rank the top deadliest cancers, primarily due to their diagnosis at advanced stages where metastasis already occurred [2], emphasizing the urgent need for new and effective treatment options [31]. SLeX plays an undeniably central role within the molecular cascade underlying the metastization process of both CRC and GC [as reviewed in 9]. We designed an HTS-based assay to identify inhibitors of the biosynthetic pathway of this aberrantly expressed glycan that could serve as therapeutic agents against CRC and GC. Remarkably, we successfully pinpointed monensin as the most promising inhibitor of SLeX in both CRC and CG cells.

Monensin, a polyether ionophore, is a fermentation product derived from *Streptomyces cinnamonensis*. Initially discovered in 1967 by Agtarap *et al.* [32], monensin finds widespread use in veterinary practice for addressing bacterial, fungal, and parasitic infections, particularly in poultry and cattle production.

In this study, we identified the capacity of monensin to inhibit SLeX, through the disruption of the well-coordinated glycosylation process, with a concomitant increased expression of the truncated *O*-glycan Tn antigen. We mainly attribute these glycan changes to the observed morphological and functional alterations in the Golgi apparatus structure, as shown by TEM analysis and the use of ERGIC and Golgi markers, crucial for protein *O*-glycosylation and terminal *N*-glycosylation. In fact, previous studies have highlighted the impact of monensin on glycosylation [33]. These studies reported undersulfation of glycosaminoglycans and alterations in both *N*- and *O*-linked glycans. Nonetheless, there are very few reports describing a direct link between monensin and altered glycan biosynthesis, with the main evidence focusing on alterations to the Golgi morphology and function [34, 35].

Given its characteristics as ionophore, monensin deregulates cation influxes, specifically Na^+^ and Ca^2+^, within the Golgi compartment. This disruption promotes, as observed by TEM analysis, a noticeable swollen phenotype, compromising Golgi’s function and consequently impacting protein glycosylation, processing, and secretion [35]. In fact, the present work revealed an altered expression in genes related to ER stress, protein folding and secretion, and adhesion processes, as shown by transcriptomic analysis. Our findings showed that there was an impairment of SLeX protein carriers at the cell membrane, as well as the secreted forms, alongside with the abnormal expression of E-cadherin in treated cells. Indeed, monensin has been used as a standard reagent in the inhibition of protein transport [24] and cytokine secretion [36, 37], corroborating the observed effects. Additionally, monensin has been described as an inducer of oxidative stress, hindering cell growth by inducing G1-phase cell cycle arrest, ultimately resulting in cancer cell apoptosis [18, 22]. The oxidative stress generated has been reported to alter mitochondrial function [15, 27], a phenomenon also observed in GC cells used in our study.

In this study, we have also observed an increased *in vitro* sensitivity of SLeX-positive cancer cells to monensin. Specifically, we showed that from six GC and CRC cell lines tested, monensin impacted only the two SLeX positive cell lines, potentially narrowing down its effect to this particular glycan structure. Monensin forms stable complexes with lipid-soluble cations, enabling rapid traversal trough cell membranes. Building on this, we hypothesize that the anionic charge conferred by sialic acids at cell surface of SLeX-positive cancer cells [38] potentially facilitates monensin’s uptake and intracellular transport, rendering SLeX-positive cancer cells more sensitive to treatment. This hypothesis aligns not only with the differences in the metabolic responses of monensin-treated SLeX-positive and SLeX-negative cells, but also with an increased immunodetection of clathrin, a fundamental protein in intracellular vesicle formation responsible for extracellular material uptake into cells [39]. Consistent with this evidence, Vanneste *et al.* conducted a screening of 23 human cancer cell lines to assess their sensitivity to monensin. Their findings revealed 12 cell lines responsive and 11 cell lines resistant to monensin, from which HT-29 and HCT-116 exhibit sensitivity and resistance to monensin, respectively [22]. Notably, HT-29 is a SLeX-positive cell line, whereas HCT-116 is SLeX-negative cell [10], underscoring the potential of SLeX in influencing the sensitivity of cancer cells to monensin treatment.

SLeX positive tumors are inherently associated with more aggressive and invasive cancer behaviors [6, 7, 10]. In the present study, we pointed out monensin’s efficacy in inhibiting SLeX expression, suppressing cancer cell motility and invasion in cell-based assays, and leading to a sustained reduction in tumor growth across various *in vivo* models, without inducing any toxic effects in mice model. This aligns with a recent report highlighting monensin’s specificity in targeting hepatocellular carcinoma allografts without adversely affecting normal hepatic and non-hepatic tissues. This selectivity is attributed to the energetic incapacity of hepatocellular carcinoma cells to compensate and survive the Na^+^ load induced by monensin [40].

Drug repurposing strategies drugs emerge as cost-effective and promising avenue for discovering novel small molecule-based therapies [41]. Despite the inherent challenges and limitations associated with drug repurposing, several anticancer drugs repurposed from existing medications are currently advancing through clinical trials, with some already obtaining approval [42]. As for monensin, despite the studies that supports its use as a promising drug for cancer treatment, no clinical trials have been conducted, and concerns regarding its toxicity in humans still persist. Toxicity reports typically involve the ingestion of uncontrolled dosages of monensin formulas used for the poultry or cattle industry [23, 43–46]. Thus, it is important the control of monensin dosage, formulation, and administration protocols to be considered its clinical use.

Overall, our study contributes to the evolving landscape of cancer drug discovery by demonstrating the remarkable capacity of monensin to target SLeX-positive CRC and GC cells. Beyond its impact on glycosylation pathways and SLeX expression, monensin exerts an influence on associated malignant properties, notably impeding tumor invasion and growth. This not only underscores the versatility of monensin but also position it as a potential therapeutic candidate for cancer treatment.

### Conclusions

The identification of novel and effective cancer treatment options is paramount in cancer research. Nevertheless, progress is hindered by the currently available approaches that often yield ineffective drugs in clinical trials. Recent studies have explored the potential of repurposing existing drugs, demonstrating its huge potential for the development of anticancer drugs [47].

This work acknowledges the importance of targeting cancer-specific glycans as a new source of cancer drugs, and further addresses how SLeX inhibition by monensin can impair the malignant behavior of cancer cells. Overall, this study highlights the significant potential of monensin as an anticancer agent for CRC and GC.

## Materials and Methods

### Cell Culture and Cell lines

AGS, CACO-2, COLO205, KATOIII and RKO were obtained from ATCC (American Type Culture Collection) and MKN45 was obtained from the Japanese Cancer Research Resources Bank. Cells were grown in monolayer culture and maintained at 37 °C in an atmosphere of 5% CO2, in Roswell Park Memorial Institute (RPMI) 1640 GlutaMAX, HEPES medium, Dulbecco’s Modified Eagle Medium (DMEM) or Iscove Modified Dulbecco Media (IMDM), all supplemented with 10% fetal bovine serum (FBS) (all from Biowest).

### High-throughput screening of chemical compound’s libraries

COLO205 cells were seeded in 384-well plates (CellCarrier-384 Ultra, PerkinElmer) at a density of 1 × 10^5^ cells/mL in RPMI medium supplemented with 10% FBS and allowed to adhere overnight. Two distinct libraries totalizing 7,836 chemical compounds (1,248 compounds from the Prestwick Chemical Library®, and 6,588 from DIVERSet® library) were then transferred, one compound per well, using a pintool (V&P Scientific) coupled to a MDT head of a JANUS Automated Workstation (PerkinElmer) for a final concentration of 4 µM in DMSO. Cells were incubated for 72 h, after which they were fixed with formaldehyde to a final concentration of 4% (Sigma Aldrich). SLeX was detected by immunofluorescence. Briefly, cells were incubated with CSLEX mouse antibody (BD Bioscience) overnight at 1:250 dilution, and fluorescent detection was performed using a sequential secondary amplification method of goat anti-mouse IgM and rabbit anti-goat Alexa 594 antibodies (Jackson ImmunoResearch), with 1 h incubation each. Nuclei were stained with DAPI (Sigma-Aldrich). The image acquisition was performed in an INCell Analyzer 2000 (GE Healthcare) with a Nikon 20x/0.45 NA Plan Fluor objective. Four fields of view (fov) were acquired per well. Image analysis was performed using the Ilastik and CellProfiler TM softwares[48, 49]. The image analysis workflow consists in using the machine learning capabilities of Ilastik to accurately segment the nuclei. After, a custom-made pipeline in CellProfiler was devised to quantify the fluorescence intensity per cell obtained from the Texas Red (Alexa 594) channel. Briefly, upon correction of the uneven illumination, the probability maps obtained from the Ilastik software are used to identify the nuclei followed by a 3 pixels expansion of the edges of these objects to create a larger area, intending to reach the cells cytoplasm. Then, the mean pixel intensity of each fov in the Texas Red channel was calculated from the mean pixel intensity values of all identified cells on the image. The pipeline also generates outlines of the identified cells that were used to perform the quality control of the image analysis. To classify a compound as a hit we looked for a significative reduction or absence of Texas Red fluorescence, taking in consideration the number of cells per well to avoid false positives.

The Z’-factor of the assay was calculated according to the mean (µ) and standard deviations (σ) of samples (s) and controls (c), following the equations:

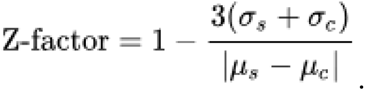

Samples comprise COLO205 WT cells, whereas controls comprise COLO205 ΔST3GalIV cells already described as SLeX negative [10].

### Monensin dose response analysis

Hit confirmation was performed by testing the compounds in a dose-response assay (0nM – 500 nM) in triplicate. For monensin validation, cells were seeded in 96-well plates (CellCarrier-96 Ultra, PerkinElmer) and tested again in dose–response (repurchased compound, Sigma Aldrich) at a density of 3×10^4^ cells/mL and allowed to grow for 24 h at 37°C and 5% CO_2_ in a humidified incubator. Then, cells were incubated with monensin for 72 h and then fixed, stained, and analyzed, as previously described for the primary screen. DMSO was used as a negative control.

### Flow Cytometry analysis

COLO205 and KATOIII cells were seeded in 6-well plates 24 h before treatment with increasing monensin concentrations for 72 h. On the day of analysis, cells were detached using trypsin (ThermoFisher), spin down at 300 g for 5 min at 4 °C. Afterwards, they were stained with CSLEX antibody (dilution 1:250, BDBioscience) for 30 min on ice. Cells were washed and incubated with secondary antibody Alexa 488 anti-mouse IgM (dilution 1:1000, Jackson ImmunoResearch) for 30 min on ice in the dark. Cells were stained with DAPI (10 μg/ml, SigmaAdrich) for 10 min at room temperature in the dark. Samples were analyzed on a FACS Canto II Flow cytometer (BDBioscience) and *FlowJo* software of analysis.

### MTT cell viability analysis

Cell proliferation/viability was analyzed *in vitro* by the tetrazolium salt 3-(4,5-dimethylthiazol-2-yl)-2,5-diphe-nyltetrazolium bromide (MTT) method using a commercially available kit (MTT Cell Proliferation Kit I, Roche). Cells were seeded into 96-well plates at the density of 5×10^3^ cells/well, treated with monensin (or DMSO as control) and MTT Kit solutions added into each well according to cell proliferation kit instructions (Roche) for each time point 24, 48 and 72h. The absorbance value was read at 600 nm using a microplate reader. Three independent assays were performed in triplicates and results are presented as means ± SEM for each sample. The viability levels obtained were normalized and compared with the DMSO treatment control.

In addition, cell proliferation/viability was also assessed by seeding cells in 24-well plate at the density of 2×10^4^ in the late afternoon and were allowed to adhere overnight. In the next morning, cells were treated with monensin (or DMSO as control) and live and death cells were counted after staining with Trypan Blue (ThermoFisher) for three consecutive days (24h, 48 and 72h after treatment). The proliferation rate was calculated using a ratio between the cell number at the specific time after the treatment and the initial cell count.

### Wound Healing Assay

Cell mobility was assessed by a wound healing assay *in vitro*. Approximately 5×10^5^ cells were seeded into 24-well plates until reaching confluency. An incision was made in the central area with a 200-μl pipette tip. After being washed twice with serum-free medium, the cells were then allowed to migrate into the cell-free area. The cells were imaged at 0, 24, 48 and 72 h at a magnification of 100x. Cell migration was calculated as the mean percentage of the cell migrated distance compared with the initial wound distance using ImageJ software (FIJI). The experiment was performed in triplicate in three biological replicates and motility capacity compared within the different conditions.

### Invasion Assay

Invasion assays were performed in a BD Biocoat Matrigel invasion chamber (Corning) with an 8-mm diameter pore size membrane with a thin layer of Matrigel, in a 24-well plate. Inserts were rehydrated for at least 1 h with culture medium. After detachment of confluent cells with trypsin/EDTA, 9×10^4^ cells were seeded in the upper surface of Transwell plates and cultured in: i) culture medium containing 10% FBS (control cells), ii) culture medium containing 10% FBS and DMSO (control treatment cells, final concentration of 0,002%), iii) culture medium containing 10% FBS and 100 nM and 200 nM of monensin for COLO205 cells and 50nM for KATOIII cells (treatment cells), all during 72h in a humidified incubator (37°C, 5% CO2). Complete culture medium (media with 10% FBS) was added in the lower part of the insert. After incubation, non-invading cells in the upper part of the insert were carefully removed with a cotton swab, invasion chambers washed with PBS and lower membrane penetrated cells fixed with methanol. Membranes were removed from the inserts and mounted in a slide using Vectashield with DAPI (Vector labs). Two independent assays were performed, and cells were seeded in duplicate for each condition. Invading cells were counted under a fluorescence microscope (Carl Zeiss), and measurement was done by counting cells, using ImageJ software, from six random fields using a microscope at a magnification of 200x. Results are presented as means ± SEM for each sample, and invasion levels obtained were compared within the different conditions.

### Transmission electron microscopy analysis

For the ultrastructure analysis, cells were fixed in a solution of 2.5% glutaraldehyde (#16316; Electron Microscopy sciences) with 2% formaldehyde (#15713; Electron Microscopy sciences) in 0.1 M sodium cacodylate buffer (pH 7.4) for 2hrs, at RT, and post fixed during 1hr in 1% osmium tetroxide (#19190; Electron Microscopy Sciences) diluted in 0.1 M sodium cacodylate buffer. After centrifugation, the pellet was resuspended in HistogelTM (ThermoFisher Scientific, HG-4000-012) and then stained with aqueous 1% uranyl acetated solution overnight, dehydrated and embedded in Embed-812 resin (#14120; Electron Microscopy sciences). Ultra-thin sections (50 nm thickness) were cut on an RMC Ultramicrotome (PowerTome) using Diatome diamond knifes, mounted on mesh copper grids (Electron Microscopy Sciences), and stained with uranyl acetate substitute (#11000; Electron Microscopy Sciences) and lead citrate (#11300; Electron Microscopy Sciences) for 5 min each. Samples were viewed on a JEOL JEM 1400 transmission electron microscope (JEOL), and images were digitally recorded using a CCD digital camera Orius 1100W. The transmission electronic microscopy was performed at the HEMS core facility at i3S, University of Porto, Portugal with the assistance of Ana Rita Malheiro, Sofia Pacheco and Rui Fernandes.

### RNASeq analysis

Cells were seeded in 6-well plates and allow to seed for 24 h. Then, cells were treated with Monensin or DMSO (vehicle) for 72 h. Cells were harvest and RNA extracted using Pure LinkTM RNA Mini Kit (Invitrogen) according to manufacturer protocol. The quality of the RNA was firstly evaluated by NanoDropTM One Spectrophotometer (ThermoFisher Scientific) and the samples were stored at -20°C until use. Three biological replicates from each condition were guaranteed. The RNA was sent Novogene Co., Ltd. for cDNA library construction, sequencing and bioinformatic analysis. The quality of the paired-end sequencing reads was checked using FastQC and the low-quality reads and adapters were removed using Trimmomatic. The pre-processed reads were mapped to the reference using HISAT2. The resulting read alignments were then used to generate the gene counts using the feature Counts software and normalised to FPKM (Fragments Per Kilobase of transcript sequence per Millions base pairs sequenced). Differential gene expression (DEG) analysis of the gene counts data was performed using DESeq2 R using a differential gene screening threshold of |log2(FoldChange)| >= 1 & padj<= 0.05. Through the enrichment analysis of the differential expressed genes, the enrichment analysis on gene sets (GO) was obtain using the clusterProfiler software.

### Immunofluorescence analysis

Cells were seeded in 13 mm coverslips (Marienfeld) and let adhere overnigth. Then, the cells were treated with Monensin or DMSO for 72 h. The cells were washed with PBS and fixed with 4% Paraformaldehyde fixative solution (Alfa Aesar) during 10 minutes at room temperature. Blocking of immunoglobulins cross-reaction was performed using goat serum in 10% BSA in PBS at room temperature for 30 minutes, followed by the primary antibody incubation at 4°C overnight. The primary antibodies used were: SLeX (CSLEX, 1:100, BD Bioscience,), Tn (1E3, 1:2, Hybridoma), E-Cadherin (4A2C7, 1:200, Invitrogen), Calnexin (C5C9, 1:100, Cell Signaling), ERGIC (LMAN1, 1:100, Cusabio), Giantin (ab24586, 1:1000, Abcam) and TGN46 (ab50595, 1:300, Abcam). Afterwards, incubation with fluorescently labelled secondary antibodies anti-mouse IgM Alexa Fluor® 594 (1:500) or IgG Alexa Fluor® 488 (1:500), or anti-rabbit IgG Alexa Fluor® 488 (1:500, ThermoFisher Scientific) were performed during 1 hour at room temperature and nuclear counter staining performed using 4’,6’-diamino-2fenil-indol (DAPI) for 10 minutes. Washes were performed with PBS. Fluorescent signal was examined using a fluorescence microscope and images were acquired using a Zeiss Axio Carl cam MRm and the AxioVisionRel (version 4.8) analysis software.

### Protein extraction of cellular fractions for downstream analysis

For extraction of total, cytosol, and membrane proteins, COLO205 and KATOIII were cultivated in 15-cm dish and let adhere overnight, and treated with Monensin or DMSO for 72 h. The commercially available kit Pierce™ Cell Surface Biotinylation and Isolation Kit (ThermoFisher) that allow for selective biotinylation, solubilization, and enrichment of plasma membrane proteins was used. For the total protein, after cells lysis step, a small amount of protein was collected. For the cytosol protein, during the isolation labeled proteins step, the flowthrough was collected as it corresponds to non-surface and non-biotinylated proteins. For the enriched membrane protein lysates, in the elution step, a 0,1% Rapigest and 10mN DTT in 50 mM ammonium bicarbonate buffer was used.

For secreted proteins enrichment, COLO205 and KATOIII were cultivated in T75 and let adhere overnight. The cells were washed five times in PBS 1X and treated with Monensin or DMSO, diluted in serum-free culture medium for 72 h. The conditioned media (secretome) was collected and centrifuged for 5 min. at 1200 rpm to remove cell debris and concentrated using 10 kDa Amicon centrifugal filter units (Merck Millipore, Burlington, MA, USA). 4 μL of complete TM protease inhibitor cocktail (Roche) was added per 100 μL of concentrated samples and then stored at −80 °C.

### SDS-PAGE and Western Blot Analysis

After protein concentration quantification through bicinchoninic acid protein assay (BCA) (Pierce), 5μg (COLO205) and 15 ug (KATOIII) of protein extracts were loaded onto 4-15% Mini-PROTEAN®TGXTM precast polyacrylamide gels (Bio-Rad) for electrophoretic separation (SDS-PAGE) (Bio-Rad). Gels were then transferred onto nitrocellulose membranes (Amersham) and blocked for one hour with 5% bovine serum albumin (BSA) (Sigma-Aldrich) diluted in PBS containing 0.05% Tween 20 (Sigma-Aldrich) (PBS-T). For Western blotting, the membranes were incubated overnight at 4°C with Sialyl Lewis X (SLeX, CSLEX, 1:500, BD Bioscience) primary antibodies, followed by incubation with horseradish peroxidase-conjugated goat anti-mouse IgM (1:15000, Jackson ImmunoResearch). Protein bands were visualized using the ECL WB detection reagent and ChemiDoc Imaging System (Bio-Rad).

### Chicken embryo in vivo tumorigenesis assay

The chicken embryo chorioallantoic membrane (CAM) model [50]. was used to evaluate the angiogenic response and growth capability of COLO205 cells treated with different concentrations of monensin compared to DMSO control (n = 13 for each group). Briefly, fertilized chick (Gallus gallus) eggs obtained from commercial sources were incubated horizontally at 37.8°C in a humidified atmosphere and referred to embryonic day (E). On E3 a square window was opened in the shell after removal of 1.5–2 mL of albumin to allow detachment of the developing CAM. The window was sealed with a transparent adhesive tape and the eggs returned to the incubator. The window in the eggshell does not interfere in any way with the normal development of the chick embryo. COLO205 cells were resuspended in a solution containing monensin 200 nM or DMSO 0,01% (control) and vitrogel (TheWellBioscience). The cells’ containing solution (2×10^6^ cells per embryo) were placed on top of E10 growing CAM into a 3 mm silicon ring under sterile conditions. The eggs were re-sealed and returned to the incubator for an additional 6 days. The embryos were euthanized by adding 2 mL of fixative in the top of the CAM which is a very efficient and fast method. After removing the ring, the CAM was excised from the embryos, photographed ex ovo under a stereoscope, at 20x magnification (Olympus, SZX16 coupled with a DP71 camera). The area of CAM tumors was determined using the Cell A (Olympus) software.

Evaluation of tumor invasion was performed in a blind fashion way by two independent observers. The semi-quantitative evaluation took into consideration the quantity of human cytokeratin AE1/AE3 labeled cells present in the CAM mesenchyme.

According to the European Directive 2010/63/EU, ethical approval is not required for experiments using embryonic chicken. Correspondingly, the Portuguese law on animal welfare does not restrict the use of chicken eggs.

### Establishment of COLO205-LUC cell line and xenograft tumors of human cancer cells

COLO205-LUC were obtained obtained by lentivirus transfection of the COLO205-LUC with the CMV-Luciferase (firefly)-2A-GFP (Puro) viral particles (LVP020, Gentarget) containing the bioluminescent reporter protein luciferase. Cells were maintained in RPMI supplemented with puromycin (1 µg/ml) (invivogen).

Approximately 1×10^6^ or 5×10^6^ of COLO205-LUC cells were injected subcutaneously into dorsum of 8 to 10-week-old female N:NIH(s)II:nu/nu nude mice, aged 8–10 weeks and weighing approximately 20 g (i3S animal facility, Portugal). At day 7, when the tumor volume reached between 50-100 mm^3^ (in 1×10^6^ cells inoculum) or 200-250 mm^3^ (in 5×10^6^ cells inoculum), the COLO-205 LUC xenograft mice were randomized into 2 groups (n =5 a 6 mice per group) and treated with 100 μL of saline solution/PBS (control group) or Monensin (10mg/kg body weight) every two days. Monensin was dissolved in 100% ethanol to prepare 40mg/ml drug storage solution. *In vivo* luminescence imaging using the Ivis Lumina series III (PerkinElmer) at 4, 7, 14 and 21 days after cells injection. Animals were anesthetized with 5% (V/V) isoflurane for induction and 1–2% (V/V) for IVIS acquisition. During the treatment period, the tumor volume was measured every other day with a caliper and calculated as TV = (a x b x c) mm^3^ (Longest axis - a, shortest axis -b, and thickness - c). Animals were sacrificed, and the tumor and organs collected after 21 days pos-cells injection. All animal assays were conducted in strict conformity with the guidance of the European Union Directive 2010/63/EU, following a protocol previously authorized by the i3S Animal Ethics Commit-tee and the Competent Authority in Portugal (Direção-Geral de Alimentação e Veterinária (DGAV)) (reference 2021_05) regarding the humane endpoints, appropriate husbandry and protection of experimental animals. Mice were bred, cared and feeded *ad libidum* at the i3S Animal Facility (Porto, Portugal).

### Patient-Derived Tumor Organoid viability assay

Gastric tumor patient-derived organoids (PDOs) were previously established by the group. For this study, four tumoral PDOs were used to assess the effect of monensin in tumor growth. Monensin dose-response in patient-derived tumor organoids was assessed using the CellTiter-Glo® 3D Reagent (Promega Corporation). For that, 400 organoids fragments/well were resuspended in 20 µL of matrigel and seeded in a 96-well plate. After one day post seeding, organoids were treated with 0-400 nM of monensin for 6 days and the compound was renewed every 3 days. Upon treatement finished, the medium of each well was removed and 80 µL of fresh simple medium was added followed by 100 µL of CellTiter-Glo® 3D Reagent. Tumor organoids were mechanically disrupetd by up-and-down using a pipette and incubated from 25 min at RT. Luminescence signal was quantified through a SynergyMx™ MultiMode Microplate Reader (BioTek™). Viability values were normalized to the medium only-control (0 nM) as 100 % activity.

### Patient-Derived Tumor Organoids immunofluorescence analysis

Free floating fixed organoids were permeabilized with 50 mM of NH4Cl for 10 min and with PBS 0.3% Triton X-100 (Sigma Aldrich) for 15 min in rotation followed by washing steps using PBS 0.3% Triton X-100. Next, samples were blocked with the UltraVision Protein Block for 15 min followed by the primary antibody incubation with SLeX (CSLEX, 1:250, BD Bioscience) at 4°C overnight in rotation. After that, organoids were incubated for 45 min in rotation at room temperature with fluorescently-labeled secondary antibodies anti-mouse IgM Alexa Fluor® 594 (1:500, ThermoFisher Scientific). Nuclei were counter stained with DAPI (4′,6-diamidino-2-phenylindole; Sigma-Aldrich). Finally, organoids suspension was visualized using the inverted microscope Leica DMI6000-CS (Leica Microsystems, Germany). Images were acquired using the software LAS AF (Leica Microsystems, Germany).

### Subcutaneous implantation of Patient-Derived Tumor Organoids Xenograft (PDOX)

To develop gastric Patient-Derived Tumor Organoid Xenograft, 8 to 10-week-old female N:NIH(s)II:nu/nu nude mice, aged 8–10 weeks went through complete anesthesia by isoflurane 5% (V/V) exposure followed by a continuous 1.5-2% (V/V) isoflurane flow. Previously, before anesthesia, buprenorphine (0.08 mg/Kg) was subcutaneously injected for pain management. Then, a small incision was made in the upper middle part of the dorsal area and a subcutaneous “pocket” was created by blunt dissection. A 50 μL suspension of an determine/ chosen organoid dissolved in a matrigel matrix/PBS mixture with approximately 8000 to 12000 organoids was injected (resorting to a 200ul slashed pipette tip), into the “pocket”. Using a PGA nylon silk (Silkam), two or three stiches were done to close the surgical incision. Mice were daily monitored, and Paracetamol was added to the drinking water for the 72 hours after surgery. Mice were weighed and monitored for tumor development in every other day. The longest axis (a), shortest axis (b), and thickness (c) of each tumor were measured using a digital caliper and the total volume was calculated (TV= a x b x c). Tumor growth curves were constructed using the average tumor areas. When tumor reached 100-150mm^3^, the mice were divided into two groups and intraperitonially treated with monensin (10mg/kg body weight) or PBS (Control group) in every two days. Once tumors achieve at max 2000 mm^3^ or when it reaches 100 days after surgery the tumors were collected and fixed in 10% buffered formalin and embedded in paraffin. A histological analysis was performed using hematoxylin and eosin (HE) staining.

### Immunohistochemistry analysis

Excided CAMs and xenograft mice tumors and organs were fixed in 10% neutral-buffered formalin, paraffin-embedded for slide sections and stained with Hematoxilin and Eosin (H&E for histological examination) and immunostained for cytokeratin and SLeX detection to characterize the phenotype of CAM and xenograft mice tumors. Briefly, sections were dewaxed, rehydrate and the endogenous peroxidase activity was blocked with 3% H_2_O_2_ in methanol for 30 min. Then, antigen retrieval was achieved with citrate buffer pH 6, and sections were incubated with Animal-free blocker (Vector Laboratories) for 1h. For CAM tissues, a goat anti-chicken IgY antibody (1:100, Abcam) was added to blocking solution to reduced unspecific staining. Then, incubation with the monoclonal antibodies SLeX (CSLeX, 1:350, BD Biosciences), cytokeratins (AE1/AE3 1:3000 (CAM tissue) and 1:500 (mouse tissue), CellMarque) and Ki67 (D3B5, 1:500, CellSignaling) was performed overnight at 4°C, followed by biotinylated polyclonal rabbit anti-mouse immunoglobulins (DAKO) 30min incubation at room temperature and avidin/biotin complex detection (Vectastain). Finally, all samples were stained with 3,3-diaminobenzidine tetrahydrochloride (SigmaAldrich) containing 0.02% H_2_O_2_ and counterstaining of the nucleus was done with Mayer’s hematoxylin (Empredia).

### Statistical Analysis

Data were analyzed with GraphPad Prism 8.0 software (CA, USA). Quantitative data were expressed as the mean ± SEM of three biological replicates. The level of significance was assessed using Student’s t-test and one or two-way ANOVA. p - values of <0.01 were considered significant.

## Supporting information

Costa AF et al Supplemental Figure

## Data Availability

All data produced in the present study are available upon reasonable request to the authors

## Acknowledgments

The authors acknowledge the support of the i3S Scientific Platforms Animal facility, BioSciences Screening member of the national infrastructure PT-OPENSCREEN (NORTE-01-0145-FEDER-085468), HEMS and Advanced Light Microscopy members of the national infrastructure PPBI - Portuguese Platform of Bioimaging (PPBI-POCI-01-0145-FEDER-022122). We also acknowledge P.CCC: Centro Compreensivo de Cancro do Porto” - NORTE-01-0145-FEDER-072678, supported by Norte Portugal Regional Operational Programme (NORTE 2020), under the PORTUGAL 2020 Partnership Agreement, through the European Regional Development Fund (ERDF).

## Contributions

CG and CR supervised the study. CG designed the experiments. DC, AP, CG and AFM performed and analysed the HTS. LSF and FP establish the organoid models. AFC, ES, IFR, SL, JG, AT, CG and MTP performed the *in vivo* experiments. VF and LP analysed the RNASeq data. AFC, ES, DC, LSF, AT, RA, HOD, MP, AT and CG performed the remaining experiments and analysed the data. RB, FCG, FS and JB collected and provided patient tissues. FC evaluated the PDOX histology. CG and AFC wrote the original paper. All authors critically revised the paper.

## Ethical Statement

All procedures concerning the inclusion of patients were approved by the Centro Hospitalar Universitário de São João Ethics Committee (CES 223/2021). Also, all sample collection procedures were conducted following the patients’ informed consent.

