## Supplementary material for "Monensin as potential drug for treatment of SLeX-positive tumors": Costa AF et al Supplemental Figure

### Supplementary Figure 1

#### Prestwick Plate 1

DAPI (Cell Conter)

| Row La | 01 | 02 | 03 | 04 | 05 | 06 | 07 | 08 | 09 | 10 | 11 | 12 | 13 | 14 | 15 | 16 | 17 | 18 | 19 | 20 | 21 | 22 | 23 | 24 |
|---|---|---|---|---|---|---|---|---|---|---|---|---|---|---|---|---|---|---|---|---|---|---|---|---|
| A | 349 | 465 | 2057 | 1639 | 2153 | 3963 | 3309 | 2916 | 3615 | 4233 | 2536 | 3650 | 2991 | 5473 | 2831 | 3655 | 4230 | 4494 | 4260 | 3699 | 3741 | 3251 | 4842 | 4354 |
| B | 2636 | 134 | 2002 | 2276 | 1321 | 1150 | 1901 | 1496 | 1994 | 3197 | 3838 | 4807 | 3280 | 3815 | 4268 | 64 | 4696 | 3154 | 3404 | 3176 | 3821 | 3817 | 3939 | 2160 |
| C | 969 | 65 | 957 | 1854 | 1498 | 1996 | 2066 | 1887 | 1904 | 2185 | 1662 | 2731 | 2754 | 1863 | 2091 | 872 | 1773 | 2747 | 2709 | 2798 | 3099 | 86 | 3134 | 2171 |
| D | 1273 | 76 | 1377 | 1364 | 2803 | 3007 | 1574 | 1642 | 2774 | 1743 | 2316 | 2624 | 2943 | 2738 | 2626 | 2814 | 2378 | 3176 | 2382 | 2586 | 3020 | 2703 | 3027 | 4800 |
| E | 2032 | 331 | 1572 | 1127 | 2482 | 2634 | 2161 | 2329 | 2835 | 3531 | 3507 | 2473 | 3297 | 2956 | 3658 | 4025 | 4158 | 3482 | 4110 | 4291 | 3887 | 3845 | 3751 | 4323 |
| F | 2215 | 201 | 730 | 1607 | 2781 | 2505 | 4687 | 3563 | 3680 | 3356 | 3348 | 4050 | 4308 | 3403 | 3610 | 3966 | 3971 | 4550 | 4541 | 3962 | 3932 | 3254 | 5317 | 2370 |
| G | 35 | 61 | 2491 | 3033 | 2670 | 3001 | 3140 | 3030 | 1563 | 3203 | 3289 | 2255 | 3011 | 3336 | 933 | 3595 | 3874 | 3763 | 3716 | 3989 | 3653 | 3227 | 4437 | 4322 |
| H | 1886 | 501 | 1680 | 2793 | 2353 | 2412 | 3245 | 3642 | 3512 | 3488 | 3751 | 3601 | 3774 | 3906 | 5776 | 3782 | 4995 | 4964 | 4338 | 3596 | 178 | 2742 | 3107 | 4130 |
| I | 52 | 111 | 1618 | 2837 | 3177 | 3668 | 3129 | 3677 | 3478 | 3479 | 3642 | 4262 | 4375 | 3751 | 4472 | 4219 | 5158 | 4909 | 5185 | 4030 | 4495 | 3935 | 3207 | 5353 |
| J | 1227 | 40 | 804 | 3643 | 3049 | 3813 | 3150 | 2948 | 4304 | 3799 | 5139 | 3771 | 4844 | 4040 | 4541 | 4520 | 4802 | 4832 | 4313 | 4373 | 73 | 4202 | 3987 | 6372 |
| K | 561 | 69 | 1592 | 2415 | 2181 | 2446 | 3879 | 3465 | 3155 | 4157 | 4320 | 5124 | 5143 | 3723 | 4456 | 4607 | 4754 | 3104 | 5104 | 5167 | 5846 | 4458 | 4806 | 4623 |
| L | 44 | 71 | 926 | 3575 | 2354 | 2467 | 2842 | 3911 | 4791 | 3762 | 3912 | 3659 | 3555 | 4082 | 42 | 3989 | 4908 | 4805 | 4837 | 4249 | 5018 | 4465 | 4378 | 4839 |
| M | 164 | 58 | 1652 | 2982 | 1768 | 3731 | 1869 | 2235 | 2073 | 3322 | 3419 | 2225 | 2731 | 3170 | 4213 | 4591 | 4193 | 3413 | 4976 | 4479 | 3786 | 5123 | 4492 | 3515 |
| N | 730 | 185 | 739 | 972 | 898 | 2001 | 1803 | 3429 | 1515 | 2571 | 1922 | 4731 | 2735 | 3006 | 5047 | 5590 | 4165 | 4723 | 4589 | 3694 | 5164 | 4494 | 4109 | 4266 |
| O | 529 | 520 | 2308 | 646 | 1517 | 2983 | 2632 | 3190 | 1615 | 2381 | 3478 | 40 | 3766 | 3415 | 3352 | 4032 | 4682 | 2804 | 3144 | 3609 | 3713 | 2623 | 2134 | 3363 |
| P | 3204 | 1648 | 2426 | 3654 | 2710 | 3040 | 3334 | 2924 | 4064 | 3014 | 278 | 2096 | 3833 | 3905 | 2876 | 4957 | 4209 | 2049 | 4385 | 5611 | 5954 | 5196 | 4769 | 5720 |

TEXAS RED

| Row La | 01 | 02 | 03 | 04 | 05 | 06 | 07 | 08 | 09 | 10 | 11 | 12 | 13 | 14 | 15 | 16 | 17 | 18 | 19 | 20 | 21 | 22 | 23 | 24 |
|---|---|---|---|---|---|---|---|---|---|---|---|---|---|---|---|---|---|---|---|---|---|---|---|---|
| A | 114 | 113 | 328 | 419 | 339 | 271 | 327 | 362 | 291 | 299 | 298 | 313 | 313 | 247 | 337 | 316 | 292 | 284 | 289 | 292 | 311 | 306 | 254 | 269 |
| B | 114 | 120 | 354 | 338 | 454 | 428 | 367 | 332 | 322 | 300 | 294 | 275 | 288 | 326 | 284 | 1044 | 267 | 307 | 323 | 334 | 290 | 337 | 300 | 409 |
| C | 115 | 126 | 507 | 390 | 363 | 336 | 340 | 348 | 375 | 396 | 388 | 332 | 350 | 336 | 344 | 475 | 429 | 324 | 321 | 325 | 312 | 527 | 286 | 294 |
| D | 113 | 174 | 414 | 431 | 300 | 294 | 400 | 428 | 333 | 384 | 377 | 320 | 340 | 319 | 311 | 319 | 340 | 312 | 317 | 359 | 315 | 332 | 344 | 222 |
| E | 114 | 121 | 422 | 496 | 362 | 350 | 372 | 354 | 344 | 307 | 338 | 330 | 309 | 345 | 282 | 315 | 272 | 291 | 285 | 280 | 300 | 313 | 316 | 281 |
| F | 115 | 144 | 569 | 362 | 338 | 336 | 264 | 334 | 286 | 326 | 317 | 277 | 269 | 326 | 293 | 270 | 272 | 281 | 287 | 299 | 299 | 312 | 249 | 350 |
| G | 132 | 135 | 315 | 257 | 292 | 328 | 282 | 266 | 368 | 253 | 300 | 322 | 295 | 269 | 428 | 270 | 261 | 271 | 286 | 259 | 277 | 301 | 275 | 262 |
| H | 114 | 131 | 384 | 336 | 346 | 340 | 305 | 258 | 302 | 278 | 297 | 298 | 285 | 298 | 241 | 302 | 252 | 241 | 285 | 297 | 1127 | 286 | 342 | 287 |
| I | 121 | 150 | 386 | 316 | 295 | 275 | 314 | 257 | 301 | 322 | 310 | 252 | 291 | 278 | 282 | 273 | 258 | 261 | 268 | 295 | 279 | 287 | 351 | 246 |
| J | 121 | 253 | 449 | 317 | 313 | 304 | 320 | 329 | 297 | 302 | 274 | 304 | 251 | 273 | 274 | 301 | 276 | 248 | 265 | 265 | 821 | 289 | 293 | 230 |
| K | 117 | 145 | 419 | 364 | 360 | 300 | 289 | 304 | 357 | 297 | 294 | 260 | 260 | 280 | 267 | 267 | 275 | 301 | 241 | 272 | 238 | 261 | 269 | 297 |
| L | 115 | 259 | 622 | 317 | 341 | 367 | 310 | 315 | 271 | 295 | 291 | 282 | 295 | 314 | 844 | 283 | 253 | 264 | 274 | 295 | 255 | 307 | 282 | 270 |
| M | 132 | 172 | 411 | 301 | 340 | 306 | 373 | 337 | 350 | 328 | 273 | 347 | 346 | 289 | 275 | 283 | 296 | 303 | 272 | 285 | 302 | 261 | 301 | 282 |
| N | 134 | 143 | 560 | 418 | 579 | 337 | 393 | 318 | 352 | 382 | 376 | 287 | 319 | 335 | 296 | 251 | 276 | 269 | 280 | 286 | 245 | 282 | 301 | 294 |
| O | 116 | 117 | 392 | 596 | 386 | 318 | 339 | 323 | 385 | 355 | 308 | 574 | 301 | 292 | 275 | 293 | 274 | 248 | 318 | 300 | 335 | 329 | 415 | 336 |
| P | 113 | 116 | 428 | 299 | 304 | 313 | 321 | 268 | 309 | 326 | 768 | 341 | 286 | 289 | 276 | 267 | 270 | 351 | 271 | 255 | 233 | 269 | 241 | 221 |

#### Prestwick Plate 2

DAPI (Cell Conter)

| Row L | 01 | 02 | 03 | 04 | 05 | 06 | 07 | 08 | 09 | 10 | 11 | 12 | 13 | 14 | 15 | 16 | 17 | 18 | 19 | 20 | 21 | 22 | 23 | 24 |
|---|---|---|---|---|---|---|---|---|---|---|---|---|---|---|---|---|---|---|---|---|---|---|---|---|
| A | 1937 | 3128 | 2083 | 4296 | 2738 | 2199 | 1817 | 4362 | 3131 | 4211 | 3775 | 3399 | 2992 | 2854 | 4097 | 3640 | 3835 | 3795 | 3684 | 2883 | 2383 | 4213 | 4198 | 5438 |
| B | 1876 | 491 | 2123 | 2121 | 3002 | 1529 | 1674 | 2590 | 1824 | 2545 | 1557 | 2983 | 3704 | 1973 | 23 | 152 | 3703 | 2998 | 3763 | 3749 | 4068 | 3484 | 4329 | 3813 |
| C | 1614 | 155 | 1423 | 1838 | 1840 | 1847 | 2568 | 2588 | 2477 | 2052 | 2165 | 2062 | 2655 | 2285 | 2496 | 2581 | 2327 | 2850 | 2993 | 1288 | 2694 | 2990 | 3014 | 3403 |
| D | 793 | 128 | 1163 | 1871 | 1930 | 1602 | 2429 | 2275 | 2446 | 2169 | 2186 | 2327 | 3078 | 2727 | 3093 | 3893 | 2668 | 3363 | 3280 | 3349 | 3326 | 2985 | 4131 | 4003 |
| E | 1048 | 147 | 2372 | 1238 | 2972 | 2509 | 2790 | 3208 | 2988 | 3749 | 3094 | 3550 | 2721 | 3208 | 1189 | 4210 | 3429 | 4314 | 3842 | 3803 | 3815 | 3412 | 4081 | 3198 |
| F | 1380 | 115 | 2210 | 2319 | 2933 | 3503 | 3649 | 3517 | 3614 | 3759 | 4122 | 3881 | 4701 | 3351 | 3660 | 3415 | 4768 | 4864 | 4689 | 3969 | 4390 | 3669 | 4646 | 4325 |
| G | 218 | 93 | 2333 | 2912 | 2802 | 3096 | 3213 | 3799 | 3456 | 3844 | 2847 | 2067 | 4012 | 448 | 3435 | 19 | 4019 | 15 | 4684 | 4574 | 4045 | 4609 | 3853 | 3928 |
| H | 2058 | 65 | 1769 | 2615 | 3472 | 3289 | 3041 | 3366 | 3574 | 2609 | 3898 | 3491 | 3948 | 4081 | 3719 | 4608 | 262 | 409 | 4477 | 4166 | 4500 | 3547 | 3064 | 5608 |
| I | 148 | 53 | 1673 | 2721 | 2402 | 3715 | 115 | 2569 | 4145 | 3606 | 4433 | 4311 | 4472 | 4787 | 5133 | 4631 | 4785 | 2862 | 5528 | 4424 | 4196 | 3881 | 4760 | 5235 |
| J | 96 | 64 | 2094 | 3252 | 116 | 3420 | 3887 | 3499 | 2984 | 3588 | 3474 | 3978 | 4422 | 4641 | 5347 | 4731 | 5424 | 216 | 5023 | 5614 | 5043 | 4008 | 5402 | 4798 |
| K | 111 | 107 | 2142 | 3288 | 3241 | 3706 | 587 | 3259 | 3772 | 3010 | 3394 | 4083 | 2749 | 4907 | 4864 | 4432 | 5428 | 4900 | 5538 | 4734 | 5103 | 4656 | 4976 | 5878 |
| L | 424 | 77 | 1325 | 2677 | 3155 | 3278 | 2990 | 3315 | 3580 | 3212 | 3478 | 4384 | 3047 | 4392 | 4415 | 4546 | 5136 | 5260 | 5321 | 4891 | 4936 | 4943 | 4832 | 3883 |
| M | 472 | 116 | 1754 | 2067 | 2719 | 2940 | 2334 | 3182 | 2608 | 3383 | 186 | 282 | 3121 | 2789 | 3143 | 4215 | 1950 | 4620 | 4370 | 5387 | 5342 | 5801 | 5221 | 5934 |
| N | 660 | 45 | 442 | 1523 | 1722 | 2386 | 1352 | 1459 | 1743 | 1792 | 158 | 195 | 1969 | 5859 | 4287 | 3896 | 5013 | 4148 | 4835 | 4635 | 4184 | 4835 | 5301 | 5450 |
| O | 245 | 153 | 1471 | 1200 | 1823 | 1484 | 2573 | 2445 | 3914 | 2492 | 1888 | 1695 | 250 | 3590 | 4765 | 4801 | 2866 | 2828 | 3123 | 3908 | 4669 | 3915 | 4390 | 4985 |
| P | 275 | 54 | 2445 | 1636 | 1971 | 1552 | 5370 | 3629 | 4795 | 4456 | 2047 | 4211 | 3901 | 4190 | 5315 | 5011 | 5333 | 6844 | 5008 | 5318 | 4825 | 5554 | 5286 | 5606 |

TEXAS RED

| Row L | 01 | 02 | 03 | 04 | 05 | 06 | 07 | 08 | 09 | 10 | 11 | 12 | 13 | 14 | 15 | 16 | 17 | 18 | 19 | 20 | 21 | 22 | 23 | 24 |
|---|---|---|---|---|---|---|---|---|---|---|---|---|---|---|---|---|---|---|---|---|---|---|---|---|
| A | 107 | 107 | 246 | 207 | 223 | 265 | 275 | 208 | 239 | 216 | 233 | 245 | 243 | 238 | 219 | 211 | 211 | 221 | 216 | 216 | 249 | 258 | 204 | 194 |
| B | 110 | 114 | 278 | 300 | 274 | 325 | 286 | 250 | 296 | 293 | 314 | 282 | 235 | 302 | 453 | 418 | 218 | 252 | 223 | 249 | 222 | 230 | 227 | 204 |
| C | 108 | 86 | 316 | 309 | 311 | 298 | 262 | 257 | 256 | 301 | 306 | 259 | 287 | 286 | 270 | 267 | 288 | 252 | 257 | 411 | 283 | 266 | 263 | 227 |
| D | 112 | 142 | 343 | 331 | 308 | 316 | 282 | 291 | 271 | 281 | 288 | 264 | 238 | 255 | 261 | 226 | 266 | 243 | 240 | 217 | 249 | 242 | 227 | 217 |
| E | 111 | 114 | 282 | 319 | 288 | 276 | 259 | 236 | 285 | 226 | 254 | 261 | 263 | 237 | 326 | 225 | 250 | 237 | 245 | 250 | 244 | 236 | 243 | 252 |
| F | 113 | 144 | 316 | 286 | 259 | 225 | 247 | 248 | 255 | 228 | 236 | 252 | 234 | 250 | 245 | 249 | 216 | 221 | 229 | 246 | 236 | 234 | 231 | 221 |
| G | 119 | 145 | 276 | 273 | 243 | 264 | 256 | 234 | 244 | 226 | 260 | 314 | 238 | 551 | 248 | 258 | 234 | 375 | 204 | 212 | 217 | 221 | 220 | 231 |
| H | 113 | 190 | 360 | 273 | 258 | 251 | 245 | 281 | 245 | 286 | 231 | 243 | 251 | 234 | 242 | 236 | 621 | 523 | 229 | 237 | 234 | 241 | 296 | 203 |
| I | 136 | 136 | 356 | 274 | 300 | 247 | 510 | 269 | 228 | 279 | 258 | 215 | 259 | 226 | 241 | 249 | 230 | 293 | 227 | 232 | 254 | 260 | 239 | 229 |
| J | 121 | 115 | 296 | 277 | 820 | 253 | 246 | 284 | 269 | 278 | 253 | 276 | 257 | 227 | 243 | 247 | 238 | 569 | 215 | 225 | 230 | 245 | 207 | 257 |
| K | 132 | 121 | 300 | 284 | 260 | 255 | 478 | 282 | 255 | 308 | 289 | 252 | 313 | 232 | 229 | 223 | 243 | 252 | 236 | 253 | 240 | 257 | 264 | 231 |
| L | 118 | 148 | 382 | 313 | 298 | 299 | 311 | 277 | 292 | 285 | 257 | 238 | 313 | 251 | 271 | 272 | 240 | 238 | 231 | 238 | 254 | 245 | 255 | 288 |
| M | 122 | 231 | 357 | 310 | 300 | 275 | 314 | 314 | 346 | 284 | 841 | 596 | 306 | 278 | 297 | 274 | 376 | 259 | 258 | 250 | 255 | 225 | 240 | 212 |
| N | 146 | 165 | 664 | 380 | 385 | 332 | 420 | 421 | 334 | 371 | 571 | 650 | 354 | 222 | 247 | 251 | 248 | 273 | 257 | 278 | 258 | 232 | 246 | 226 |
| O | 159 | 126 | 466 | 412 | 355 | 371 | 305 | 321 | 255 | 333 | 354 | 337 | 648 | 314 | 241 | 261 | 304 | 329 | 252 | 302 | 247 | 269 | 257 | 246 |
| P | 116 | 118 | 319 | 341 | 360 | 387 | 225 | 268 | 244 | 264 | 345 | 256 | 251 | 242 | 247 | 243 | 248 | 223 | 252 | 240 | 244 | 244 | 249 | 199 |

#### Prestwick Plate 3

DAPI (Cell Conter)

| Row La | 01 | 02 | 03 | 04 | 05 | 06 | 07 | 08 | 09 | 10 | 11 | 12 | 13 | 14 | 15 | 16 | 17 | 18 | 19 | 20 | 21 | 22 | 23 | 24 |
|---|---|---|---|---|---|---|---|---|---|---|---|---|---|---|---|---|---|---|---|---|---|---|---|---|
| A | 1132 | 2908 | 1988 | 2903 | 1195 | 2380 | 2478 | 1892 | 1780 | 3477 | 3593 | 2728 | 2090 | 2013 | 2430 | 3473 | 2674 | 3048 | 2588 | 2960 | 3003 | 3830 | 2389 | 4793 |
| B | 81 | 2519 | 2365 | 2038 | 344 | 2176 | 2155 | 1305 | 3062 | 1863 | 2160 | 2173 | 1996 | 1184 | 396 | 2219 | 2857 | 2507 | 3356 | 4224 | 3530 | 3628 | 286 | 3153 |
| C | 1754 | 1552 | 2 | 0 | 59 | 4659 | 3296 | 3704 | 2209 | 1006 | 2635 | 4372 | 10 | 2684 | 3041 | 3146 | 3741 | 3371 | 3120 | 3254 | 4513 | 3239 | 3465 | 3598 |
| D | 481 | 647 | 9 | 2492 | 2446 | 56 | 1751 | 3652 | 2520 | 3643 | 3101 | 3707 | 2155 | 2746 | 2804 | 4101 | 2742 | 3350 | 3040 | 3036 | 2978 | 2600 | 324 | 5316 |
| E | 2791 | 1470 | 1725 | 3037 | 2887 | 3378 | 3250 | 3025 | 3322 | 3691 | 3656 | 2452 | 3093 | 3184 | 3550 | 3063 | 4036 | 509 | 3472 | 3024 | 2448 | 2692 | 3169 | 3248 |
| F | 32 | 1719 | 3330 | 3064 | 3716 | 3380 | 3301 | 3039 | 3474 | 2935 | 3623 | 4419 | 4242 | 3660 | 3013 | 4129 | 2923 | 2927 | 3996 | 3822 | 3006 | 3325 | 396 | 4829 |
| G | 801 | 2390 | 4278 | 4058 | 4165 | 4764 | 4947 | 4084 | 3835 | 3303 | 4947 | 4110 | 3040 | 4238 | 4184 | 4373 | 4148 | 4908 | 5010 | 4267 | 5258 | 4050 | 4469 | 5415 |
| H | 92 | 2309 | 3321 | 2271 | 4408 | 3431 | 3749 | 5282 | 4154 | 2875 | 4821 | 4646 | 4973 | 4724 | 4870 | 4920 | 5599 | 5062 | 4889 | 5889 | 4979 | 3982 | 374 | 5068 |
| I | 1676 | 2784 | 3707 | 3776 | 3293 | 4697 | 4907 | 4521 | 4564 | 4448 | 4330 | 5051 | 5651 | 4567 | 5387 | 4259 | 5263 | 4098 | 4834 | 4713 | 497 | 4532 | 4628 | 5105 |
| J | 47 | 1833 | 2822 | 4335 | 848 | 4005 | 3053 | 4418 | 3769 | 3959 | 5217 | 4139 | 5001 | 4678 | 4945 | 4943 | 4695 | 4638 | 5274 | 5636 | 4972 | 4329 | 593 | 5002 |
| K | 1691 | 2785 | 3799 | 4698 | 4639 | 6417 | 4822 | 5013 | 5107 | 4829 | 5260 | 4334 | 4583 | 5440 | 5234 | 985 | 4446 | 5280 | 5307 | 5306 | 6059 | 4175 | 6259 | 4051 |
| L | 53 | 3350 | 3195 | 5291 | 6047 | 5656 | 4278 | 5278 | 5342 | 4911 | 4617 | 5616 | 4348 | 5621 | 5193 | 5969 | 5182 | 5235 | 4248 | 5008 | 4620 | 5465 | 733 | 5447 |
| M | 1261 | 1735 | 4575 | 3615 | 4846 | 50 | 3937 | 5745 | 4795 | 4114 | 3566 | 5102 | 6014 | 5460 | 5636 | 5619 | 5183 | 5540 | 6631 | 5497 | 6954 | 497 | 6039 | 5990 |
| N | 122 | 3754 | 778 | 4557 | 5162 | 4564 | 3836 | 4100 | 6013 | 4723 | 3337 | 4236 | 1570 | 5464 | 6802 | 1943 | 6245 | 5292 | 5949 | 4957 | 5970 | 5709 | 908 | 4996 |
| O | 3294 | 2147 | 2632 | 3073 | 2690 | 3331 | 2958 | 2820 | 4027 | 2828 | 2838 | 4853 | 4486 | 4903 | 4232 | 4515 | 4263 | 3881 | 3530 | 3977 | 3658 | 4535 | 3869 | 6102 |
| P | 86 | 2055 | 2597 | 2832 | 242 | 3073 | 3477 | 2831 | 3596 | 2309 | 2385 | 5146 | 4612 | 4298 | 4355 | 6750 | 6532 | 8086 | 6698 | 6490 | 6001 | 6302 | 374 | 6304 |

TEXAS RED

| Row La | 01 | 02 | 03 | 04 | 05 | 06 | 07 | 08 | 09 | 10 | 11 | 12 | 13 | 14 | 15 | 16 | 17 | 18 | 19 | 20 | 21 | 22 | 23 | 24 |
|---|---|---|---|---|---|---|---|---|---|---|---|---|---|---|---|---|---|---|---|---|---|---|---|---|
| A | 108 | 108 | 292 | 247 | 359 | 232 | 298 | 285 | 273 | 232 | 242 | 242 | 279 | 269 | 288 | 239 | 242 | 260 | 243 | 244 | 242 | 234 | 259 | 187 |
| B | 113 | 111 | 314 | 294 | 551 | 286 | 286 | 380 | 260 | 259 | 288 | 275 | 277 | 345 | 573 | 257 | 269 | 236 | 250 | 231 | 252 | 238 | 601 | 262 |
| C | 110 | 110 | 59 | 0 | 513 | 217 | 232 | 253 | 276 | 365 | 246 | 223 | 118 | 257 | 241 | 252 | 225 | 260 | 245 | 232 | 235 | 249 | 243 | 214 |
| D | 110 | 112 | 79 | 270 | 303 | 488 | 352 | 250 | 274 | 243 | 268 | 249 | 296 | 272 | 287 | 230 | 276 | 251 | 252 | 251 | 241 | 264 | 550 | 203 |
| E | 111 | 113 | 330 | 264 | 292 | 243 | 252 | 232 | 270 | 227 | 256 | 298 | 265 | 266 | 238 | 266 | 253 | 150 | 259 | 278 | 282 | 210 | 267 | 242 |
| F | 113 | 115 | 257 | 240 | 280 | 255 | 289 | 274 | 276 | 271 | 272 | 247 | 260 | 243 | 230 | 237 | 270 | 272 | 208 | 228 | 266 | 236 | 559 | 213 |
| G | 114 | 113 | 235 | 232 | 228 | 230 | 201 | 210 | 221 | 261 | 215 | 205 | 266 | 216 | 228 | 225 | 209 | 201 | 212 | 229 | 197 | 233 | 248 | 217 |
| H | 134 | 113 | 239 | 301 | 251 | 271 | 263 | 238 | 270 | 274 | 249 | 251 | 233 | 248 | 227 | 242 | 237 | 242 | 249 | 224 | 235 | 235 | 571 | 248 |
| I | 115 | 113 | 258 | 263 | 293 | 261 | 248 | 257 | 244 | 263 | 270 | 240 | 243 | 263 | 248 | 270 | 245 | 289 | 239 | 267 | 545 | 265 | 283 | 269 |
| J | 132 | 113 | 314 | 254 | 517 | 263 | 281 | 243 | 267 | 278 | 249 | 245 | 262 | 253 | 244 | 256 | 251 | 251 | 247 | 221 | 250 | 240 | 570 | 268 |
| K | 118 | 116 | 279 | 249 | 263 | 236 | 264 | 256 | 237 | 247 | 259 | 262 | 262 | 247 | 248 | 395 | 274 | 257 | 250 | 258 | 221 | 277 | 236 | 383 |
| L | 157 | 114 | 263 | 244 | 226 | 225 | 279 | 255 | 259 | 263 | 236 | 262 | 286 | 239 | 253 | 239 | 250 | 245 | 290 | 267 | 263 | 243 | 651 | 251 |
| M | 112 | 113 | 271 | 278 | 240 | 522 | 291 | 248 | 231 | 239 | 310 | 258 | 241 | 260 | 252 | 249 | 252 | 256 | 239 | 250 | 219 | 472 | 235 | 244 |
| N | 116 | 114 | 683 | 286 | 284 | 288 | 318 | 297 | 253 | 278 | 320 | 298 | 414 | 275 | 247 | 385 | 260 | 271 | 237 | 258 | 236 | 242 | 592 | 270 |
| O | 113 | 115 | 284 | 271 | 238 | 298 | 245 | 291 | 252 | 272 | 222 | 269 | 239 | 227 | 270 | 242 | 265 | 249 | 257 | 226 | 242 | 243 | 284 | 229 |
| P | 113 | 114 | 273 | 312 | 711 | 293 | 329 | 307 | 301 | 301 | 312 | 265 | 285 | 244 | 246 | 212 | 216 | 206 | 219 | 213 | 209 | 217 | 652 | 212 |

#### Prestwick Plate 4

DAPI (Cell Conter)

| Row L | 01 | 02 | 03 | 04 | 05 | 06 | 07 | 08 | 09 | 10 | 11 | 12 | 13 | 14 | 15 | 16 | 17 | 18 | 19 | 20 | 21 | 22 | 23 | 24 |
|---|---|---|---|---|---|---|---|---|---|---|---|---|---|---|---|---|---|---|---|---|---|---|---|---|
| A | 3656 | 4417 | 3273 | 3166 | 3667 | 6012 | 5101 | 2837 | 2848 | 4775 | 425 | 5261 | 4731 | 5390 | 5404 | 4682 | 5890 | 5357 | 5459 | 5433 | 4880 | 6048 | 3421 | 3611 |
| B | 3647 | 3777 | 2731 | 3668 | 5888 | 6266 | 3748 | 4013 | 4669 | 3777 | 3823 | 3599 | 4573 | 4431 | 3734 | 4578 | 6561 | 5166 | 3816 | 6567 | 4871 | 2980 | 4175 | 3738 |
| C | 4214 | 2392 | 3253 | 4161 | 4665 | 5562 | 4184 | 4922 | 5317 | 4889 | 4684 | 4830 | 4944 | 700 | 4591 | 4840 | 4881 | 4509 | 5157 | 4957 | 4072 | 4369 | 5185 | 3509 |
| D | 3925 | 2485 | 3102 | 3466 | 2799 | 3700 | 4075 | 3888 | 4237 | 2960 | 4513 | 5033 | 4703 | 3827 | 4094 | 3979 | 3759 | 4094 | 4617 | 3620 | 3855 | 3155 | 3607 | 3680 |
| E | 2580 | 2514 | 3356 | 4604 | 3910 | 4539 | 4103 | 4159 | 4941 | 4808 | 5080 | 4365 | 5234 | 4327 | 4619 | 5247 | 5086 | 4582 | 5075 | 5549 | 5194 | 2748 | 4175 | 4504 |
| F | 3496 | 2154 | 2701 | 2641 | 3381 | 3183 | 3826 | 3835 | 3436 | 4396 | 4219 | 5411 | 4311 | 3202 | 4149 | 3183 | 221 | 3948 | 4103 | 4435 | 4627 | 4187 | 4667 | 4207 |
| G | 299 | 568 | 566 | 1355 | 1473 | 1041 | 1141 | 1452 | 1874 | 1792 | 1848 | 1920 | 2218 | 1794 | 2246 | 2365 | 1363 | 2258 | 3198 | 2566 | 2051 | 49 | 2638 | 349 |
| H | 1002 | 861 | 368 | 660 | 1620 | 1625 | 1995 | 1610 | 1590 | 1412 | 2123 | 1942 | 2843 | 1838 | 2224 | 1068 | 2523 | 2960 | 2473 | 1810 | 2691 | 2171 | 2930 | 83 |
| I | 980 | 1028 | 1347 | 1646 | 2462 | 1767 | 2325 | 2474 | 1967 | 2375 | 1832 | 2497 | 2331 | 2926 | 2802 | 2635 | 3338 | 2884 | 2369 | 2694 | 3367 | 3078 | 3424 | 2298 |
| J | 1765 | 1419 | 1485 | 1485 | 2539 | 2369 | 2540 | 2287 | 3295 | 2609 | 2916 | 3026 | 2798 | 2042 | 2033 | 2784 | 2969 | 3328 | 2247 | 2467 | 2431 | 2179 | 2861 | 6452 |
| K | 446 | 941 | 2556 | 3170 | 2590 | 2884 | 2725 | 2884 | 2723 | 3167 | 2863 | 3423 | 2737 | 3567 | 3007 | 3711 | 3175 | 3593 | 3421 | 4179 | 3215 | 3720 | 3445 | 5555 |
| L | 2825 | 1766 | 111 | 2157 | 2919 | 2771 | 2831 | 2711 | 3216 | 3494 | 3052 | 2969 | 2795 | 3314 | 3183 | 2975 | 3491 | 2748 | 3655 | 2384 | 3411 | 3231 | 3521 | 2477 |
| M | 1754 | 2750 | 2186 | 2014 | 2399 | 3178 | 2562 | 2094 | 4421 | 3359 | 3543 | 3684 | 3407 | 2903 | 2585 | 4434 | 3116 | 3556 | 2779 | 3202 | 3599 | 52 | 4326 | 744 |
| N | 2886 | 3124 | 831 | 38 | 2266 | 3202 | 150 | 2706 | 3835 | 3272 | 2559 | 2892 | 3508 | 3314 | 115 | 817 | 4829 | 3760 | 3865 | 3710 | 2939 | 3231 | 2005 | 2702 |
| O | 1572 | 3286 | 1843 | 1742 | 1217 | 2530 | 3268 | 2470 | 1557 | 1297 | 2530 | 3220 | 2183 | 2580 | 3868 | 2794 | 3129 | 2907 | 2991 | 2357 | 115 | 1750 | 2534 | 14 |
| P | 1790 | 1857 | 2149 | 3456 | 3279 | 403 | 2328 | 320 | 2703 | 2661 | 3937 | 4349 | 2925 | 2419 | 4607 | 3327 | 5497 | 2180 | 4547 | 2291 | 400 | 2522 | 3783 | 323 |

TEXAS RED

| Row L | 01 | 02 | 03 | 04 | 05 | 06 | 07 | 08 | 09 | 10 | 11 | 12 | 13 | 14 | 15 | 16 | 17 | 18 | 19 | 20 | 21 | 22 | 23 | 24 |
|---|---|---|---|---|---|---|---|---|---|---|---|---|---|---|---|---|---|---|---|---|---|---|---|---|
| A | 260 | 303 | 322 | 329 | 361 | 253 | 283 | 364 | 366 | 313 | 780 | 280 | 297 | 277 | 246 | 282 | 254 | 269 | 269 | 267 | 321 | 282 | 332 | 117 |
| B | 350 | 341 | 342 | 302 | 259 | 265 | 293 | 281 | 260 | 303 | 323 | 313 | 261 | 282 | 334 | 282 | 249 | 286 | 270 | 265 | 270 | 364 | 284 | 116 |
| C | 332 | 336 | 329 | 301 | 297 | 289 | 284 | 272 | 281 | 278 | 287 | 256 | 278 | 490 | 289 | 279 | 287 | 298 | 302 | 281 | 307 | 321 | 277 | 117 |
| D | 321 | 341 | 344 | 293 | 318 | 319 | 323 | 296 | 316 | 317 | 306 | 257 | 269 | 295 | 305 | 322 | 307 | 309 | 289 | 313 | 332 | 338 | 327 | 123 |
| E | 286 | 317 | 358 | 312 | 292 | 263 | 323 | 277 | 304 | 252 | 281 | 305 | 290 | 322 | 289 | 286 | 296 | 281 | 293 | 288 | 284 | 372 | 312 | 117 |
| F | 352 | 376 | 316 | 354 | 319 | 307 | 340 | 316 | 346 | 301 | 340 | 295 | 313 | 332 | 305 | 382 | 871 | 304 | 316 | 311 | 300 | 330 | 317 | 117 |
| G | 724 | 542 | 586 | 412 | 410 | 399 | 485 | 352 | 382 | 389 | 380 | 384 | 318 | 384 | 372 | 371 | 340 | 328 | 373 | 363 | 357 | 1036 | 364 | 116 |
| H | 544 | 526 | 597 | 533 | 430 | 398 | 408 | 446 | 435 | 474 | 429 | 432 | 364 | 435 | 389 | 505 | 324 | 349 | 361 | 414 | 310 | 379 | 361 | 116 |
| I | 576 | 502 | 497 | 396 | 360 | 382 | 377 | 370 | 387 | 382 | 305 | 275 | 337 | 312 | 363 | 340 | 315 | 363 | 341 | 319 | 328 | 369 | 297 | 115 |
| J | 380 | 471 | 445 | 390 | 368 | 414 | 370 | 395 | 322 | 346 | 384 | 363 | 379 | 380 | 392 | 348 | 362 | 315 | 372 | 394 | 347 | 400 | 341 | 116 |
| K | 680 | 546 | 386 | 348 | 404 | 369 | 371 | 355 | 367 | 362 | 389 | 328 | 404 | 339 | 371 | 358 | 360 | 323 | 350 | 334 | 339 | 329 | 330 | 114 |
| L | 434 | 490 | 1375 | 409 | 372 | 362 | 408 | 430 | 377 | 337 | 363 | 345 | 389 | 359 | 341 | 342 | 345 | 346 | 321 | 409 | 375 | 372 | 393 | 116 |
| M | 416 | 366 | 418 | 384 | 404 | 344 | 366 | 436 | 304 | 336 | 354 | 348 | 364 | 364 | 351 | 284 | 351 | 355 | 386 | 311 | 332 | 534 | 306 | 116 |
| N | 360 | 401 | 487 | 977 | 442 | 350 | 998 | 384 | 356 | 366 | 400 | 376 | 350 | 347 | 747 | 705 | 290 | 327 | 339 | 342 | 419 | 373 | 466 | 86 |
| O | 420 | 362 | 436 | 491 | 500 | 367 | 350 | 405 | 472 | 541 | 378 | 355 | 414 | 392 | 316 | 305 | 379 | 380 | 381 | 427 | 941 | 482 | 419 | 113 |
| P | 446 | 439 | 365 | 355 | 384 | 645 | 364 | 505 | 317 | 378 | 297 | 402 | 425 | 389 | 314 | 336 | 298 | 427 | 291 | 429 | 728 | 337 | 307 | 114 |

### Diverset Plate 1

**DAPI (Cell Conter)**

| Row Lab | 01 | 02 | 03 | 04 | 05 | 06 | 07 | 08 | 09 | 10 | 11 | 12 | 13 | 14 | 15 | 16 | 17 | 18 | 19 | 20 | 21 | 22 | 23 | 24 |
|---|---|---|---|---|---|---|---|---|---|---|---|---|---|---|---|---|---|---|---|---|---|---|---|---|
| A | 516 | 95 | 950 | 2837 | 3333 | 69 | 4036 | 1643 | 3251 | 103 | 2520 | 60 | 4045 | 111 | 4372 | 4233 | 3901 | 58 | 3606 | 2938 | 2743 | 45 | 888 | 90 |
| B | 15 | 206 | 321 | 275 | 335 | 3058 | 243 | 2800 | 333 | 848 | 291 | 2518 | 368 | 3723 | 278 | 3349 | 372 | 392 | 273 | 372 | 330 | 306 | 207 | 131 |
| C | 248 | 431 | 1325 | 102 | 2746 | 3525 | 2907 | 3286 | 2924 | 3596 | 3255 | 4019 | 4348 | 4441 | 5 | 3863 | 3642 | 3841 | 3490 | 3978 | 4135 | 3467 | 3054 | 20 |
| D | 32 | 2476 | 25 | 2765 | 897 | 3320 | 3008 | 3851 | 3434 | 2504 | 3593 | 315 | 4138 | 3539 | 4293 | 3684 | 4356 | 3079 | 42 | 3764 | 3784 | 4116 | 203 | 9 |
| E | 1136 | 196 | 1004 | 409 | 2378 | 2874 | 3196 | 3247 | 3597 | 4002 | 2526 | 2191 | 3716 | 3891 | 3406 | 3928 | 3416 | 4214 | 2965 | 425 | 1019 | 427 | 3275 | 167 |
| F | 190 | 304 | 547 | 309 | 348 | 349 | 3024 | 3384 | 4524 | 3279 | 3621 | 3218 | 3800 | 4457 | 422 | 595 | 390 | 4129 | 396 | 451 | 409 | 4006 | 410 | 264 |
| G | 52 | 18 | 336 | 1654 | 1467 | 1583 | 1238 | 282 | 1928 | 2594 | 1845 | 266 | 2118 | 2291 | 2535 | 2275 | 2956 | 3095 | 2678 | 2947 | 2504 | 3426 | 2490 | 46 |
| H | 18 | 648 | 963 | 1080 | 1292 | 1090 | 4 | 1271 | 1799 | 2261 | 1722 | 588 | 3058 | 2433 | 1509 | 2843 | 3403 | 748 | 2891 | 2887 | 2423 | 3098 | 18 | 111 |
| I | 2266 | 103 | 688 | 248 | 2014 | 326 | 1822 | 342 | 2588 | 2913 | 2440 | 3854 | 3347 | 3590 | 2730 | 560 | 4020 | 3658 | 3279 | 3630 | 3430 | 350 | 748 | 284 |
| J | 100 | 175 | 179 | 257 | 2333 | 2504 | 2060 | 2314 | 2666 | 2740 | 447 | 1020 | 3219 | 3605 | 410 | 3319 | 3809 | 272 | 4188 | 3560 | 523 | 423 | 323 | 310 |
| K | 958 | 145 | 310 | 269 | 1271 | 246 | 276 | 2473 | 155 | 2336 | 2346 | 2844 | 2653 | 3309 | 1243 | 3324 | 2954 | 2898 | 3023 | 3100 | 2776 | 365 | 2296 | 73 |
| L | 6 | 156 | 214 | 226 | 376 | 330 | 2022 | 2309 | 2422 | 2191 | 351 | 1152 | 462 | 3306 | 2745 | 2780 | 473 | 2622 | 34 | 2678 | 473 | 2881 | 248 | 55 |
| M | 19 | 149 | 189 | 239 | 1427 | 234 | 2221 | 299 | 2127 | 311 | 2851 | 338 | 307 | 379 | 2474 | 2871 | 2890 | 293 | 2710 | 2486 | 2975 | 438 | 646 | 231 |
| N | 72 | 97 | 161 | 177 | 175 | 189 | 216 | 299 | 294 | 1112 | 215 | 1654 | 260 | 2469 | 252 | 327 | 292 | 264 | 2006 | 390 | 246 | 295 | 283 | 298 |
| O | 946 | 19 | 809 | 144 | 1568 | 164 | 1588 | 192 | 1826 | 177 | 2169 | 272 | 3260 | 282 | 2384 | 2882 | 3023 | 230 | 2805 | 277 | 2634 | 217 | 2500 | 134 |
| P | 3 | 63 | 85 | 38 | 59 | 405 | 156 | 98 | 126 | 114 | 29 | 3455 | 127 | 3425 | 154 | 3945 | 2852 | 3847 | 3263 | 4190 | 362 | 195 | 44 | 77 |

**TEXAS RED**

| Row Lab | 01 | 02 | 03 | 04 | 05 | 06 | 07 | 08 | 09 | 10 | 11 | 12 | 13 | 14 | 15 | 16 | 17 | 18 | 19 | 20 | 21 | 22 | 23 | 24 |
|---|---|---|---|---|---|---|---|---|---|---|---|---|---|---|---|---|---|---|---|---|---|---|---|---|
| A | 433 | 657 | 407 | 312 | 269 | 715 | 263 | 429 | 293 | 661 | 270 | 739 | 321 | 772 | 290 | 325 | 294 | 777 | 260 | 361 | 281 | 585 | 620 | 180 |
| B | 635 | 772 | 724 | 700 | 711 | 345 | 732 | 355 | 661 | 594 | 664 | 400 | 648 | 328 | 647 | 300 | 665 | 643 | 622 | 633 | 638 | 672 | 792 | 125 |
| C | 113 | 476 | 329 | 605 | 266 | 272 | 295 | 266 | 295 | 259 | 292 | 256 | 242 | 234 | 631 | 268 | 237 | 266 | 257 | 271 | 248 | 261 | 260 | 89 |
| D | 550 | 315 | 605 | 277 | 453 | 305 | 302 | 264 | 289 | 311 | 261 | 701 | 271 | 271 | 251 | 229 | 265 | 290 | 435 | 260 | 260 | 250 | 646 | 178 |
| E | 261 | 774 | 353 | 625 | 366 | 296 | 300 | 314 | 280 | 291 | 289 | 371 | 289 | 295 | 304 | 277 | 302 | 253 | 309 | 652 | 432 | 653 | 333 | 125 |
| F | 464 | 761 | 710 | 737 | 669 | 728 | 315 | 310 | 278 | 319 | 302 | 315 | 293 | 282 | 587 | 593 | 549 | 264 | 573 | 552 | 674 | 275 | 684 | 125 |
| G | 661 | 453 | 590 | 333 | 374 | 379 | 378 | 657 | 323 | 300 | 348 | 626 | 345 | 324 | 300 | 302 | 320 | 292 | 320 | 324 | 319 | 267 | 327 | 136 |
| H | 428 | 482 | 427 | 500 | 383 | 450 | 223 | 444 | 331 | 301 | 355 | 553 | 268 | 321 | 387 | 304 | 266 | 518 | 310 | 289 | 315 | 303 | 632 | 120 |
| I | 239 | 587 | 496 | 662 | 330 | 608 | 346 | 628 | 311 | 308 | 302 | 292 | 299 | 279 | 294 | 535 | 271 | 271 | 317 | 274 | 313 | 610 | 610 | 129 |
| J | 820 | 743 | 693 | 690 | 368 | 325 | 359 | 372 | 296 | 304 | 642 | 432 | 299 | 284 | 594 | 279 | 285 | 726 | 260 | 296 | 617 | 540 | 688 | 127 |
| K | 430 | 801 | 737 | 705 | 453 | 676 | 846 | 346 | 646 | 357 | 367 | 317 | 380 | 308 | 439 | 320 | 339 | 321 | 320 | 341 | 321 | 659 | 417 | 119 |
| L | 400 | 694 | 676 | 725 | 651 | 662 | 330 | 344 | 310 | 371 | 651 | 468 | 600 | 294 | 302 | 290 | 581 | 344 | 797 | 308 | 607 | 296 | 682 | 88 |
| M | 439 | 838 | 647 | 850 | 442 | 719 | 372 | 688 | 357 | 695 | 330 | 697 | 687 | 697 | 365 | 363 | 355 | 652 | 350 | 413 | 368 | 635 | 627 | 118 |
| N | 817 | 782 | 929 | 773 | 827 | 744 | 762 | 641 | 646 | 516 | 734 | 443 | 666 | 401 | 699 | 668 | 679 | 701 | 376 | 667 | 725 | 668 | 750 | 125 |
| O | 339 | 625 | 410 | 742 | 359 | 669 | 392 | 578 | 383 | 643 | 345 | 560 | 276 | 563 | 305 | 271 | 309 | 616 | 279 | 534 | 248 | 577 | 288 | 134 |
| P | 238 | 667 | 646 | 443 | 585 | 618 | 663 | 607 | 679 | 690 | 504 | 279 | 544 | 245 | 514 | 253 | 293 | 236 | 281 | 199 | 507 | 431 | 488 | 117 |

### Diverset Plate 2

**DAPI (Cell Conter)**

| Row L | 01 | 02 | 03 | 04 | 05 | 06 | 07 | 08 | 09 | 10 | 11 | 12 | 13 | 14 | 15 | 16 | 17 | 18 | 19 | 20 | 21 | 22 | 23 | 24 |
|---|---|---|---|---|---|---|---|---|---|---|---|---|---|---|---|---|---|---|---|---|---|---|---|---|
| A | 393 | 1144 | 1267 | 2453 | 3715 | 4911 | 3328 | 3793 | 2208 | 2007 | 1540 | 2789 | 1871 | 2613 | 2226 | 2924 | 3432 | 4407 | 3557 | 5127 | 3384 | 3289 | 4132 | 1532 |
| B | 2129 | 3850 | 1784 | 3428 | 4945 | 2970 | 4255 | 3710 | 4522 | 3527 | 4653 | 3795 | 3590 | 4863 | 3607 | 4669 | 4478 | 4697 | 5192 | 4391 | 4039 | 3882 | 3880 | 2485 |
| C | 1737 | 607 | 319 | 1986 | 2710 | 2188 | 3175 | 2412 | 3390 | 3499 | 3252 | 3730 | 4158 | 4021 | 3587 | 3218 | 3096 | 3759 | 2714 | 2598 | 3083 | 2907 | 2316 | 2411 |
| D | 1146 | 901 | 1483 | 1682 | 2421 | 1937 | 2829 | 2560 | 2505 | 3010 | 2673 | 2785 | 2652 | 10 | 3391 | 3338 | 3547 | 3083 | 3092 | 2887 | 2961 | 2687 | 2303 | 2210 |
| E | 1875 | 502 | 741 | 2833 | 3192 | 3052 | 3381 | 3014 | 3613 | 3873 | 3969 | 4118 | 4117 | 4122 | 4536 | 3009 | 3848 | 4011 | 3695 | 4384 | 3658 | 3699 | 3494 | 3246 |
| F | 2817 | 2079 | 1883 | 2465 | 3594 | 4063 | 3206 | 3441 | 4377 | 3802 | 3450 | 2876 | 3508 | 4140 | 3532 | 2027 | 2779 | 4078 | 3204 | 3420 | 2880 | 4051 | 3655 | 2835 |
| G | 9 | 5 | 45 | 474 | 765 | 610 | 1130 | 861 | 1258 | 2000 | 1089 | 1840 | 1581 | 1809 | 1191 | 1379 | 43 | 2141 | 2181 | 1547 | 2402 | 2433 | 1628 | 342 |
| H | 179 | 113 | 390 | 537 | 848 | 608 | 1455 | 1278 | 1361 | 1462 | 1274 | 1608 | 898 | 1306 | 1529 | 1912 | 1943 | 1562 | 2111 | 761 | 1463 | 1819 | 1508 | 88 |
| I | 1177 | 470 | 195 | 508 | 1934 | 793 | 1014 | 1627 | 1773 | 2652 | 2047 | 82 | 1819 | 2324 | 2590 | 2165 | 1238 | 2283 | 1239 | 2637 | 2746 | 2883 | 3075 | 2343 |
| J | 1930 | 447 | 1393 | 1363 | 1378 | 1849 | 1278 | 2682 | 1981 | 2191 | 2028 | 1966 | 1954 | 1997 | 2142 | 2079 | 2477 | 2112 | 2475 | 1887 | 3038 | 2846 | 2815 | 3233 |
| K | 1079 | 133 | 426 | 2866 | 1195 | 909 | 1914 | 2296 | 2603 | 5 | 2312 | 97 | 2464 | 1950 | 1799 | 2154 | 169 | 2063 | 2236 | 2151 | 2344 | 2334 | 2096 | 3239 |
| L | 1456 | 754 | 934 | 748 | 1131 | 1168 | 1008 | 1012 | 1545 | 1506 | 1165 | 1686 | 1429 | 1787 | 1736 | 1796 | 2007 | 2253 | 129 | 1749 | 2265 | 1968 | 1615 | 1775 |
| M | 1511 | 103 | 147 | 563 | 1146 | 417 | 717 | 941 | 1616 | 9 | 2195 | 896 | 1521 | 1446 | 1657 | 2369 | 1781 | 1779 | 1024 | 1575 | 1352 | 1352 | 1614 | 279 |
| N | 3067 | 950 | 344 | 1275 | 1124 | 1066 | 1234 | 2986 | 1323 | 895 | 1898 | 2447 | 1287 | 1161 | 1572 | 2442 | 1249 | 1381 | 393 | 1059 | 2024 | 1125 | 1814 | 44 |
| O | 29 | 483 | 498 | 570 | 391 | 199 | 465 | 128 | 306 | 554 | 541 | 588 | 1380 | 775 | 832 | 1473 | 1269 | 728 | 1190 | 531 | 2202 | 778 | 1882 | 1260 |
| P | 1387 | 957 | 1105 | 2056 | 1561 | 1047 | 1759 | 716 | 974 | 2590 | 1522 | 3085 | 2986 | 2691 | 3074 | 1052 | 2725 | 363 | 2085 | 3038 | 1969 | 2649 | 1466 | 1971 |

**TEXAS RED**

| Row L | 01 | 02 | 03 | 04 | 05 | 06 | 07 | 08 | 09 | 10 | 11 | 12 | 13 | 14 | 15 | 16 | 17 | 18 | 19 | 20 | 21 | 22 | 23 | 24 |
|---|---|---|---|---|---|---|---|---|---|---|---|---|---|---|---|---|---|---|---|---|---|---|---|---|
| A | 467 | 331 | 493 | 271 | 221 | 240 | 248 | 257 | 331 | 327 | 370 | 246 | 355 | 312 | 346 | 336 | 258 | 244 | 276 | 231 | 290 | 282 | 275 | 117 |
| B | 382 | 240 | 432 | 230 | 243 | 251 | 298 | 242 | 247 | 269 | 241 | 246 | 279 | 214 | 280 | 258 | 251 | 243 | 253 | 262 | 267 | 252 | 292 | 115 |
| C | 287 | 453 | 635 | 383 | 327 | 372 | 283 | 325 | 293 | 265 | 291 | 275 | 250 | 258 | 266 | 291 | 255 | 282 | 309 | 324 | 296 | 317 | 423 | 86 |
| D | 490 | 521 | 479 | 390 | 354 | 385 | 327 | 327 | 343 | 311 | 318 | 335 | 342 | 448 | 292 | 296 | 283 | 343 | 303 | 295 | 330 | 288 | 375 | 116 |
| E | 447 | 671 | 514 | 323 | 338 | 306 | 253 | 296 | 260 | 260 | 278 | 269 | 243 | 277 | 261 | 309 | 280 | 288 | 287 | 255 | 253 | 285 | 286 | 117 |
| F | 290 | 382 | 359 | 369 | 298 | 275 | 307 | 304 | 275 | 253 | 320 | 332 | 281 | 282 | 302 | 354 | 319 | 277 | 306 | 321 | 312 | 285 | 305 | 117 |
| G | 475 | 549 | 728 | 574 | 461 | 619 | 494 | 471 | 445 | 364 | 438 | 381 | 399 | 366 | 362 | 434 | 780 | 364 | 360 | 364 | 322 | 306 | 437 | 116 |
| H | 945 | 937 | 607 | 552 | 502 | 517 | 352 | 390 | 448 | 416 | 418 | 412 | 480 | 389 | 445 | 393 | 354 | 345 | 340 | 443 | 357 | 383 | 364 | 117 |
| I | 401 | 821 | 687 | 503 | 340 | 491 | 472 | 354 | 350 | 300 | 351 | 798 | 334 | 319 | 303 | 312 | 420 | 308 | 459 | 305 | 308 | 323 | 270 | 116 |
| J | 352 | 705 | 431 | 349 | 411 | 421 | 532 | 325 | 365 | 369 | 344 | 380 | 397 | 363 | 344 | 299 | 344 | 352 | 335 | 417 | 304 | 335 | 340 | 116 |
| K | 530 | 868 | 688 | 323 | 427 | 582 | 418 | 311 | 380 | 248 | 337 | 965 | 382 | 393 | 419 | 388 | 860 | 363 | 397 | 391 | 382 | 356 | 460 | 116 |
| L | 423 | 647 | 514 | 599 | 476 | 474 | 447 | 508 | 339 | 470 | 542 | 436 | 428 | 442 | 400 | 407 | 463 | 380 | 923 | 417 | 353 | 410 | 422 | 116 |
| M | 323 | 711 | 941 | 713 | 557 | 705 | 535 | 547 | 355 | 334 | 410 | 610 | 378 | 532 | 439 | 389 | 379 | 484 | 506 | 382 | 483 | 475 | 447 | 248 |
| N | 323 | 467 | 671 | 418 | 495 | 563 | 463 | 326 | 473 | 573 | 408 | 400 | 468 | 438 | 405 | 394 | 475 | 423 | 694 | 541 | 403 | 575 | 439 | 118 |
| O | 699 | 417 | 757 | 806 | 789 | 802 | 656 | 665 | 779 | 710 | 744 | 485 | 423 | 530 | 600 | 499 | 470 | 558 | 563 | 671 | 466 | 478 | 414 | 120 |
| P | 408 | 368 | 563 | 392 | 482 | 500 | 378 | 637 | 549 | 369 | 486 | 378 | 356 | 386 | 340 | 700 | 329 | 946 | 401 | 328 | 433 | 402 | 510 | 117 |

### Diverset Plate 3

**DAPI (Cell Conter)**

| ow Lab | 01 | 02 | 03 | 04 | 05 | 06 | 07 | 08 | 09 | 10 | 11 | 12 | 13 | 14 | 15 | 16 | 17 | 18 | 19 | 20 | 21 | 22 | 23 | 24 |
|---|---|---|---|---|---|---|---|---|---|---|---|---|---|---|---|---|---|---|---|---|---|---|---|---|
| A | 1635 | 4017 | 3059 | 3018 | 2616 | 2017 | 2709 | 1735 | 2726 | 3195 | 2832 | 2184 | 2800 | 463 | 1979 | 4026 | 2583 | 2600 | 2354 | 627 | 2807 | 3368 | 2886 | 3672 |
| B | 2583 | 66 | 3892 | 3522 | 2627 | 3744 | 1665 | 3552 | 3548 | 3171 | 2378 | 2812 | 3498 | 3303 | 3896 | 3748 | 3860 | 3953 | 3614 | 3686 | 2822 | 3472 | 4099 | 2532 |
| C | 1529 | 2012 | 1722 | 3084 | 2613 | 1026 | 2357 | 2530 | 3191 | 2702 | 2599 | 2942 | 3175 | 2755 | 2529 | 2619 | 1293 | 3127 | 3234 | 3401 | 3863 | 2929 | 3531 | 3594 |
| D | 2301 | 5 | 2380 | 1827 | 3186 | 2564 | 3047 | 3459 | 2344 | 2994 | 2337 | 3048 | 2913 | 2893 | 2290 | 3166 | 2827 | 2989 | 3479 | 2636 | 2585 | 3162 | 3524 | 2229 |
| E | 1717 | 1556 | 1359 | 3003 | 2385 | 2722 | 2561 | 3461 | 3238 | 2512 | 2997 | 3904 | 3115 | 3140 | 3332 | 3325 | 3012 | 3221 | 2966 | 3098 | 3013 | 3519 | 3410 | 4311 |
| F | 1775 | 2368 | 2615 | 2973 | 2735 | 3081 | 3044 | 2284 | 2705 | 3385 | 2233 | 2727 | 3278 | 2884 | 3636 | 2707 | 3110 | 2683 | 2978 | 3263 | 3113 | 3019 | 3050 | 4124 |
| G | 1119 | 950 | 678 | 26 | 1370 | 2122 | 2158 | 2246 | 1658 | 2377 | 2196 | 1415 | 1042 | 2691 | 1820 | 1884 | 2343 | 2359 | 1143 | 1438 | 2259 | 2164 | 2229 | 1286 |
| H | 1046 | 1495 | 1779 | 2225 | 2008 | 1689 | 2012 | 652 | 2701 | 1720 | 2657 | 2184 | 1647 | 2125 | 1773 | 2249 | 2173 | 2479 | 2014 | 2577 | 1954 | 2076 | 2415 | 2427 |
| I | 1894 | 1349 | 40 | 12 | 1360 | 1098 | 2417 | 2171 | 1529 | 2209 | 2007 | 2111 | 2010 | 2919 | 1802 | 1979 | 2111 | 2261 | 1610 | 1789 | 1970 | 2477 | 2719 | 1707 |
| J | 1612 | 1689 | 862 | 1871 | 1356 | 1887 | 1539 | 1033 | 1514 | 2203 | 300 | 1736 | 1653 | 2070 | 1886 | 1283 | 1536 | 2110 | 1714 | 2586 | 2166 | 2776 | 2029 | 1123 |
| K | 3153 | 2015 | 2822 | 27 | 1713 | 3054 | 2494 | 2470 | 3593 | 3412 | 2030 | 3564 | 3061 | 2704 | 2680 | 3855 | 3427 | 3602 | 3075 | 3515 | 3263 | 3292 | 3992 | 2197 |
| L | 1826 | 1044 | 2096 | 3090 | 3471 | 2599 | 2907 | 3036 | 2299 | 3546 | 2561 | 2489 | 2996 | 2997 | 3339 | 2810 | 3348 | 3255 | 3404 | 3086 | 3414 | 3658 | 4050 | 2101 |
| M | 2 | 960 | 666 | 242 | 956 | 1888 | 1263 | 1328 | 1691 | 2691 | 1707 | 2100 | 2357 | 2033 | 1998 | 2201 | 3140 | 2612 | 2889 | 2894 | 2393 | 2187 | 2514 | 3541 |
| N | 2181 | 2417 | 1450 | 1446 | 1935 | 1762 | 2258 | 2885 | 2174 | 1918 | 1281 | 2455 | 2185 | 2138 | 3091 | 2100 | 1658 | 2149 | 859 | 2499 | 1506 | 3411 | 3264 | 132 |
| O | 257 | 1146 | 1531 | 180 | 1289 | 845 | 1074 | 669 | 1109 | 1570 | 2167 | 2003 | 1803 | 1070 | 1710 | 1227 | 1134 | 1607 | 2062 | 1828 | 1063 | 1416 | 2974 | 1074 |
| P | 1570 | 1216 | 623 | 1412 | 663 | 1006 | 1291 | 1661 | 1846 | 775 | 890 | 771 | 227 | 2770 | 2648 | 1697 | 1242 | 2345 | 1993 | 1994 | 967 | 2716 | 2247 | 740 |

**TEXAS RED**

| ow Lab | 01 | 02 | 03 | 04 | 05 | 06 | 07 | 08 | 09 | 10 | 11 | 12 | 13 | 14 | 15 | 16 | 17 | 18 | 19 | 20 | 21 | 22 | 23 | 24 |
|---|---|---|---|---|---|---|---|---|---|---|---|---|---|---|---|---|---|---|---|---|---|---|---|---|
| A | 263 | 268 | 422 | 361 | 272 | 395 | 334 | 403 | 374 | 267 | 309 | 304 | 296 | 526 | 406 | 262 | 357 | 311 | 312 | 515 | 387 | 304 | 267 | 118 |
| B | 372 | 699 | 299 | 309 | 336 | 304 | 372 | 317 | 295 | 315 | 355 | 326 | 269 | 303 | 275 | 290 | 301 | 291 | 287 | 311 | 326 | 302 | 276 | 117 |
| C | 344 | 372 | 446 | 364 | 287 | 470 | 339 | 364 | 284 | 289 | 363 | 325 | 312 | 345 | 315 | 309 | 467 | 273 | 269 | 278 | 265 | 289 | 293 | 116 |
| D | 322 | 88 | 322 | 404 | 318 | 313 | 326 | 298 | 323 | 317 | 341 | 266 | 321 | 327 | 349 | 301 | 309 | 310 | 305 | 316 | 335 | 313 | 289 | 115 |
| E | 431 | 385 | 420 | 329 | 302 | 255 | 335 | 260 | 320 | 316 | 294 | 295 | 304 | 318 | 298 | 281 | 293 | 313 | 318 | 294 | 286 | 333 | 311 | 117 |
| F | 416 | 365 | 325 | 312 | 325 | 329 | 328 | 383 | 329 | 307 | 377 | 291 | 337 | 351 | 310 | 382 | 335 | 365 | 305 | 319 | 312 | 326 | 327 | 116 |
| G | 440 | 419 | 463 | 917 | 383 | 369 | 362 | 340 | 383 | 278 | 346 | 408 | 406 | 314 | 334 | 323 | 297 | 361 | 384 | 413 | 324 | 326 | 326 | 115 |
| H | 469 | 457 | 353 | 354 | 347 | 399 | 390 | 506 | 326 | 382 | 343 | 403 | 425 | 354 | 447 | 354 | 317 | 351 | 369 | 313 | 387 | 345 | 347 | 114 |
| I | 321 | 488 | 659 | 698 | 417 | 505 | 355 | 284 | 437 | 323 | 380 | 341 | 336 | 303 | 364 | 335 | 329 | 323 | 359 | 335 | 329 | 350 | 329 | 119 |
| J | 457 | 392 | 580 | 425 | 449 | 361 | 387 | 456 | 411 | 390 | 738 | 403 | 392 | 361 | 414 | 490 | 416 | 343 | 384 | 340 | 386 | 296 | 408 | 118 |
| K | 348 | 368 | 326 | 1290 | 365 | 324 | 382 | 335 | 322 | 309 | 384 | 303 | 335 | 347 | 310 | 276 | 317 | 314 | 316 | 327 | 344 | 323 | 321 | 115 |
| L | 433 | 542 | 378 | 383 | 353 | 381 | 312 | 354 | 359 | 332 | 382 | 395 | 329 | 350 | 324 | 345 | 318 | 332 | 327 | 316 | 323 | 325 | 318 | 115 |
| M | 92 | 502 | 663 | 866 | 452 | 406 | 535 | 453 | 412 | 380 | 442 | 389 | 383 | 428 | 365 | 450 | 348 | 359 | 358 | 334 | 385 | 369 | 389 | 118 |
| N | 397 | 380 | 426 | 475 | 452 | 442 | 407 | 380 | 358 | 426 | 495 | 379 | 416 | 383 | 341 | 412 | 463 | 400 | 689 | 366 | 394 | 305 | 329 | 117 |
| O | 669 | 532 | 497 | 814 | 435 | 555 | 461 | 546 | 473 | 453 | 360 | 319 | 411 | 488 | 487 | 445 | 513 | 410 | 366 | 355 | 512 | 482 | 329 | 117 |
| P | 342 | 554 | 562 | 432 | 558 | 470 | 485 | 380 | 326 | 671 | 567 | 505 | 968 | 371 | 316 | 415 | 409 | 363 | 377 | 388 | 485 | 323 | 384 | 117 |

### Diverset Plate 4

**DAPI (Cell Conter)**

| Rótulo | 01 | 02 | 03 | 04 | 05 | 06 | 07 | 08 | 09 | 10 | 11 | 12 | 13 | 14 | 15 | 16 | 17 | 18 | 19 | 20 | 21 | 22 | 23 | 24 |
|---|---|---|---|---|---|---|---|---|---|---|---|---|---|---|---|---|---|---|---|---|---|---|---|---|
| A | 2099 | 3109 | 2514 | 3694 | 3832 | 3676 | 3578 | 3827 | 3021 | 3141 | 2068 | 341 | 411 | 2637 | 2414 | 1403 | 3002 | 2414 | 968 | 1917 | 2420 | 2510 | 1682 | 1500 |
| B | 2021 | 1533 | 1605 | 1817 | 1794 | 12 | 2656 | 1491 | 2281 | 2132 | 1664 | 9 | 2071 | 308 | 2232 | 1857 | 2443 | 2305 | 1950 | 2354 | 1617 | 1801 | 1414 | 3027 |
| C | 872 | 554 | 1976 | 1915 | 2486 | 1479 | 1582 | 2763 | 2848 | 2105 | 2445 | 1736 | 88 | 147 | 110 | 2608 | 2226 | 3338 | 530 | 2270 | 2709 | 1447 | 2201 | 2416 |
| D | 3856 | 1928 | 2629 | 17 | 1133 | 2129 | 2850 | 3233 | 3548 | 2916 | 2 | 2657 | 3362 | 129 | 1581 | 2554 | 2845 | 2175 | 1479 | 1631 | 3188 | 2628 | 2000 | 1103 |
| E | 2870 | 3042 | 1710 | 2149 | 1951 | 2926 | 2611 | 2850 | 1941 | 2519 | 1051 | 1710 | 1995 | 1743 | 303 | 2048 | 1413 | 1917 | 261 | 1660 | 1102 | 1062 | 884 | 1199 |
| F | 1354 | 7 | 2518 | 12 | 2626 | 2545 | 2864 | 2771 | 2773 | 2227 | 2376 | 2721 | 2521 | 2481 | 2391 | 2254 | 2324 | 1441 | 1805 | 1953 | 2133 | 1792 | 1576 | 2084 |
| G | 1662 | 1847 | 1239 | 1294 | 1700 | 920 | 1918 | 1741 | 1353 | 1731 | 1510 | 1807 | 1790 | 1894 | 509 | 1687 | 2090 | 1466 | 1518 | 1804 | 1723 | 1651 | 1822 | 1696 |
| H | 1326 | 1588 | 1811 | 1432 | 1283 | 1794 | 1370 | 1624 | 1424 | 1124 | 15 | 1040 | 1833 | 1499 | 1914 | 2090 | 1983 | 1763 | 1202 | 1449 | 1170 | 1389 | 1704 | 1881 |
| I | 1185 | 2587 | 2306 | 521 | 2926 | 3568 | 2301 | 2855 | 3113 | 2942 | 1828 | 2954 | 1357 | 2340 | 266 | 2653 | 2715 | 2411 | 2250 | 2203 | 1886 | 2542 | 2255 | 1938 |
| J | 1572 | 2448 | 2982 | 2192 | 2157 | 2680 | 63 | 2510 | 839 | 2403 | 2 | 1807 | 2571 | 2266 | 2260 | 1803 | 3263 | 2137 | 224 | 2385 | 2601 | 2188 | 2648 | 3375 |
| K | 1600 | 1437 | 1588 | 2551 | 1540 | 2108 | 180 | 1901 | 2225 | 2296 | 2265 | 2076 | 336 | 2334 | 260 | 2219 | 2305 | 1605 | 1390 | 1675 | 1192 | 1556 | 815 | 1328 |
| L | 3200 | 2848 | 515 | 2827 | 986 | 2865 | 2659 | 2592 | 1917 | 2728 | 6 | 30 | 3019 | 1562 | 1490 | 1415 | 2127 | 2027 | 2294 | 1079 | 1167 | 1807 | 2450 | 1081 |
| M | 1681 | 2016 | 1277 | 1540 | 1368 | 763 | 1689 | 2127 | 1585 | 1819 | 1520 | 1249 | 80 | 1068 | 1336 | 1401 | 1468 | 1722 | 1412 | 1018 | 1731 | 2419 | 1552 | 1088 |
| N | 3214 | 1002 | 2004 | 1724 | 1233 | 3033 | 2689 | 1875 | 1941 | 1290 | 1907 | 1411 | 2163 | 1738 | 1890 | 1806 | 1681 | 1078 | 1526 | 1634 | 1612 | 1683 | 1381 | 387 |
| O | 2201 | 447 | 555 | 2824 | 2237 | 2321 | 2168 | 2045 | 2424 | 2005 | 1743 | 2026 | 593 | 1461 | 1200 | 1954 | 1118 | 961 | 1655 | 1835 | 1354 | 1945 | 1385 | 1388 |
| P | 593 | 1456 | 1279 | 1557 | 1765 | 2143 | 2606 | 1504 | 2579 | 1001 | 2245 | 1381 | 1759 | 1740 | 2908 | 3661 | 3128 | 2452 | 879 | 2046 | 1546 | 2026 | 1813 | 1530 |

**TEXAS RED**

| Rótulo | 01 | 02 | 03 | 04 | 05 | 06 | 07 | 08 | 09 | 10 | 11 | 12 | 13 | 14 | 15 | 16 | 17 | 18 | 19 | 20 | 21 | 22 | 23 | 24 |
|---|---|---|---|---|---|---|---|---|---|---|---|---|---|---|---|---|---|---|---|---|---|---|---|---|
| A | 161 | 263 | 234 | 230 | 243 | 227 | 196 | 253 | 276 | 177 | 337 | 749 | 721 | 333 | 313 | 518 | 274 | 293 | 575 | 272 | 254 | 377 | 322 | 119 |
| B | 366 | 590 | 570 | 536 | 380 | 612 | 341 | 603 | 374 | 373 | 469 | 348 | 502 | 763 | 392 | 484 | 346 | 387 | 365 | 401 | 376 | 537 | 621 | 117 |
| C | 411 | 590 | 319 | 354 | 359 | 372 | 476 | 382 | 318 | 406 | 349 | 384 | 1078 | 955 | 1017 | 333 | 383 | 286 | 712 | 357 | 336 | 573 | 335 | 118 |
| D | 294 | 483 | 419 | 958 | 598 | 471 | 365 | 353 | 349 | 400 | 129 | 390 | 308 | 995 | 548 | 370 | 421 | 398 | 525 | 538 | 333 | 414 | 489 | 124 |
| E | 227 | 254 | 290 | 316 | 294 | 216 | 241 | 223 | 233 | 219 | 332 | 246 | 237 | 271 | 844 | 220 | 311 | 231 | 618 | 281 | 284 | 305 | 417 | 113 |
| F | 407 | 357 | 307 | 489 | 247 | 311 | 241 | 252 | 277 | 287 | 264 | 260 | 266 | 256 | 278 | 266 | 304 | 372 | 284 | 309 | 347 | 335 | 343 | 116 |
| G | 531 | 296 | 403 | 408 | 358 | 548 | 383 | 384 | 481 | 418 | 396 | 340 | 449 | 446 | 746 | 477 | 495 | 390 | 442 | 492 | 449 | 595 | 458 | 126 |
| H | 424 | 442 | 446 | 412 | 548 | 500 | 572 | 488 | 425 | 706 | 581 | 611 | 496 | 460 | 475 | 453 | 405 | 451 | 681 | 557 | 692 | 650 | 409 | 122 |
| I | 457 | 412 | 311 | 841 | 370 | 390 | 483 | 472 | 435 | 441 | 491 | 378 | 567 | 401 | 1044 | 464 | 431 | 438 | 504 | 429 | 447 | 430 | 436 | 119 |
| J | 485 | 451 | 409 | 487 | 483 | 477 | 1067 | 434 | 802 | 446 | 507 | 493 | 436 | 490 | 519 | 500 | 437 | 512 | 1067 | 487 | 447 | 463 | 428 | 122 |
| K | 405 | 508 | 525 | 420 | 611 | 492 | 937 | 508 | 528 | 440 | 491 | 530 | 843 | 413 | 1067 | 437 | 468 | 453 | 480 | 556 | 659 | 536 | 700 | 123 |
| L | 428 | 509 | 832 | 443 | 657 | 465 | 522 | 508 | 548 | 488 | 165 | 1470 | 456 | 552 | 511 | 640 | 452 | 487 | 584 | 639 | 534 | 521 | 385 | 127 |
| M | 544 | 510 | 614 | 497 | 627 | 818 | 402 | 454 | 477 | 548 | 615 | 448 | 1320 | 714 | 616 | 514 | 417 | 373 | 599 | 735 | 565 | 538 | 561 | 122 |
| N | 297 | 420 | 460 | 577 | 658 | 369 | 526 | 581 | 418 | 679 | 450 | 614 | 451 | 463 | 598 | 451 | 382 | 581 | 534 | 468 | 529 | 394 | 483 | 129 |
| O | 327 | 795 | 910 | 531 | 439 | 349 | 488 | 522 | 523 | 550 | 639 | 438 | 783 | 555 | 567 | 574 | 626 | 727 | 495 | 589 | 642 | 477 | 633 | 127 |
| P | 720 | 417 | 557 | 567 | 520 | 387 | 415 | 594 | 391 | 760 | 404 | 579 | 526 | 658 | 459 | 336 | 367 | 416 | 728 | 517 | 427 | 489 | 426 | 123 |

### Diverset Plate 5

**DAPI (Cell Conter)**

| Row La | 01 | 02 | 03 | 04 | 05 | 06 | 07 | 08 | 09 | 10 | 11 | 12 | 13 | 14 | 15 | 16 | 17 | 18 | 19 | 20 | 21 | 22 | 23 | 24 |
|---|---|---|---|---|---|---|---|---|---|---|---|---|---|---|---|---|---|---|---|---|---|---|---|---|
| A | 597 | 1057 | 2380 | 2764 | 1441 | 1595 | 7 | 2190 | 3063 | 2051 | 2795 | 1787 | 110 | 2856 | 2514 | 679 | 1643 | 2041 | 1307 | 1930 | 844 | 1836 | 1987 | 3016 |
| B | 1210 | 1038 | 1128 | 1767 | 2606 | 1806 | 2311 | 2070 | 1827 | 2856 | 2676 | 1571 | 854 | 1407 | 999 | 1247 | 1741 | 1958 | 2601 | 2753 | 2322 | 1248 | 3035 | 3570 |
| C | 1237 | 319 | 1065 | 1470 | 1666 | 2070 | 1333 | 1296 | 667 | 1697 | 1697 | 896 | 1863 | 1796 | 449 | 1085 | 1174 | 1769 | 2400 | 2452 | 2492 | 1236 | 2941 | 2855 |
| D | 1644 | 943 | 1238 | 448 | 1700 | 1646 | 194 | 924 | 2240 | 1198 | 2070 | 1444 | 1944 | 1371 | 1892 | 1759 | 2265 | 2265 | 2061 | 2971 | 2599 | 3590 | 3097 | 2320 |
| E | 1952 | 1457 | 1082 | 2103 | 2120 | 1581 | 1704 | 1194 | 1179 | 1983 | 1734 | 1361 | 1518 | 2287 | 2289 | 2274 | 2155 | 2009 | 1789 | 2725 | 2487 | 2734 | 2237 | 1984 |
| F | 1248 | 569 | 1774 | 1300 | 362 | 1376 | 1557 | 1558 | 1790 | 1646 | 1293 | 1913 | 2050 | 2086 | 1709 | 1960 | 2061 | 2050 | 1953 | 2367 | 2415 | 1923 | 2536 | 949 |
| G | 297 | 19 | 190 | 519 | 537 | 520 | 519 | 627 | 201 | 508 | 1329 | 993 | 977 | 1734 | 1214 | 1247 | 1582 | 2182 | 1208 | 674 | 2045 | 2770 | 2408 | 1730 |
| H | 458 | 197 | 356 | 659 | 1026 | 590 | 941 | 1276 | 732 | 922 | 1296 | 342 | 857 | 1075 | 308 | 338 | 1137 | 1618 | 1625 | 2214 | 1763 | 1974 | 2482 | 308 |
| I | 427 | 496 | 445 | 878 | 561 | 814 | 1023 | 962 | 926 | 986 | 1134 | 862 | 630 | 942 | 1169 | 1899 | 1342 | 1070 | 1170 | 2104 | 2013 | 1534 | 2785 | 3449 |
| J | 306 | 1222 | 705 | 281 | 23 | 544 | 1091 | 741 | 661 | 991 | 381 | 883 | 957 | 1435 | 874 | 1482 | 1144 | 1273 | 1494 | 1353 | 1730 | 1172 | 2233 | 1895 |
| K | 174 | 67 | 149 | 596 | 387 | 366 | 578 | 892 | 770 | 791 | 315 | 119 | 1460 | 1421 | 619 | 547 | 617 | 600 | 1092 | 898 | 1875 | 1121 | 957 | 616 |
| L | 17 | 85 | 61 | 634 | 147 | 600 | 484 | 430 | 258 | 676 | 61 | 246 | 746 | 621 | 551 | 697 | 1651 | 977 | 843 | 698 | 223 | 1079 | 1803 | 1480 |
| M | 137 | 54 | 272 | 505 | 387 | 801 | 499 | 809 | 900 | 768 | 547 | 602 | 1481 | 1384 | 1199 | 1905 | 940 | 1239 | 1663 | 1262 | 2029 | 1667 | 2492 | 1948 |
| N | 67 | 65 | 496 | 293 | 236 | 451 | 306 | 505 | 454 | 479 | 320 | 963 | 1490 | 1012 | 848 | 1419 | 1658 | 534 | 1562 | 1642 | 1957 | 1462 | 1623 | 254 |
| O | 114 | 128 | 101 | 273 | 240 | 283 | 146 | 214 | 107 | 217 | 195 | 293 | 714 | 432 | 571 | 503 | 1169 | 358 | 419 | 1046 | 141 | 724 | 1351 | 477 |
| P | 440 | 1929 | 467 | 492 | 387 | 1969 | 6 | 291 | 349 | 352 | 162 | 382 | 840 | 198 | 2687 | 433 | 402 | 2513 | 847 | 2301 | 673 | 517 | 1196 | 758 |

**TEXAS RED**

| Row La | 01 | 02 | 03 | 04 | 05 | 06 | 07 | 08 | 09 | 10 | 11 | 12 | 13 | 14 | 15 | 16 | 17 | 18 | 19 | 20 | 21 | 22 | 23 | 24 |
|---|---|---|---|---|---|---|---|---|---|---|---|---|---|---|---|---|---|---|---|---|---|---|---|---|
| A | 903 | 351 | 379 | 278 | 741 | 440 | 776 | 472 | 189 | 567 | 344 | 472 | 1220 | 265 | 442 | 608 | 527 | 377 | 504 | 445 | 761 | 196 | 488 | 121 |
| B | 389 | 352 | 526 | 713 | 403 | 395 | 387 | 448 | 486 | 311 | 340 | 578 | 655 | 445 | 740 | 542 | 451 | 481 | 421 | 445 | 390 | 730 | 316 | 123 |
| C | 398 | 840 | 562 | 548 | 554 | 358 | 768 | 571 | 774 | 513 | 506 | 565 | 356 | 442 | 704 | 541 | 497 | 419 | 326 | 303 | 420 | 470 | 433 | 125 |
| D | 373 | 661 | 492 | 985 | 696 | 615 | 1176 | 422 | 487 | 696 | 441 | 549 | 456 | 661 | 384 | 433 | 374 | 366 | 407 | 404 | 395 | 340 | 259 | 121 |
| E | 436 | 346 | 512 | 340 | 280 | 282 | 314 | 397 | 430 | 245 | 276 | 386 | 284 | 274 | 234 | 273 | 264 | 272 | 271 | 214 | 236 | 269 | 267 | 114 |
| F | 380 | 533 | 370 | 511 | 622 | 441 | 340 | 337 | 259 | 322 | 357 | 350 | 293 | 252 | 305 | 281 | 304 | 251 | 254 | 246 | 300 | 311 | 263 | 114 |
| G | 718 | 882 | 780 | 558 | 462 | 552 | 469 | 519 | 997 | 602 | 420 | 404 | 358 | 400 | 356 | 492 | 281 | 266 | 478 | 632 | 484 | 365 | 521 | 128 |
| H | 778 | 1179 | 1018 | 536 | 489 | 786 | 443 | 428 | 833 | 414 | 564 | 776 | 600 | 436 | 817 | 805 | 589 | 558 | 467 | 552 | 507 | 369 | 449 | 176 |
| I | 973 | 704 | 850 | 557 | 664 | 731 | 466 | 691 | 760 | 516 | 548 | 616 | 789 | 659 | 522 | 513 | 528 | 546 | 457 | 504 | 465 | 570 | 475 | 125 |
| J | 1174 | 512 | 629 | 992 | 1200 | 661 | 558 | 542 | 698 | 479 | 809 | 739 | 742 | 604 | 629 | 572 | 640 | 678 | 471 | 574 | 703 | 651 | 474 | 131 |
| K | 1197 | 1741 | 1549 | 536 | 788 | 903 | 387 | 529 | 668 | 517 | 922 | 1135 | 527 | 625 | 613 | 543 | 833 | 643 | 583 | 659 | 519 | 795 | 638 | 95 |
| L | 1461 | 1348 | 1026 | 891 | 1397 | 650 | 767 | 742 | 1088 | 658 | 1479 | 723 | 703 | 821 | 691 | 640 | 589 | 587 | 743 | 818 | 1248 | 935 | 498 | 132 |
| M | 1090 | 1552 | 964 | 921 | 874 | 779 | 889 | 496 | 607 | 716 | 618 | 552 | 629 | 518 | 588 | 588 | 715 | 709 | 704 | 570 | 740 | 638 | 556 | 126 |
| N | 1020 | 1188 | 585 | 1214 | 965 | 741 | 988 | 810 | 1035 | 960 | 892 | 703 | 560 | 792 | 784 | 582 | 393 | 1043 | 633 | 493 | 541 | 473 | 503 | 101 |
| O | 761 | 1381 | 882 | 869 | 942 | 960 | 1321 | 1200 | 1354 | 908 | 1064 | 812 | 947 | 1042 | 736 | 618 | 617 | 721 | 915 | 616 | 955 | 578 | 629 | 136 |
| P | 590 | 446 | 1092 | 855 | 685 | 454 | 1308 | 977 | 793 | 821 | 1372 | 830 | 657 | 1229 | 338 | 646 | 804 | 442 | 698 | 442 | 694 | 558 | 435 | 133 |

### Diverset Plate 6

**DAPI (Cell Conter)**

| Row La | 01 | 02 | 03 | 04 | 05 | 06 | 07 | 08 | 09 | 10 | 11 | 12 | 13 | 14 | 15 | 16 | 17 | 18 | 19 | 20 | 21 | 22 | 23 | 24 |
|---|---|---|---|---|---|---|---|---|---|---|---|---|---|---|---|---|---|---|---|---|---|---|---|---|
| A | 551 | 610 | 804 | 1663 | 522 | 322 | 1107 | 905 | 2116 | 250 | 688 | 1222 | 574 | 1378 | 873 | 2849 | 1629 | 2041 | 2636 | 1479 | 559 | 4 | 1200 | 3252 |
| B | 619 | 1462 | 2014 | 1868 | 443 | 1455 | 2427 | 1228 | 1438 | 1375 | 1661 | 2698 | 2953 | 2575 | 1556 | 1908 | 2336 | 3111 | 2964 | 2221 | 1545 | 2864 | 2074 | 1895 |
| C | 672 | 1122 | 1152 | 1022 | 966 | 1147 | 1817 | 591 | 1018 | 4 | 1736 | 1790 | 1653 | 2160 | 2026 | 2147 | 2028 | 1693 | 616 | 92 | 2144 | 1948 | 2668 | 2113 |
| D | 1413 | 750 | 1107 | 1518 | 1507 | 1988 | 864 | 1406 | 2134 | 1509 | 1027 | 2448 | 2842 | 2240 | 1450 | 2397 | 2672 | 1953 | 2235 | 2198 | 2407 | 2722 | 3033 | 3802 |
| E | 245 | 1154 | 533 | 1700 | 1200 | 392 | 1836 | 1659 | 1871 | 1763 | 1689 | 1503 | 1679 | 1753 | 1599 | 2140 | 1538 | 2246 | 2074 | 1928 | 2187 | 1036 | 2474 | 2231 |
| F | 1369 | 1106 | 1103 | 1386 | 1909 | 1742 | 2082 | 2274 | 2076 | 2780 | 2134 | 2005 | 2214 | 1924 | 2188 | 1835 | 1547 | 1634 | 1848 | 2494 | 2802 | 2437 | 2906 | 4045 |
| G | 998 | 373 | 276 | 478 | 1310 | 656 | 1162 | 1292 | 1572 | 585 | 954 | 1008 | 1158 | 946 | 1141 | 691 | 1659 | 1608 | 2027 | 1634 | 1437 | 1086 | 2185 | 1975 |
| H | 119 | 351 | 156 | 357 | 602 | 484 | 713 | 792 | 581 | 832 | 677 | 1259 | 1219 | 1674 | 872 | 1058 | 810 | 1007 | 923 | 1277 | 1104 | 1093 | 2155 | 983 |
| I | 223 | 210 | 140 | 272 | 546 | 434 | 1012 | 945 | 797 | 1114 | 954 | 1156 | 1205 | 1136 | 1690 | 907 | 1398 | 142 | 1444 | 1536 | 1365 | 1392 | 1955 | 2078 |
| J | 165 | 353 | 281 | 211 | 406 | 746 | 424 | 908 | 803 | 752 | 895 | 619 | 968 | 1097 | 973 | 1386 | 2170 | 1182 | 1221 | 1168 | 1305 | 86 | 2112 | 3288 |
| K | 49 | 41 | 39 | 213 | 205 | 762 | 150 | 994 | 436 | 523 | 533 | 202 | 478 | 720 | 210 | 229 | 341 | 259 | 468 | 518 | 545 | 841 | 651 | 229 |
| L | 9 | 39 | 240 | 268 | 703 | 261 | 312 | 387 | 970 | 597 | 587 | 598 | 587 | 1254 | 910 | 714 | 511 | 1147 | 556 | 730 | 306 | 1190 | 886 | 1288 |
| M | 75 | 37 | 339 | 296 | 429 | 580 | 455 | 332 | 597 | 728 | 583 | 1236 | 703 | 960 | 930 | 670 | 1246 | 825 | 1595 | 1963 | 1669 | 1059 | 1341 | 1363 |
| N | 101 | 178 | 215 | 425 | 164 | 374 | 401 | 189 | 757 | 736 | 574 | 439 | 414 | 590 | 507 | 409 | 353 | 586 | 614 | 864 | 637 | 403 | 1227 | 101 |
| O | 80 | 38 | 49 | 393 | 216 | 130 | 118 | 308 | 75 | 258 | 54 | 296 | 321 | 229 | 179 | 401 | 259 | 178 | 308 | 175 | 248 | 423 | 556 | 803 |
| P | 519 | 614 | 144 | 207 | 799 | 401 | 146 | 693 | 271 | 75 | 239 | 463 | 224 | 194 | 113 | 103 | 60 | 52 | 64 | 272 | 74 | 225 | 237 | 373 |

**TEXAS RED**

| Row La | 01 | 02 | 03 | 04 | 05 | 06 | 07 | 08 | 09 | 10 | 11 | 12 | 13 | 14 | 15 | 16 | 17 | 18 | 19 | 20 | 21 | 22 | 23 | 24 |
|---|---|---|---|---|---|---|---|---|---|---|---|---|---|---|---|---|---|---|---|---|---|---|---|---|
| A | 245 | 396 | 278 | 396 | 497 | 799 | 501 | 785 | 295 | 782 | 788 | 602 | 1006 | 801 | 931 | 250 | 350 | 380 | 284 | 472 | 747 | 1084 | 673 | 126 |
| B | 622 | 448 | 539 | 343 | 766 | 368 | 541 | 501 | 803 | 519 | 374 | 287 | 279 | 367 | 437 | 551 | 355 | 438 | 408 | 361 | 569 | 299 | 365 | 138 |
| C | 550 | 529 | 550 | 698 | 726 | 363 | 406 | 831 | 506 | 688 | 554 | 512 | 390 | 313 | 263 | 381 | 357 | 759 | 626 | 1360 | 550 | 503 | 368 | 122 |
| D | 404 | 644 | 528 | 412 | 341 | 399 | 562 | 667 | 512 | 454 | 541 | 482 | 339 | 395 | 649 | 349 | 322 | 445 | 435 | 431 | 395 | 355 | 408 | 125 |
| E | 564 | 334 | 518 | 288 | 318 | 803 | 303 | 308 | 276 | 295 | 299 | 331 | 324 | 324 | 419 | 269 | 308 | 351 | 310 | 311 | 286 | 385 | 260 | 115 |
| F | 457 | 645 | 406 | 445 | 333 | 362 | 354 | 309 | 316 | 268 | 312 | 317 | 327 | 308 | 289 | 372 | 341 | 318 | 401 | 292 | 235 | 316 | 264 | 116 |
| G | 603 | 757 | 851 | 821 | 425 | 841 | 753 | 508 | 510 | 641 | 540 | 400 | 594 | 488 | 547 | 734 | 405 | 473 | 635 | 646 | 671 | 538 | 494 | 120 |
| H | 827 | 518 | 814 | 763 | 922 | 798 | 737 | 658 | 756 | 419 | 731 | 699 | 505 | 583 | 545 | 498 | 817 | 862 | 526 | 504 | 757 | 573 | 410 | 124 |
| I | 724 | 1084 | 1195 | 1275 | 1058 | 767 | 620 | 833 | 732 | 603 | 598 | 414 | 696 | 521 | 511 | 720 | 651 | 1091 | 498 | 684 | 629 | 712 | 507 | 130 |
| J | 1165 | 915 | 996 | 936 | 847 | 806 | 1231 | 676 | 727 | 536 | 657 | 710 | 696 | 572 | 626 | 656 | 567 | 525 | 602 | 867 | 539 | 1413 | 470 | 127 |
| K | 1146 | 1293 | 1509 | 1031 | 1096 | 697 | 1336 | 546 | 548 | 797 | 900 | 1007 | 636 | 651 | 1355 | 1052 | 866 | 1028 | 866 | 802 | 616 | 749 | 739 | 95 |
| L | 436 | 878 | 891 | 1035 | 839 | 1070 | 913 | 761 | 655 | 654 | 634 | 581 | 725 | 445 | 681 | 741 | 852 | 630 | 737 | 669 | 785 | 586 | 808 | 124 |
| M | 942 | 927 | 641 | 888 | 962 | 761 | 870 | 869 | 882 | 525 | 776 | 720 | 1043 | 723 | 554 | 874 | 521 | 676 | 726 | 623 | 670 | 495 | 715 | 130 |
| N | 1384 | 1258 | 1184 | 944 | 1419 | 857 | 829 | 1092 | 784 | 943 | 636 | 754 | 1002 | 800 | 668 | 926 | 794 | 1062 | 585 | 605 | 885 | 668 | 613 | 137 |
| O | 472 | 1366 | 1465 | 693 | 1177 | 1090 | 1325 | 821 | 1457 | 1132 | 1426 | 809 | 802 | 1407 | 1290 | 1014 | 962 | 1078 | 1113 | 1048 | 967 | 892 | 663 | 133 |
| P | 215 | 555 | 1249 | 757 | 730 | 904 | 1157 | 706 | 865 | 1504 | 929 | 782 | 1012 | 895 | 1099 | 1538 | 1209 | 1024 | 1325 | 1068 | 1129 | 983 | 891 | 98 |
| (blank) | | | | | | | | | | | | | | | | | | | | | | | | |

### Diverset Plate 7

**DAPI (Cell Conter)**

| Row La | 01 | 02 | 03 | 04 | 05 | 06 | 07 | 08 | 09 | 10 | 11 | 12 | 13 | 14 | 15 | 16 | 17 | 18 | 19 | 20 | 21 | 22 | 23 | 24 |
|---|---|---|---|---|---|---|---|---|---|---|---|---|---|---|---|---|---|---|---|---|---|---|---|---|
| A | 19 | 18 | 697 | 786 | 1550 | 2012 | 841 | 1360 | 1030 | 1223 | 817 | 761 | 1877 | 652 | 2016 | 1874 | 2219 | 646 | 1779 | 679 | 2045 | 281 | 2020 | 958 |
| B | 889 | 296 | 645 | 1400 | 2848 | 1869 | 2039 | 1318 | 1138 | 2380 | 1616 | 1478 | 1371 | 2107 | 1150 | 1860 | 1546 | 1083 | 2238 | 2171 | 1363 | 1387 | 3385 | 3121 |
| C | 146 | 215 | 789 | 1047 | 1095 | 1321 | 1541 | 1547 | 1417 | 1534 | 956 | 1891 | 1726 | 141 | 1341 | 1562 | 1036 | 1383 | 1938 | 1281 | 1537 | 1548 | 2315 | 2751 |
| D | 720 | 479 | 496 | 1992 | 1482 | 1120 | 1442 | 851 | 1984 | 1408 | 1095 | 1272 | 1399 | 1181 | 1102 | 1405 | 1241 | 1424 | 1616 | 1861 | 1970 | 2038 | 2151 | 2620 |
| E | 1190 | 439 | 556 | 1372 | 946 | 988 | 1709 | 1637 | 1342 | 1557 | 1200 | 1778 | 1949 | 1708 | 1627 | 1623 | 1452 | 1686 | 1472 | 1333 | 385 | 2108 | 1728 | 2110 |
| F | 1069 | 312 | 674 | 1183 | 1349 | 696 | 1303 | 1331 | 1317 | 1892 | 1505 | 1439 | 2202 | 1212 | 1336 | 1737 | 1629 | 1742 | 1761 | 29 | 1519 | 2338 | 2394 | 2640 |
| G | 537 | 87 | 185 | 489 | 822 | 972 | 964 | 704 | 1305 | 452 | 1039 | 1379 | 1431 | 1259 | 1672 | 1265 | 1313 | 1538 | 1390 | 1552 | 1683 | 1454 | 2350 | 1842 |
| H | 191 | 154 | 328 | 671 | 686 | 834 | 342 | 1087 | 1677 | 1398 | 718 | 998 | 284 | 818 | 1223 | 622 | 1518 | 1137 | 769 | 1141 | 1005 | 1308 | 1652 | 1399 |
| I | 115 | 172 | 231 | 530 | 382 | 619 | 932 | 993 | 1035 | 1244 | 947 | 1080 | 937 | 1213 | 1677 | 1331 | 2215 | 2064 | 1718 | 1770 | 1965 | 1385 | 1944 | 2288 |
| J | 20 | 676 | 85 | 428 | 645 | 637 | 1112 | 1721 | 65 | 827 | 1130 | 953 | 1323 | 1694 | 1438 | 1453 | 1390 | 1295 | 1655 | 1439 | 1372 | 1773 | 2280 | 3251 |
| K | 114 | 66 | 47 | 439 | 638 | 43 | 444 | 772 | 630 | 1923 | 813 | 1552 | 1058 | 985 | 885 | 693 | 1280 | 999 | 1526 | 1160 | 513 | 1670 | 2654 | 406 |
| L | 229 | 48 | 356 | 207 | 139 | 210 | 228 | 22 | 339 | 544 | 471 | 559 | 609 | 107 | 425 | 528 | 607 | 518 | 554 | 726 | 506 | 1033 | 658 | 1009 |
| M | 80 | 152 | 237 | 148 | 649 | 510 | 671 | 574 | 479 | 581 | 842 | 690 | 1289 | 38 | 263 | 1198 | 920 | 618 | 913 | 1319 | 202 | 820 | 525 | 1140 |
| N | 332 | 226 | 33 | 217 | 424 | 369 | 314 | 646 | 734 | 602 | 814 | 786 | 458 | 366 | 732 | 494 | 859 | 676 | 829 | 883 | 970 | 1049 | 842 | 1128 |
| O | 96 | 248 | 197 | 157 | 171 | 197 | 161 | 482 | 325 | 279 | 529 | 161 | 333 | 513 | 299 | 379 | 536 | 32 | 495 | 726 | 381 | 583 | 393 | 1085 |
| P | 558 | 211 | 88 | 1159 | 393 | 235 | 1246 | 334 | 1580 | 651 | 243 | 1460 | 1525 | 413 | 287 | 847 | 82 | 627 | 118 | 285 | 335 | 321 | 595 | 107 |

**TEXAS RED**

| Row La | 01 | 02 | 03 | 04 | 05 | 06 | 07 | 08 | 09 | 10 | 11 | 12 | 13 | 14 | 15 | 16 | 17 | 18 | 19 | 20 | 21 | 22 | 23 | 24 |
|---|---|---|---|---|---|---|---|---|---|---|---|---|---|---|---|---|---|---|---|---|---|---|---|---|
| A | 1161 | 990 | 285 | 1007 | 457 | 368 | 730 | 689 | 536 | 639 | 772 | 643 | 462 | 310 | 474 | 577 | 277 | 202 | 429 | 662 | 332 | 575 | 620 | 94 |
| B | 580 | 949 | 955 | 382 | 328 | 316 | 437 | 559 | 526 | 421 | 551 | 560 | 642 | 421 | 675 | 467 | 482 | 660 | 423 | 365 | 485 | 350 | 477 | 124 |
| C | 1051 | 1126 | 856 | 498 | 623 | 780 | 424 | 405 | 520 | 580 | 994 | 517 | 379 | 1293 | 664 | 592 | 697 | 564 | 557 | 477 | 459 | 660 | 447 | 125 |
| D | 614 | 727 | 711 | 610 | 354 | 477 | 421 | 871 | 416 | 426 | 556 | 570 | 399 | 724 | 545 | 445 | 497 | 449 | 354 | 482 | 634 | 355 | 425 | 124 |
| E | 523 | 609 | 658 | 350 | 334 | 407 | 322 | 322 | 362 | 413 | 347 | 309 | 295 | 305 | 321 | 399 | 381 | 294 | 271 | 376 | 502 | 254 | 309 | 123 |
| F | 456 | 763 | 496 | 424 | 429 | 557 | 399 | 374 | 440 | 327 | 370 | 328 | 302 | 390 | 378 | 315 | 372 | 345 | 361 | 776 | 366 | 320 | 279 | 116 |
| G | 726 | 799 | 750 | 453 | 555 | 480 | 490 | 433 | 606 | 875 | 601 | 417 | 441 | 670 | 282 | 524 | 411 | 507 | 630 | 377 | 526 | 554 | 377 | 136 |
| H | 1155 | 822 | 915 | 516 | 474 | 442 | 985 | 701 | 533 | 390 | 679 | 663 | 1126 | 594 | 348 | 510 | 406 | 454 | 691 | 741 | 581 | 536 | 408 | 130 |
| I | 1233 | 940 | 792 | 902 | 1055 | 901 | 707 | 530 | 513 | 613 | 791 | 581 | 497 | 514 | 567 | 677 | 441 | 480 | 619 | 683 | 527 | 590 | 638 | 127 |
| J | 1621 | 715 | 1330 | 853 | 633 | 924 | 629 | 571 | 1361 | 745 | 468 | 639 | 706 | 569 | 652 | 598 | 590 | 535 | 567 | 651 | 592 | 614 | 630 | 127 |
| K | 1481 | 1350 | 1333 | 924 | 743 | 1488 | 786 | 530 | 887 | 525 | 751 | 630 | 414 | 372 | 676 | 590 | 600 | 695 | 484 | 556 | 718 | 583 | 482 | 136 |
| L | 984 | 1155 | 817 | 839 | 1457 | 905 | 910 | 1067 | 528 | 605 | 979 | 675 | 961 | 1293 | 1041 | 924 | 899 | 433 | 565 | 558 | 926 | 559 | 860 | 132 |
| M | 858 | 1176 | 922 | 1181 | 570 | 870 | 550 | 710 | 644 | 697 | 666 | 765 | 614 | 1474 | 1115 | 440 | 629 | 819 | 512 | 551 | 1132 | 913 | 723 | 134 |
| N | 1037 | 992 | 1464 | 1134 | 907 | 655 | 979 | 905 | 449 | 646 | 952 | 720 | 1057 | 932 | 880 | 839 | 717 | 716 | 809 | 667 | 625 | 965 | 656 | 131 |
| O | 1228 | 988 | 912 | 1182 | 1052 | 1126 | 1547 | 625 | 1165 | 1035 | 968 | 1457 | 1076 | 1032 | 1025 | 866 | 852 | 1453 | 591 | 642 | 715 | 810 | 930 | 135 |
| P | 670 | 764 | 727 | 428 | 931 | 1202 | 754 | 1182 | 485 | 829 | 568 | 659 | 341 | 612 | 904 | 774 | 1702 | 594 | 725 | 759 | 875 | 842 | 681 | 138 |

### Diverset Plate 8

**DAPI (Cell Conter)**

| Row La | 01 | 02 | 03 | 04 | 05 | 06 | 07 | 08 | 09 | 10 | 11 | 12 | 13 | 14 | 15 | 16 | 17 | 18 | 19 | 20 | 21 | 22 | 23 | 24 |
|---|---|---|---|---|---|---|---|---|---|---|---|---|---|---|---|---|---|---|---|---|---|---|---|---|
| A | 24 | 230 | 459 | 845 | 934 | 1537 | 772 | 1720 | 1318 | 1031 | 698 | 1687 | 1509 | 1496 | 1104 | 83 | 2393 | 3373 | 1759 | 2069 | 2464 | 3037 | 1723 | 1479 |
| B | 93 | 1193 | 1380 | 2718 | 2830 | 1765 | 2037 | 1633 | 1972 | 2066 | 2372 | 1872 | 2010 | 1993 | 230 | 1820 | 1454 | 1277 | 127 | 2013 | 2453 | 9 | 1221 | 936 |
| C | 496 | 380 | 109 | 227 | 2136 | 632 | 2126 | 40 | 1689 | 1123 | 1349 | 1860 | 2189 | 180 | 1637 | 1129 | 1700 | 1165 | 486 | 1315 | 853 | 1280 | 1151 | 1232 |
| D | 170 | 688 | 490 | 254 | 11 | 847 | 1093 | 1608 | 921 | 1843 | 343 | 2240 | 1878 | 1584 | 1589 | 1862 | 1126 | 1320 | 1016 | 622 | 1299 | 379 | 1203 | 1522 |
| E | 73 | 325 | 574 | 1329 | 1124 | 1338 | 1389 | 1382 | 1088 | 1732 | 1836 | 1398 | 1985 | 1163 | 1493 | 2233 | 932 | 1486 | 1992 | 1650 | 2075 | 1767 | 1075 | 589 |
| F | 190 | 758 | 483 | 772 | 771 | 923 | 1067 | 1297 | 1291 | 135 | 1120 | 934 | 1525 | 1025 | 1286 | 1070 | 1286 | 1317 | 1370 | 1659 | 1189 | 925 | 714 | 650 |
| G | 255 | 304 | 465 | 865 | 1134 | 1369 | 967 | 1247 | 845 | 835 | 990 | 339 | 1123 | 1209 | 84 | 893 | 1073 | 931 | 981 | 751 | 1680 | 1489 | 1054 | 1327 |
| H | 285 | 386 | 692 | 640 | 1118 | 571 | 79 | 706 | 976 | 69 | 689 | 508 | 552 | 391 | 345 | 636 | 433 | 512 | 1031 | 1017 | 952 | 1239 | 869 | 1293 |
| I | 359 | 72 | 1712 | 1113 | 942 | 920 | 1136 | 975 | 1209 | 1104 | 1030 | 314 | 617 | 1180 | 1376 | 1262 | 1285 | 1203 | 1116 | 1293 | 1063 | 1428 | 1050 | 1916 |
| J | 470 | 188 | 495 | 1177 | 858 | 586 | 1315 | 7 | 1087 | 1165 | 1025 | 307 | 441 | 1000 | 238 | 1188 | 576 | 242 | 451 | 1899 | 1218 | 509 | 668 | 885 |
| K | 414 | 133 | 542 | 923 | 984 | 871 | 227 | 384 | 1138 | 679 | 1313 | 592 | 968 | 985 | 917 | 1480 | 1223 | 1350 | 1583 | 1409 | 692 | 649 | 1026 | 1172 |
| L | 349 | 741 | 561 | 758 | 106 | 232 | 405 | 355 | 710 | 777 | 956 | 586 | 297 | 1216 | 1294 | 1075 | 944 | 1293 | 163 | 675 | 695 | 569 | 779 | 746 |
| M | 432 | 102 | 604 | 457 | 678 | 502 | 668 | 856 | 531 | 552 | 988 | 1090 | 1357 | 1022 | 964 | 602 | 948 | 1256 | 663 | 67 | 508 | 696 | 526 | 982 |
| N | 590 | 383 | 604 | 286 | 350 | 277 | 444 | 425 | 293 | 301 | 616 | 702 | 594 | 376 | 581 | 540 | 732 | 296 | 1129 | 157 | 784 | 606 | 462 | 825 |
| O | 1336 | 225 | 156 | 503 | 290 | 349 | 879 | 205 | 169 | 357 | 663 | 466 | 985 | 357 | 823 | 433 | 1395 | 982 | 815 | 529 | 660 | 1430 | 477 | 958 |
| P | 272 | 329 | 252 | 213 | 794 | 235 | 301 | 478 | 144 | 308 | 609 | 447 | 731 | 1117 | 21 | 984 | 773 | 183 | 591 | 708 | 1142 | 1420 | 1617 | 1455 |

**TEXAS RED**

| Row La | 01 | 02 | 03 | 04 | 05 | 06 | 07 | 08 | 09 | 10 | 11 | 12 | 13 | 14 | 15 | 16 | 17 | 18 | 19 | 20 | 21 | 22 | 23 | 24 |
|---|---|---|---|---|---|---|---|---|---|---|---|---|---|---|---|---|---|---|---|---|---|---|---|---|
| A | 682 | 882 | 560 | 499 | 521 | 370 | 442 | 311 | 400 | 459 | 491 | 357 | 436 | 392 | 407 | 870 | 306 | 266 | 341 | 346 | 316 | 278 | 331 | 404 |
| B | 835 | 381 | 363 | 270 | 252 | 268 | 324 | 311 | 324 | 280 | 313 | 305 | 318 | 321 | 781 | 302 | 301 | 419 | 846 | 303 | 284 | 570 | 373 | 424 |
| C | 430 | 683 | 950 | 673 | 301 | 465 | 263 | 1108 | 326 | 370 | 351 | 270 | 251 | 772 | 337 | 393 | 378 | 413 | 525 | 382 | 425 | 341 | 351 | 404 |
| D | 790 | 518 | 550 | 775 | 615 | 402 | 367 | 292 | 390 | 279 | 671 | 287 | 326 | 297 | 324 | 294 | 404 | 362 | 368 | 508 | 392 | 597 | 339 | 332 |
| E | 620 | 562 | 432 | 324 | 325 | 349 | 326 | 316 | 365 | 320 | 298 | 346 | 357 | 348 | 328 | 294 | 378 | 321 | 300 | 329 | 301 | 316 | 353 | 497 |
| F | 584 | 424 | 459 | 449 | 399 | 403 | 396 | 355 | 355 | 536 | 389 | 378 | 337 | 340 | 327 | 352 | 331 | 339 | 323 | 317 | 332 | 401 | 484 | 466 |
| G | 721 | 565 | 567 | 422 | 381 | 391 | 430 | 410 | 426 | 436 | 383 | 617 | 381 | 377 | 1007 | 444 | 393 | 439 | 411 | 453 | 342 | 353 | 389 | 385 |
| H | 632 | 598 | 538 | 454 | 372 | 530 | 1037 | 448 | 444 | 1023 | 494 | 517 | 559 | 626 | 615 | 515 | 529 | 521 | 394 | 393 | 453 | 405 | 528 | 417 |
| I | 631 | 927 | 312 | 375 | 444 | 411 | 377 | 393 | 372 | 389 | 444 | 560 | 593 | 431 | 361 | 406 | 395 | 388 | 406 | 336 | 437 | 403 | 466 | 311 |
| J | 578 | 740 | 573 | 417 | 445 | 516 | 382 | 743 | 446 | 370 | 439 | 608 | 622 | 434 | 757 | 411 | 518 | 689 | 543 | 334 | 426 | 499 | 483 | 470 |
| K | 618 | 808 | 594 | 490 | 453 | 471 | 621 | 629 | 387 | 544 | 363 | 515 | 424 | 442 | 475 | 358 | 417 | 436 | 330 | 370 | 541 | 551 | 497 | 386 |
| L | 701 | 540 | 584 | 491 | 844 | 755 | 640 | 591 | 506 | 519 | 491 | 523 | 648 | 398 | 403 | 450 | 418 | 406 | 869 | 566 | 551 | 520 | 489 | 489 |
| M | 658 | 858 | 498 | 529 | 493 | 569 | 562 | 512 | 585 | 580 | 467 | 435 | 391 | 439 | 491 | 535 | 460 | 435 | 574 | 965 | 616 | 580 | 581 | 461 |
| N | 598 | 671 | 577 | 744 | 650 | 724 | 620 | 572 | 729 | 700 | 489 | 546 | 612 | 646 | 573 | 537 | 604 | 764 | 392 | 930 | 509 | 638 | 522 | 529 |
| O | 393 | 746 | 834 | 544 | 648 | 713 | 442 | 802 | 699 | 600 | 597 | 595 | 442 | 566 | 468 | 569 | 436 | 481 | 522 | 557 | 588 | 383 | 642 | 471 |
| P | 621 | 708 | 762 | 745 | 465 | 744 | 647 | 616 | 920 | 669 | 552 | 621 | 500 | 541 | 796 | 574 | 490 | 864 | 537 | 565 | 480 | 455 | 355 | 354 |

### Diverset Plate 9

DAPI (Cell Conter)

| Row | 01 | 02 | 03 | 04 | 05 | 06 | 07 | 08 | 09 | 10 | 11 | 12 | 13 | 14 | 15 | 16 | 17 | 18 | 19 | 20 | 21 | 22 | 23 | 24 |
|---|---|---|---|---|---|---|---|---|---|---|---|---|---|---|---|---|---|---|---|---|---|---|---|---|
| A | 657 | 900 | 871 | 746 | 1390 | 1427 | 2216 | 1175 | 247 | 2240 | 665 | 1113 | 1428 | 1197 | 1572 | 851 | 912 | 398 | 1703 | 283 | 774 | 813 | 789 | 841 |
| B | 1126 | 735 | 1715 | 1935 | 1331 | 2113 | 1328 | 1491 | 1437 | 1579 | 1063 | 552 | 375 | 1013 | 1786 | 759 | 1291 | 516 | 1104 | 595 | 711 | 734 | 780 | 664 |
| C | 428 | 580 | 2036 | 91 | 1330 | 142 | 1331 | 855 | 853 | 1004 | 866 | 103 | 1120 | 715 | 50 | 844 | 142 | 641 | 839 | 975 | 249 | 1392 | 441 | 422 |
| D | 415 | 425 | 857 | 655 | 923 | 787 | 751 | 1059 | 1097 | 749 | 895 | 591 | 126 | 564 | 675 | 561 | 1391 | 779 | 730 | 540 | 1079 | 689 | 280 | 141 |
| E | 386 | 537 | 626 | 1053 | 635 | 1172 | 1857 | 1381 | 1275 | 988 | 605 | 1033 | 1386 | 875 | 916 | 1139 | 762 | 1062 | 622 | 456 | 873 | 762 | 289 | 151 |
| F | 751 | 816 | 1100 | 957 | 903 | 907 | 1014 | 919 | 1446 | 926 | 1283 | 845 | 980 | 895 | 617 | 943 | 958 | 600 | 597 | 539 | 288 | 430 | 281 | 86 |
| G | 419 | 835 | 780 | 622 | 423 | 1084 | 461 | 581 | 1186 | 881 | 441 | 113 | 809 | 1173 | 811 | 385 | 937 | 751 | 427 | 116 | 580 | 500 | 418 | 127 |
| H | 812 | 162 | 423 | 1057 | 710 | 450 | 671 | 593 | 707 | 695 | 803 | 662 | 758 | 631 | 981 | 768 | 484 | 377 | 801 | 546 | 467 | 247 | 117 | 161 |
| I | 480 | 656 | 278 | 1189 | 1247 | 490 | 819 | 18 | 841 | 1202 | 996 | 1239 | 104 | 1347 | 1269 | 1200 | 1499 | 925 | 879 | 561 | 589 | 265 | 347 | 278 |
| J | 372 | 515 | 1175 | 232 | 171 | 900 | 1070 | 1127 | 1058 | 674 | 1059 | 946 | 1172 | 1025 | 1233 | 731 | 962 | 768 | 649 | 603 | 559 | 277 | 224 | 427 |
| K | 826 | 1286 | 2139 | 628 | 763 | 2156 | 111 | 1758 | 1316 | 1079 | 1158 | 1064 | 1361 | 993 | 1093 | 1368 | 1107 | 1278 | 852 | 204 | 592 | 411 | 253 | 620 |
| L | 949 | 460 | 89 | 336 | 1391 | 905 | 1494 | 1011 | 1257 | 1118 | 1552 | 998 | 974 | 1037 | 332 | 999 | 980 | 754 | 913 | 594 | 802 | 389 | 230 | 489 |
| M | 681 | 1884 | 678 | 551 | 364 | 1246 | 274 | 1200 | 1826 | 1390 | 1266 | 1310 | 1006 | 398 | 1214 | 870 | 770 | 905 | 20 | 717 | 656 | 235 | 215 | 167 |
| N | 1888 | 818 | 1090 | 587 | 917 | 730 | 458 | 844 | 856 | 739 | 1159 | 947 | 609 | 476 | 720 | 599 | 564 | 431 | 522 | 253 | 447 | 230 | 225 | 170 |
| O | 940 | 1669 | 433 | 1259 | 1109 | 357 | 333 | 641 | 790 | 767 | 741 | 877 | 75 | 544 | 537 | 477 | 45 | 379 | 228 | 494 | 401 | 302 | 343 | 350 |
| P | 1892 | 1020 | 1129 | 1458 | 1726 | 1459 | 1853 | 1292 | 1109 | 1651 | 1517 | 817 | 1373 | 1045 | 1382 | 959 | 936 | 762 | 719 | 536 | 828 | 436 | 878 | 639 |

TEXAS RED

| Row | 01 | 02 | 03 | 04 | 05 | 06 | 07 | 08 | 09 | 10 | 11 | 12 | 13 | 14 | 15 | 16 | 17 | 18 | 19 | 20 | 21 | 22 | 23 | 24 |
|---|---|---|---|---|---|---|---|---|---|---|---|---|---|---|---|---|---|---|---|---|---|---|---|---|
| A | 413 | 403 | 427 | 418 | 331 | 309 | 301 | 400 | 752 | 310 | 490 | 396 | 385 | 408 | 335 | 451 | 457 | 596 | 332 | 644 | 460 | 346 | 440 | 416 |
| B | 301 | 605 | 381 | 347 | 333 | 353 | 561 | 359 | 374 | 358 | 452 | 488 | 588 | 452 | 313 | 479 | 382 | 536 | 410 | 485 | 458 | 512 | 449 | 483 |
| C | 527 | 564 | 279 | 928 | 327 | 831 | 441 | 478 | 434 | 462 | 474 | 952 | 320 | 512 | 973 | 516 | 853 | 528 | 433 | 342 | 697 | 359 | 589 | 569 |
| D | 698 | 797 | 554 | 540 | 403 | 447 | 486 | 337 | 385 | 483 | 411 | 548 | 886 | 565 | 515 | 569 | 330 | 486 | 448 | 543 | 472 | 490 | 809 | 677 |
| E | 467 | 379 | 387 | 395 | 373 | 342 | 287 | 335 | 324 | 402 | 367 | 322 | 355 | 364 | 371 | 349 | 407 | 344 | 433 | 446 | 420 | 447 | 429 | 550 |
| F | 455 | 515 | 442 | 379 | 405 | 391 | 388 | 375 | 342 | 368 | 386 | 390 | 391 | 357 | 410 | 437 | 433 | 443 | 452 | 496 | 561 | 523 | 642 | 503 |
| G | 628 | 504 | 477 | 522 | 481 | 437 | 535 | 611 | 375 | 440 | 593 | 845 | 447 | 413 | 442 | 592 | 437 | 459 | 589 | 687 | 551 | 557 | 526 | 833 |
| H | 457 | 821 | 654 | 476 | 508 | 633 | 560 | 530 | 468 | 541 | 519 | 499 | 468 | 541 | 435 | 511 | 580 | 627 | 521 | 559 | 547 | 663 | 856 | 679 |
| I | 633 | 518 | 809 | 417 | 435 | 590 | 481 | 986 | 477 | 432 | 415 | 428 | 873 | 413 | 402 | 401 | 400 | 411 | 532 | 630 | 539 | 791 | 657 | 718 |
| J | 485 | 670 | 427 | 503 | 859 | 481 | 481 | 438 | 455 | 485 | 478 | 438 | 425 | 418 | 429 | 524 | 492 | 475 | 639 | 624 | 554 | 664 | 719 | 728 |
| K | 533 | 401 | 334 | 554 | 520 | 313 | 938 | 400 | 435 | 429 | 422 | 454 | 388 | 465 | 482 | 424 | 441 | 451 | 523 | 775 | 596 | 691 | 653 | 593 |
| L | 466 | 632 | 851 | 743 | 432 | 487 | 416 | 459 | 400 | 446 | 433 | 523 | 444 | 459 | 661 | 511 | 529 | 518 | 444 | 529 | 536 | 709 | 846 | 679 |
| M | 614 | 341 | 586 | 642 | 758 | 452 | 746 | 453 | 435 | 422 | 425 | 437 | 483 | 680 | 449 | 536 | 569 | 498 | 971 | 554 | 512 | 716 | 804 | 819 |
| N | 366 | 531 | 505 | 571 | 539 | 587 | 627 | 568 | 545 | 598 | 552 | 601 | 671 | 674 | 610 | 551 | 660 | 672 | 606 | 842 | 697 | 670 | 736 | 616 |
| O | 506 | 418 | 724 | 457 | 472 | 730 | 679 | 609 | 540 | 443 | 529 | 484 | 1005 | 633 | 630 | 643 | 737 | 715 | 755 | 603 | 734 | 735 | 756 | 741 |
| P | 323 | 464 | 490 | 427 | 383 | 375 | 422 | 360 | 431 | 352 | 378 | 494 | 423 | 426 | 445 | 436 | 560 | 578 | 510 | 596 | 442 | 619 | 457 | 672 |

### Diverset Plate 10

DAPI (Cell Conter)

| Row | 01 | 02 | 03 | 04 | 05 | 06 | 07 | 08 | 09 | 10 | 11 | 12 | 13 | 14 | 15 | 16 | 17 | 18 | 19 | 20 | 21 | 22 | 23 | 24 |
|---|---|---|---|---|---|---|---|---|---|---|---|---|---|---|---|---|---|---|---|---|---|---|---|---|
| A | 1965 | 387 | 573 | 644 | 1258 | 2769 | 1589 | 1840 | 1114 | 1049 | 877 | 2607 | 2501 | 2135 | 629 | 1321 | 789 | 994 | 1270 | 1425 | 467 | 1155 | 1128 | 1 |
| B | 691 | 568 | 1718 | 792 | 1813 | 761 | 578 | 644 | 796 | 1221 | 22 | 1274 | 686 | 1326 | 871 | 775 | 763 | 750 | 1261 | 1279 | 1299 | 784 | 898 | 1 |
| C | 117 | 158 | 59 | 541 | 1362 | 493 | 1619 | 751 | 1010 | 1654 | 2067 | 1428 | 919 | 804 | 703 | 526 | 1263 | 836 | 865 | 607 | 764 | 358 | 434 | 0 |
| D | 795 | 715 | 935 | 652 | 661 | 87 | 966 | 1609 | 1601 | 1141 | 1372 | 1042 | 1133 | 99 | 950 | 631 | 1459 | 727 | 141 | 839 | 946 | 304 | 449 | 2 |
| E | 28 | 299 | 69 | 1186 | 1191 | 926 | 827 | 505 | 1151 | 829 | 571 | 813 | 767 | 812 | 654 | 499 | 497 | 764 | 715 | 1202 | 719 | 287 | 516 | 0 |
| F | 5 | 303 | 771 | 856 | 1578 | 727 | 945 | 1032 | 321 | 744 | 767 | 519 | 1004 | 1070 | 237 | 924 | 652 | 674 | 941 | 634 | 723 | 661 | 416 | 0 |
| G | 98 | 185 | 87 | 1159 | 520 | 689 | 777 | 658 | 633 | 306 | 894 | 302 | 896 | 458 | 1038 | 1226 | 521 | 637 | 787 | 709 | 513 | 92 | 587 | 0 |
| H | 213 | 565 | 677 | 875 | 946 | 1013 | 548 | 1161 | 680 | 956 | 258 | 314 | 412 | 816 | 317 | 429 | 321 | 945 | 355 | 725 | 556 | 400 | 546 | 0 |
| I | 56 | 251 | 428 | 922 | 589 | 587 | 1078 | 640 | 801 | 296 | 1821 | 466 | 619 | 941 | 647 | 1008 | 976 | 996 | 890 | 897 | 1040 | 606 | 429 | 0 |
| J | 524 | 594 | 1030 | 554 | 1107 | 772 | 1341 | 1214 | 760 | 604 | 737 | 1666 | 374 | 969 | 565 | 613 | 108 | 1153 | 1397 | 443 | 829 | 1029 | 397 | 0 |
| K | 451 | 250 | 178 | 1595 | 1189 | 1270 | 868 | 1318 | 1365 | 835 | 860 | 1388 | 811 | 533 | 362 | 649 | 951 | 1066 | 915 | 840 | 1085 | 794 | 287 | 0 |
| L | 1679 | 1341 | 790 | 1320 | 1209 | 1026 | 1188 | 1817 | 949 | 302 | 1205 | 8 | 1279 | 797 | 718 | 638 | 804 | 1006 | 768 | 1104 | 1131 | 680 | 262 | 0 |
| M | 903 | 822 | 714 | 1109 | 20 | 1398 | 1174 | 1718 | 1827 | 962 | 1297 | 1184 | 1136 | 1465 | 1019 | 996 | 417 | 1055 | 917 | 965 | 211 | 500 | 276 | 1 |
| N | 1072 | 1084 | 1232 | 893 | 1103 | 100 | 1118 | 1223 | 914 | 1057 | 937 | 198 | 1249 | 685 | 773 | 683 | 482 | 891 | 835 | 902 | 523 | 208 | 208 | 0 |
| O | 1580 | 665 | 334 | 1584 | 1144 | 1129 | 1026 | 849 | 306 | 896 | 1367 | 511 | 1214 | 1243 | 568 | 350 | 724 | 756 | 466 | 699 | 741 | 285 | 323 | 1 |
| P | 1645 | 2470 | 1193 | 1072 | 502 | 1889 | 1755 | 1898 | 1189 | 1317 | 989 | 852 | 881 | 864 | 609 | 1026 | 507 | 596 | 383 | 460 | 618 | 433 | 592 | 0 |

TEXAS RED

| Row | 01 | 02 | 03 | 04 | 05 | 06 | 07 | 08 | 09 | 10 | 11 | 12 | 13 | 14 | 15 | 16 | 17 | 18 | 19 | 20 | 21 | 22 | 23 | 24 |
|---|---|---|---|---|---|---|---|---|---|---|---|---|---|---|---|---|---|---|---|---|---|---|---|---|
| A | 236 | 423 | 423 | 443 | 311 | 240 | 302 | 290 | 408 | 442 | 412 | 271 | 263 | 285 | 560 | 393 | 513 | 396 | 402 | 364 | 613 | 404 | 381 | 28 |
| B | 473 | 491 | 318 | 479 | 358 | 422 | 487 | 485 | 451 | 349 | 928 | 389 | 519 | 405 | 427 | 451 | 467 | 470 | 404 | 364 | 352 | 423 | 363 | 29 |
| C | 705 | 835 | 1017 | 456 | 331 | 543 | 440 | 547 | 410 | 321 | 289 | 369 | 439 | 443 | 490 | 559 | 323 | 470 | 455 | 473 | 396 | 544 | 462 | 0 |
| D | 428 | 535 | 442 | 497 | 476 | 852 | 427 | 327 | 358 | 380 | 340 | 434 | 381 | 774 | 389 | 461 | 340 | 414 | 732 | 442 | 404 | 589 | 548 | 28 |
| E | 730 | 448 | 453 | 167 | 148 | 194 | 324 | 178 | 185 | 198 | 385 | 222 | 311 | 276 | 376 | 407 | 388 | 293 | 328 | 231 | 248 | 310 | 315 | 0 |
| F | 217 | 479 | 401 | 399 | 304 | 398 | 364 | 376 | 550 | 412 | 412 | 483 | 366 | 383 | 517 | 369 | 402 | 415 | 374 | 437 | 431 | 413 | 475 | 0 |
| G | 697 | 855 | 767 | 365 | 537 | 449 | 473 | 424 | 458 | 545 | 439 | 618 | 408 | 521 | 361 | 334 | 456 | 430 | 404 | 428 | 484 | 791 | 487 | 0 |
| H | 643 | 586 | 456 | 416 | 412 | 453 | 513 | 381 | 459 | 404 | 650 | 672 | 561 | 485 | 638 | 542 | 528 | 403 | 548 | 459 | 471 | 619 | 520 | 0 |
| I | 790 | 746 | 595 | 465 | 498 | 495 | 438 | 561 | 464 | 724 | 335 | 608 | 495 | 454 | 499 | 395 | 445 | 458 | 448 | 435 | 406 | 512 | 632 | 0 |
| J | 641 | 500 | 476 | 593 | 418 | 488 | 425 | 339 | 501 | 522 | 451 | 340 | 611 | 430 | 579 | 546 | 736 | 428 | 398 | 577 | 455 | 456 | 572 | 0 |
| K | 652 | 788 | 843 | 395 | 444 | 392 | 472 | 355 | 401 | 493 | 468 | 410 | 536 | 599 | 682 | 523 | 445 | 418 | 445 | 467 | 471 | 492 | 654 | 0 |
| L | 372 | 366 | 509 | 381 | 415 | 470 | 427 | 341 | 469 | 715 | 409 | 1057 | 401 | 473 | 517 | 528 | 509 | 459 | 516 | 434 | 420 | 591 | 721 | 0 |
| M | 592 | 406 | 532 | 475 | 962 | 421 | 406 | 367 | 349 | 497 | 431 | 403 | 433 | 411 | 448 | 438 | 591 | 392 | 531 | 444 | 701 | 598 | 748 | 140 |
| N | 492 | 496 | 458 | 463 | 454 | 867 | 497 | 465 | 524 | 512 | 472 | 853 | 463 | 625 | 552 | 587 | 571 | 503 | 504 | 473 | 546 | 768 | 776 | 0 |
| O | 359 | 450 | 638 | 363 | 405 | 398 | 472 | 457 | 670 | 435 | 399 | 549 | 464 | 377 | 569 | 663 | 517 | 526 | 595 | 473 | 614 | 730 | 645 | 29 |
| P | 350 | 331 | 472 | 449 | 582 | 298 | 406 | 363 | 398 | 435 | 486 | 473 | 507 | 529 | 572 | 442 | 562 | 538 | 620 | 601 | 564 | 697 | 607 | 0 |

### Diverset Plate 11

DAPI (Cell Conter)

| Row La | 01 | 02 | 03 | 04 | 05 | 06 | 07 | 08 | 09 | 10 | 11 | 12 | 13 | 14 | 15 | 16 | 17 | 18 | 19 | 20 | 21 | 22 | 23 | 24 |
|---|---|---|---|---|---|---|---|---|---|---|---|---|---|---|---|---|---|---|---|---|---|---|---|---|
| A | 680 | 1606 | 1924 | 1078 | 2106 | 2233 | 3108 | 67 | 1882 | 3234 | 2452 | 2559 | 2134 | 2253 | 602 | 1868 | 2800 | 1259 | 1825 | 1213 | 2475 | 1450 | 2769 | 3100 |
| B | 2092 | 1870 | 1155 | 1158 | 2292 | 1783 | 2273 | 376 | 2513 | 2425 | 1715 | 1958 | 2291 | 2534 | 135 | 389 | 1872 | 1512 | 2111 | 2328 | 1875 | 1032 | 2175 | 3186 |
| C | 8 | 659 | 510 | 1303 | 1629 | 1358 | 1412 | 1765 | 358 | 1524 | 1169 | 1955 | 2607 | 1516 | 1901 | 1567 | 1658 | 1707 | 1597 | 931 | 1352 | 1483 | 980 | 3396 |
| D | 1269 | 1037 | 1416 | 1757 | 1188 | 1337 | 1388 | 1881 | 505 | 1310 | 2030 | 1443 | 1594 | 2143 | 2104 | 233 | 531 | 1702 | 635 | 1921 | 1464 | 1851 | 941 | 3025 |
| E | 89 | 939 | 1488 | 2230 | 1435 | 1506 | 1393 | 29 | 1038 | 42 | 22 | 1832 | 97 | 10 | 55 | 2069 | 98 | 24 | 476 | 176 | 165 | 2176 | 103 | 1195 |
| F | 54 | 511 | 74 | 45 | 880 | 1394 | 785 | 1159 | 118 | 144 | 2235 | 73 | 65 | 1597 | 138 | 2018 | 1938 | 138 | 1682 | 182 | 203 | 1963 | 251 | 285 |
| G | 363 | 494 | 357 | 850 | 1007 | 1568 | 982 | 1212 | 1321 | 1168 | 175 | 1368 | 1017 | 857 | 652 | 110 | 1573 | 1353 | 1527 | 1546 | 1789 | 1459 | 1601 | 2813 |
| H | 375 | 777 | 22 | 1113 | 19 | 1052 | 985 | 1423 | 1097 | 1262 | 943 | 116 | 1290 | 909 | 986 | 2083 | 1325 | 1267 | 1036 | 1783 | 1079 | 533 | 1588 | 2925 |
| I | 824 | 830 | 935 | 1165 | 1305 | 760 | 834 | 1360 | 650 | 1455 | 884 | 823 | 1166 | 1371 | 1496 | 859 | 1279 | 1469 | 1175 | 1575 | 1387 | 1439 | 2002 | 3093 |
| J | 1090 | 571 | 1215 | 1167 | 465 | 1281 | 1357 | 1304 | 9 | 1388 | 1707 | 154 | 1351 | 1336 | 1750 | 1758 | 1853 | 1611 | 1343 | 1454 | 1238 | 1276 | 1610 | 2809 |
| K | 576 | 448 | 581 | 1014 | 797 | 1210 | 1598 | 1226 | 1500 | 838 | 1436 | 236 | 643 | 1237 | 1253 | 1434 | 1085 | 1246 | 1404 | 1114 | 1732 | 1714 | 1767 | 3493 |
| L | 1098 | 1118 | 1040 | 1280 | 938 | 1611 | 1393 | 1835 | 1408 | 904 | 629 | 1376 | 1333 | 1367 | 1316 | 1168 | 1723 | 1458 | 832 | 1565 | 1089 | 1848 | 1130 | 4192 |
| M | 1346 | 264 | 121 | 642 | 344 | 1099 | 493 | 125 | 1196 | 992 | 398 | 76 | 1321 | 1449 | 1114 | 1577 | 1852 | 1928 | 1546 | 1868 | 1955 | 826 | 1614 | 4388 |
| N | 1565 | 1255 | 1243 | 1054 | 1196 | 1256 | 1137 | 999 | 1551 | 680 | 1783 | 1082 | 1231 | 607 | 1466 | 1548 | 1453 | 1357 | 1484 | 1010 | 2562 | 1758 | 1107 | 2473 |
| O | 250 | 203 | 29 | 350 | 575 | 188 | 852 | 612 | 559 | 356 | 554 | 750 | 542 | 926 | 674 | 1611 | 759 | 631 | 862 | 953 | 670 | 929 | 1279 | 2339 |
| P | 487 | 428 | 201 | 504 | 798 | 439 | 298 | 447 | 390 | 818 | 828 | 684 | 507 | 169 | 943 | 1077 | 651 | 495 | 447 | 182 | 805 | 722 | 1373 | 1442 |

TEXAS RED

| Row La | 01 | 02 | 03 | 04 | 05 | 06 | 07 | 08 | 09 | 10 | 11 | 12 | 13 | 14 | 15 | 16 | 17 | 18 | 19 | 20 | 21 | 22 | 23 | 24 |
|---|---|---|---|---|---|---|---|---|---|---|---|---|---|---|---|---|---|---|---|---|---|---|---|---|
| A | 426 | 388 | 305 | 385 | 306 | 317 | 235 | 761 | 322 | 273 | 261 | 282 | 269 | 289 | 446 | 303 | 262 | 356 | 291 | 382 | 290 | 357 | 259 | 116 |
| B | 388 | 367 | 430 | 439 | 330 | 353 | 298 | 691 | 302 | 313 | 318 | 289 | 296 | 278 | 802 | 392 | 337 | 325 | 303 | 287 | 295 | 443 | 284 | 117 |
| C | 293 | 515 | 489 | 376 | 321 | 349 | 326 | 320 | 619 | 317 | 425 | 280 | 262 | 337 | 301 | 324 | 305 | 305 | 319 | 373 | 345 | 361 | 389 | 115 |
| D | 477 | 485 | 367 | 315 | 367 | 421 | 367 | 310 | 504 | 373 | 297 | 411 | 332 | 284 | 293 | 436 | 490 | 306 | 460 | 292 | 344 | 301 | 426 | 114 |
| E | 574 | 379 | 293 | 276 | 290 | 291 | 367 | 384 | 345 | 382 | 481 | 263 | 355 | 750 | 375 | 272 | 363 | 573 | 396 | 384 | 415 | 271 | 391 | 127 |
| F | 325 | 417 | 404 | 458 | 443 | 323 | 449 | 283 | 327 | 332 | 265 | 391 | 406 | 284 | 368 | 293 | 303 | 333 | 287 | 392 | 366 | 306 | 272 | 126 |
| G | 624 | 500 | 592 | 407 | 390 | 356 | 444 | 377 | 368 | 378 | 661 | 346 | 462 | 446 | 457 | 842 | 382 | 347 | 339 | 320 | 322 | 360 | 349 | 118 |
| H | 720 | 531 | 725 | 440 | 496 | 412 | 470 | 373 | 389 | 351 | 420 | 685 | 411 | 476 | 442 | 316 | 412 | 370 | 442 | 341 | 442 | 587 | 318 | 116 |
| I | 525 | 503 | 403 | 486 | 347 | 505 | 507 | 379 | 483 | 302 | 424 | 438 | 426 | 363 | 355 | 461 | 407 | 359 | 412 | 336 | 420 | 383 | 286 | 116 |
| J | 475 | 539 | 386 | 408 | 612 | 451 | 433 | 406 | 652 | 409 | 364 | 679 | 388 | 392 | 368 | 399 | 352 | 390 | 440 | 373 | 458 | 460 | 418 | 116 |
| K | 596 | 533 | 595 | 450 | 483 | 416 | 323 | 414 | 367 | 555 | 388 | 600 | 538 | 439 | 408 | 392 | 435 | 444 | 371 | 421 | 359 | 377 | 388 | 117 |
| L | 531 | 435 | 391 | 460 | 554 | 393 | 432 | 385 | 424 | 532 | 609 | 393 | 424 | 443 | 421 | 427 | 371 | 406 | 492 | 362 | 393 | 353 | 488 | 117 |
| M | 411 | 739 | 890 | 592 | 584 | 449 | 596 | 899 | 404 | 453 | 576 | 1015 | 424 | 434 | 476 | 396 | 377 | 329 | 394 | 371 | 373 | 535 | 390 | 115 |
| N | 448 | 476 | 493 | 482 | 469 | 454 | 407 | 500 | 436 | 556 | 396 | 500 | 434 | 636 | 420 | 413 | 377 | 397 | 379 | 470 | 315 | 398 | 600 | 124 |
| O | 841 | 752 | 746 | 647 | 577 | 786 | 494 | 544 | 508 | 637 | 592 | 566 | 525 | 478 | 504 | 421 | 463 | 492 | 482 | 464 | 555 | 430 | 391 | 117 |
| P | 666 | 582 | 832 | 640 | 463 | 604 | 740 | 659 | 633 | 580 | 526 | 538 | 568 | 829 | 520 | 484 | 500 | 545 | 566 | 835 | 469 | 466 | 353 | 117 |

### Diverset Plate 12

DAPI (Cell Conter)

| Row La | 01 | 02 | 03 | 04 | 05 | 06 | 07 | 08 | 09 | 10 | 11 | 12 | 13 | 14 | 15 | 16 | 17 | 18 | 19 | 20 | 21 | 22 | 23 | 24 |
|---|---|---|---|---|---|---|---|---|---|---|---|---|---|---|---|---|---|---|---|---|---|---|---|---|
| A | 1151 | 892 | 593 | 1078 | 950 | 1027 | 1215 | 1402 | 1016 | 2214 | 1148 | 1447 | 1577 | 1593 | 79 | 1481 | 53 | 1305 | 1514 | 1427 | 1264 | 1191 | 2043 | 1938 |
| B | 1285 | 570 | 576 | 889 | 918 | 946 | 1111 | 758 | 497 | 1360 | 918 | 1090 | 960 | 929 | 1162 | 1187 | 1055 | 879 | 1047 | 266 | 1502 | 1193 | 1107 | 1256 |
| C | 871 | 262 | 415 | 419 | 640 | 128 | 481 | 669 | 711 | 1104 | 741 | 746 | 1014 | 1199 | 82 | 1095 | 719 | 758 | 1062 | 1095 | 790 | 1474 | 852 | 696 |
| D | 798 | 17 | 671 | 699 | 979 | 410 | 992 | 688 | 1162 | 947 | 1029 | 939 | 714 | 457 | 781 | 364 | 1094 | 904 | 912 | 1035 | 961 | 190 | 688 | 964 |
| E | 1039 | 702 | 976 | 1969 | 1890 | 969 | 2014 | 1882 | 1359 | 1578 | 1514 | 238 | 2052 | 1524 | 71 | 1847 | 83 | 1395 | 2123 | 1971 | 2236 | 1668 | 1399 | 1022 |
| F | 591 | 671 | 1024 | 1140 | 1187 | 1197 | 1828 | 705 | 1121 | 1478 | 1167 | 880 | 1051 | 996 | 1029 | 915 | 1421 | 1130 | 1235 | 1867 | 1683 | 1725 | 1870 | 728 |
| G | 1328 | 782 | 1003 | 969 | 100 | 1025 | 1276 | 835 | 985 | 908 | 1094 | 1190 | 1393 | 938 | 1071 | 888 | 1095 | 1037 | 1234 | 1325 | 1396 | 1726 | 1309 | 2067 |
| H | 1266 | 706 | 821 | 944 | 327 | 919 | 1090 | 1103 | 788 | 1078 | 881 | 1088 | 1193 | 1337 | 121 | 1411 | 496 | 1290 | 1592 | 1190 | 308 | 1013 | 1488 | 1747 |
| I | 1302 | 953 | 65 | 1151 | 1218 | 1571 | 841 | 319 | 75 | 332 | 1428 | 1418 | 1673 | 1395 | 1180 | 871 | 1564 | 1759 | 1589 | 1870 | 1556 | 1439 | 1728 | 2439 |
| J | 7 | 1370 | 1535 | 1335 | 1521 | 1635 | 1289 | 1068 | 1469 | 689 | 1371 | 1433 | 1816 | 1888 | 1772 | 1048 | 1536 | 1562 | 2104 | 1860 | 1551 | 1613 | 1747 | 2191 |
| K | 1458 | 11 | 1283 | 1426 | 131 | 1330 | 1357 | 1049 | 111 | 1496 | 1776 | 1739 | 326 | 1611 | 325 | 1011 | 1472 | 1827 | 1798 | 1972 | 597 | 1768 | 1935 | 2618 |
| L | 684 | 1081 | 228 | 1529 | 1543 | 1334 | 1510 | 1351 | 1112 | 884 | 1795 | 1760 | 1339 | 233 | 1603 | 1720 | 2054 | 1787 | 1948 | 1910 | 966 | 1068 | 2028 | 1951 |
| M | 336 | 1252 | 1381 | 1377 | 851 | 1338 | 973 | 1109 | 255 | 1585 | 339 | 1393 | 71 | 1629 | 41 | 1299 | 1164 | 1770 | 1843 | 2057 | 97 | 1571 | 1655 | 2912 |
| N | 1072 | 1706 | 804 | 1000 | 1077 | 1147 | 1385 | 1580 | 1526 | 1696 | 1475 | 851 | 1201 | 1438 | 1676 | 2076 | 1966 | 1286 | 1284 | 1544 | 1907 | 1627 | 1626 | 3170 |
| O | 455 | 264 | 305 | 254 | 775 | 1184 | 1045 | 823 | 75 | 919 | 228 | 1031 | 1545 | 1606 | 1448 | 1705 | 825 | 1042 | 1182 | 1007 | 1016 | 1071 | 813 | 2122 |
| P | 398 | 721 | 351 | 373 | 536 | 913 | 969 | 807 | 595 | 896 | 363 | 611 | 901 | 836 | 1165 | 967 | 639 | 1151 | 772 | 930 | 1033 | 1076 | 1187 | 3103 |

TEXAS RED

| Row La | 01 | 02 | 03 | 04 | 05 | 06 | 07 | 08 | 09 | 10 | 11 | 12 | 13 | 14 | 15 | 16 | 17 | 18 | 19 | 20 | 21 | 22 | 23 | 24 |
|---|---|---|---|---|---|---|---|---|---|---|---|---|---|---|---|---|---|---|---|---|---|---|---|---|
| A | 435 | 457 | 504 | 428 | 437 | 427 | 468 | 382 | 356 | 296 | 414 | 353 | 341 | 372 | 851 | 371 | 805 | 366 | 356 | 401 | 368 | 394 | 304 | 118 |
| B | 466 | 573 | 667 | 612 | 493 | 489 | 401 | 498 | 529 | 414 | 463 | 433 | 455 | 370 | 432 | 424 | 452 | 456 | 441 | 734 | 382 | 404 | 418 | 119 |
| C | 497 | 667 | 670 | 547 | 538 | 1021 | 555 | 517 | 565 | 426 | 478 | 503 | 402 | 422 | 785 | 419 | 493 | 475 | 391 | 415 | 495 | 330 | 401 | 118 |
| D | 438 | 521 | 517 | 532 | 438 | 571 | 471 | 441 | 389 | 433 | 434 | 495 | 512 | 613 | 491 | 566 | 368 | 423 | 421 | 431 | 385 | 721 | 435 | 126 |
| E | 367 | 190 | 386 | 118 | 122 | 391 | 117 | 338 | 147 | 126 | 375 | 184 | 122 | 130 | 787 | 116 | 115 | 121 | 124 | 297 | 116 | 126 | 115 | 114 |
| F | 437 | 135 | 143 | 133 | 136 | 393 | 120 | 160 | 470 | 381 | 121 | 469 | 119 | 117 | 116 | 120 | 121 | 412 | 120 | 118 | 118 | 118 | 295 | 118 |
| G | 403 | 511 | 437 | 455 | 777 | 424 | 419 | 472 | 409 | 453 | 381 | 377 | 422 | 400 | 419 | 475 | 388 | 428 | 401 | 336 | 387 | 360 | 352 | 118 |
| H | 423 | 599 | 552 | 520 | 759 | 471 | 421 | 533 | 546 | 442 | 446 | 445 | 412 | 394 | 884 | 372 | 621 | 411 | 332 | 428 | 640 | 437 | 403 | 118 |
| I | 444 | 477 | 894 | 417 | 482 | 402 | 582 | 714 | 857 | 727 | 458 | 435 | 340 | 370 | 408 | 448 | 339 | 350 | 343 | 358 | 357 | 389 | 370 | 116 |
| J | 479 | 486 | 423 | 374 | 446 | 381 | 413 | 528 | 379 | 504 | 428 | 375 | 358 | 376 | 411 | 482 | 373 | 409 | 301 | 343 | 404 | 397 | 393 | 123 |
| K | 390 | 642 | 384 | 427 | 881 | 428 | 395 | 475 | 918 | 388 | 355 | 339 | 725 | 388 | 710 | 446 | 391 | 336 | 346 | 358 | 610 | 354 | 366 | 118 |
| L | 567 | 459 | 792 | 422 | 384 | 393 | 449 | 433 | 462 | 522 | 385 | 381 | 474 | 831 | 345 | 326 | 336 | 344 | 353 | 392 | 432 | 443 | 376 | 119 |
| M | 796 | 474 | 437 | 452 | 480 | 455 | 466 | 410 | 790 | 415 | 748 | 402 | 871 | 414 | 764 | 397 | 449 | 385 | 409 | 364 | 780 | 409 | 390 | 117 |
| N | 537 | 422 | 525 | 480 | 496 | 507 | 456 | 455 | 382 | 369 | 447 | 571 | 492 | 440 | 356 | 370 | 357 | 486 | 426 | 438 | 369 | 371 | 426 | 115 |
| O | 637 | 680 | 615 | 875 | 436 | 402 | 449 | 514 | 972 | 436 | 650 | 382 | 367 | 390 | 405 | 374 | 564 | 451 | 417 | 443 | 419 | 407 | 507 | 115 |
| P | 678 | 516 | 731 | 703 | 624 | 497 | 480 | 589 | 539 | 482 | 738 | 538 | 491 | 500 | 521 | 432 | 599 | 372 | 477 | 489 | 472 | 435 | 460 | 120 |

### Diverset Plate 13

**DAPI (Cell Conter)**

| Row | 01 | 02 | 03 | 04 | 05 | 06 | 07 | 08 | 09 | 10 | 11 | 12 | 13 | 14 | 15 | 16 | 17 | 18 | 19 | 20 | 21 | 22 | 23 | 24 |
|---|---|---|---|---|---|---|---|---|---|---|---|---|---|---|---|---|---|---|---|---|---|---|---|---|
| A | 1016 | 539 | 924 | 595 | 987 | 1166 | 1600 | 1283 | 1132 | 1261 | 1461 | 1786 | 898 | 1384 | 1154 | 1633 | 1585 | 1527 | 2058 | 2200 | 1150 | 1675 | 1862 | 2942 |
| B | 786 | 236 | 908 | 1179 | 896 | 858 | 951 | 1018 | 1394 | 1101 | 1169 | 1964 | 1745 | 1442 | 713 | 1751 | 1338 | 1254 | 1225 | 354 | 1168 | 1010 | 1560 | 3151 |
| C | 902 | 156 | 456 | 557 | 603 | 826 | 837 | 751 | 774 | 1085 | 573 | 1039 | 857 | 1003 | 80 | 1138 | 950 | 67 | 1208 | 150 | 1095 | 961 | 709 | 1637 |
| D | 997 | 681 | 506 | 867 | 832 | 840 | 1169 | 840 | 455 | 1012 | 942 | 1161 | 983 | 732 | 813 | 907 | 613 | 269 | 1012 | 1426 | 1470 | 307 | 1030 | 923 |
| E | 284 | 716 | 1189 | 334 | 1162 | 1251 | 1464 | 1361 | 1435 | 1387 | 1567 | 1205 | 1912 | 1828 | 841 | 1397 | 1167 | 1455 | 2114 | 1381 | 1850 | 1441 | 1299 | 907 |
| F | 542 | 835 | 1214 | 1227 | 1549 | 1115 | 1380 | 1257 | 842 | 926 | 1369 | 1168 | 1001 | 1030 | 921 | 1177 | 1534 | 1737 | 879 | 980 | 916 | 1482 | 1133 | 1104 |
| G | 825 | 454 | 649 | 753 | 1012 | 1290 | 552 | 850 | 1002 | 893 | 1118 | 1058 | 777 | 1015 | 683 | 898 | 1124 | 1140 | 934 | 883 | 1055 | 1400 | 1196 | 2676 |
| H | 1217 | 779 | 884 | 883 | 1196 | 923 | 940 | 902 | 1221 | 924 | 1407 | 954 | 849 | 951 | 1332 | 839 | 916 | 804 | 1385 | 1190 | 1032 | 1293 | 1103 | 1222 |
| I | 445 | 1579 | 1010 | 417 | 1000 | 1227 | 1458 | 1466 | 902 | 1369 | 1075 | 1294 | 1478 | 1519 | 1577 | 1088 | 1356 | 1152 | 1219 | 1381 | 1351 | 1295 | 1265 | 3295 |
| J | 1140 | 1148 | 1188 | 1090 | 1015 | 1097 | 1151 | 1146 | 1304 | 1427 | 1305 | 673 | 1381 | 1311 | 1271 | 1134 | 1109 | 1450 | 1620 | 1194 | 1522 | 849 | 1160 | 2205 |
| K | 465 | 1158 | 1129 | 1380 | 1194 | 1373 | 1323 | 1740 | 1474 | 1306 | 1312 | 1312 | 1316 | 1088 | 1006 | 1532 | 1329 | 1504 | 882 | 1698 | 1814 | 1607 | 2072 | 1378 |
| L | 1302 | 881 | 1383 | 1293 | 1549 | 1599 | 1422 | 942 | 1128 | 1848 | 575 | 1420 | 244 | 193 | 1248 | 1580 | 1434 | 1564 | 1585 | 1678 | 1749 | 1100 | 2002 | 2523 |
| M | 1105 | 1179 | 1341 | 17 | 1356 | 1500 | 1192 | 1554 | 1088 | 1115 | 1466 | 2050 | 1328 | 1302 | 1094 | 1763 | 1707 | 1020 | 1282 | 1504 | 813 | 1881 | 1902 | 922 |
| N | 1732 | 1417 | 1726 | 1574 | 1178 | 985 | 306 | 1271 | 1304 | 1009 | 9 | 1703 | 1381 | 981 | 1375 | 1100 | 1119 | 1194 | 1249 | 1443 | 1647 | 1299 | 1303 | 4013 |
| O | 212 | 219 | 363 | 1342 | 278 | 948 | 562 | 722 | 1076 | 891 | 959 | 1001 | 1532 | 778 | 942 | 1213 | 789 | 1038 | 1102 | 856 | 835 | 925 | 637 | 2826 |
| P | 436 | 831 | 375 | 285 | 1196 | 1221 | 292 | 443 | 1160 | 548 | 730 | 453 | 1193 | 507 | 467 | 788 | 422 | 710 | 771 | 615 | 632 | 943 | 1021 | 695 |

**TEXAS RED**

| Row | 01 | 02 | 03 | 04 | 05 | 06 | 07 | 08 | 09 | 10 | 11 | 12 | 13 | 14 | 15 | 16 | 17 | 18 | 19 | 20 | 21 | 22 | 23 | 24 |
|---|---|---|---|---|---|---|---|---|---|---|---|---|---|---|---|---|---|---|---|---|---|---|---|---|
| A | 458 | 522 | 468 | 536 | 513 | 410 | 357 | 400 | 509 | 464 | 397 | 361 | 467 | 398 | 406 | 431 | 388 | 327 | 377 | 348 | 433 | 348 | 316 | 119 |
| B | 556 | 585 | 439 | 445 | 491 | 506 | 484 | 474 | 418 | 484 | 448 | 357 | 376 | 430 | 512 | 411 | 428 | 492 | 400 | 655 | 386 | 454 | 366 | 118 |
| C | 494 | 796 | 719 | 563 | 564 | 506 | 552 | 491 | 508 | 449 | 542 | 400 | 451 | 455 | 821 | 371 | 358 | 893 | 390 | 570 | 415 | 387 | 499 | 117 |
| D | 479 | 552 | 596 | 501 | 533 | 413 | 400 | 599 | 683 | 381 | 434 | 396 | 391 | 470 | 489 | 390 | 516 | 737 | 396 | 352 | 363 | 654 | 467 | 118 |
| E | 486 | 400 | 351 | 503 | 359 | 357 | 322 | 355 | 365 | 371 | 339 | 351 | 292 | 339 | 366 | 313 | 339 | 296 | 259 | 297 | 283 | 336 | 315 | 114 |
| F | 531 | 532 | 384 | 397 | 347 | 473 | 422 | 396 | 370 | 417 | 340 | 432 | 465 | 464 | 298 | 390 | 357 | 350 | 401 | 407 | 417 | 354 | 418 | 122 |
| G | 577 | 660 | 500 | 464 | 368 | 380 | 638 | 407 | 416 | 412 | 406 | 430 | 496 | 442 | 443 | 423 | 364 | 386 | 397 | 475 | 400 | 365 | 438 | 118 |
| H | 438 | 538 | 518 | 478 | 493 | 425 | 505 | 429 | 429 | 498 | 436 | 443 | 498 | 419 | 401 | 426 | 467 | 467 | 372 | 389 | 414 | 368 | 459 | 117 |
| I | 739 | 378 | 479 | 641 | 435 | 428 | 424 | 420 | 460 | 438 | 436 | 410 | 352 | 369 | 365 | 456 | 358 | 411 | 405 | 389 | 373 | 432 | 356 | 118 |
| J | 469 | 449 | 469 | 465 | 365 | 419 | 429 | 426 | 391 | 418 | 474 | 508 | 404 | 396 | 475 | 441 | 425 | 390 | 402 | 427 | 388 | 459 | 407 | 116 |
| K | 557 | 457 | 471 | 420 | 354 | 415 | 408 | 345 | 420 | 412 | 448 | 438 | 422 | 422 | 382 | 394 | 402 | 399 | 445 | 366 | 372 | 355 | 379 | 117 |
| L | 435 | 465 | 461 | 464 | 402 | 407 | 437 | 473 | 430 | 388 | 420 | 437 | 865 | 716 | 422 | 394 | 404 | 361 | 380 | 362 | 397 | 410 | 373 | 116 |
| M | 519 | 398 | 457 | 792 | 431 | 402 | 415 | 361 | 450 | 484 | 429 | 332 | 405 | 432 | 451 | 385 | 404 | 442 | 459 | 426 | 487 | 385 | 395 | 116 |
| N | 438 | 426 | 404 | 490 | 464 | 558 | 854 | 447 | 482 | 486 | 883 | 439 | 479 | 528 | 509 | 462 | 483 | 477 | 446 | 445 | 394 | 457 | 452 | 116 |
| O | 754 | 774 | 708 | 452 | 780 | 498 | 602 | 514 | 406 | 419 | 457 | 414 | 373 | 457 | 484 | 371 | 506 | 456 | 487 | 515 | 531 | 473 | 560 | 117 |
| P | 642 | 588 | 639 | 764 | 409 | 427 | 705 | 675 | 460 | 644 | 533 | 607 | 471 | 681 | 580 | 513 | 630 | 520 | 498 | 568 | 516 | 436 | 459 | 124 |

### Diverset Plate 14

**DAPI (Cell Conter)**

| Row L | 01 | 02 | 03 | 04 | 05 | 06 | 07 | 08 | 09 | 10 | 11 | 12 | 13 | 14 | 15 | 16 | 17 | 18 | 19 | 20 | 21 | 22 | 23 | 24 |
|---|---|---|---|---|---|---|---|---|---|---|---|---|---|---|---|---|---|---|---|---|---|---|---|---|
| A | 977 | 885 | 1577 | 1850 | 1718 | 1528 | 1275 | 35 | 1321 | 1548 | 1787 | 2169 | 2101 | 1809 | 1133 | 1567 | 2096 | 782 | 2029 | 2156 | 2071 | 2012 | 1883 | 3689 |
| B | 757 | 869 | 1074 | 1203 | 1345 | 1243 | 1073 | 187 | 1174 | 998 | 1253 | 1502 | 1855 | 1265 | 1583 | 1524 | 1552 | 864 | 1309 | 1375 | 1263 | 999 | 1410 | 2149 |
| C | 756 | 630 | 804 | 555 | 754 | 1000 | 827 | 6 | 910 | 935 | 263 | 989 | 1204 | 797 | 740 | 917 | 896 | 789 | 1005 | 621 | 910 | 654 | 712 | 438 |
| D | 856 | 513 | 747 | 483 | 544 | 947 | 530 | 691 | 1038 | 974 | 1263 | 1222 | 535 | 977 | 1034 | 1389 | 934 | 1051 | 933 | 963 | 984 | 997 | 709 | 260 |
| E | 436 | 1297 | 27 | 1089 | 479 | 1304 | 1121 | 1478 | 1740 | 763 | 1612 | 1932 | 1573 | 1424 | 232 | 1663 | 2059 | 1520 | 2076 | 674 | 1918 | 1539 | 1449 | 844 |
| F | 79 | 484 | 353 | 1439 | 1151 | 953 | 949 | 838 | 992 | 852 | 1308 | 1206 | 1395 | 1019 | 1279 | 1295 | 1153 | 1254 | 1441 | 1647 | 1478 | 1221 | 1414 | 755 |
| G | 487 | 799 | 681 | 427 | 818 | 15 | 1220 | 1169 | 1049 | 1090 | 1125 | 1100 | 1449 | 1214 | 1073 | 887 | 1287 | 838 | 1390 | 1312 | 928 | 1353 | 1263 | 1520 |
| H | 1271 | 567 | 1117 | 954 | 1150 | 1264 | 1180 | 974 | 1506 | 801 | 860 | 1177 | 1356 | 989 | 1013 | 634 | 1101 | 1303 | 284 | 1181 | 1447 | 1245 | 1436 | 1929 |
| I | 1301 | 1093 | 1267 | 1100 | 1011 | 1306 | 924 | 1610 | 1883 | 775 | 1594 | 1275 | 1225 | 1258 | 1505 | 900 | 1390 | 1571 | 968 | 1380 | 1359 | 1555 | 1688 | 2694 |
| J | 1229 | 1422 | 215 | 1179 | 1319 | 1509 | 1386 | 287 | 1376 | 1593 | 1710 | 1244 | 1580 | 594 | 935 | 1277 | 865 | 1457 | 1521 | 1706 | 1420 | 1489 | 1957 | 2529 |
| K | 1487 | 670 | 1010 | 1005 | 1339 | 1385 | 1103 | 1832 | 1728 | 1759 | 1518 | 1347 | 1599 | 1071 | 501 | 1437 | 11 | 1380 | 1483 | 1446 | 1891 | 1763 | 1756 | 2158 |
| L | 1994 | 1110 | 1471 | 1576 | 1661 | 1545 | 1565 | 1562 | 1764 | 1324 | 1663 | 1723 | 1117 | 1271 | 1576 | 1516 | 1694 | 1329 | 1853 | 1717 | 2460 | 2137 | 1824 | 3451 |
| M | 536 | 1035 | 1495 | 403 | 1313 | 684 | 1748 | 1484 | 1176 | 1414 | 1502 | 1507 | 730 | 1476 | 8 | 1249 | 765 | 1997 | 2111 | 2084 | 2281 | 1963 | 2347 | 3249 |
| N | 2076 | 810 | 1081 | 1141 | 1290 | 1128 | 601 | 1378 | 1642 | 584 | 1539 | 1911 | 1279 | 1454 | 1040 | 1570 | 983 | 1136 | 1481 | 1845 | 1691 | 1854 | 1428 | 2808 |
| O | 270 | 516 | 767 | 563 | 810 | 783 | 796 | 539 | 697 | 575 | 1212 | 998 | 581 | 1296 | 874 | 1418 | 1177 | 1028 | 1407 | 1457 | 1306 | 1476 | 1352 | 2041 |
| P | 344 | 838 | 136 | 656 | 420 | 261 | 548 | 537 | 953 | 1330 | 1108 | 358 | 805 | 818 | 522 | 12 | 400 | 1296 | 886 | 4 | 1123 | 2072 | 1804 | 3352 |

**TEXAS RED**

| Row L | 01 | 02 | 03 | 04 | 05 | 06 | 07 | 08 | 09 | 10 | 11 | 12 | 13 | 14 | 15 | 16 | 17 | 18 | 19 | 20 | 21 | 22 | 23 | 24 |
|---|---|---|---|---|---|---|---|---|---|---|---|---|---|---|---|---|---|---|---|---|---|---|---|---|
| A | 539 | 568 | 457 | 430 | 404 | 449 | 493 | 902 | 424 | 355 | 365 | 377 | 361 | 398 | 471 | 375 | 380 | 571 | 411 | 306 | 390 | 384 | 419 | 120 |
| B | 568 | 519 | 473 | 419 | 354 | 456 | 500 | 888 | 466 | 523 | 432 | 475 | 368 | 430 | 386 | 331 | 408 | 538 | 424 | 418 | 460 | 486 | 391 | 116 |
| C | 528 | 527 | 580 | 457 | 470 | 454 | 433 | 459 | 427 | 418 | 759 | 403 | 391 | 515 | 525 | 432 | 366 | 528 | 452 | 422 | 435 | 533 | 459 | 124 |
| D | 523 | 548 | 486 | 623 | 600 | 484 | 564 | 481 | 439 | 493 | 393 | 437 | 592 | 459 | 495 | 389 | 452 | 461 | 405 | 418 | 478 | 439 | 454 | 116 |
| E | 116 | 255 | 202 | 210 | 555 | 366 | 122 | 136 | 135 | 138 | 127 | 158 | 149 | 382 | 118 | 118 | 120 | 119 | 119 | 121 | 115 | 118 | 322 | 113 |
| F | 669 | 369 | 237 | 206 | 116 | 146 | 426 | 396 | 121 | 405 | 187 | 117 | 328 | 340 | 190 | 123 | 370 | 115 | 116 | 118 | 116 | 122 | 117 | 115 |
| G | 543 | 479 | 519 | 574 | 471 | 363 | 403 | 464 | 489 | 411 | 449 | 363 | 380 | 400 | 461 | 468 | 440 | 466 | 373 | 350 | 469 | 368 | 372 | 116 |
| H | 431 | 631 | 401 | 464 | 460 | 474 | 447 | 514 | 404 | 528 | 525 | 509 | 459 | 472 | 470 | 534 | 390 | 436 | 647 | 406 | 379 | 359 | 391 | 116 |
| I | 500 | 501 | 451 | 434 | 454 | 444 | 466 | 345 | 356 | 497 | 392 | 361 | 439 | 456 | 405 | 566 | 382 | 384 | 455 | 394 | 399 | 386 | 366 | 116 |
| J | 476 | 457 | 765 | 460 | 432 | 436 | 398 | 795 | 412 | 382 | 388 | 392 | 414 | 586 | 446 | 439 | 475 | 426 | 379 | 412 | 431 | 409 | 352 | 117 |
| K | 444 | 531 | 498 | 461 | 440 | 433 | 494 | 396 | 365 | 410 | 436 | 450 | 400 | 466 | 648 | 406 | 835 | 445 | 430 | 346 | 354 | 399 | 394 | 116 |
| L | 379 | 506 | 368 | 429 | 343 | 416 | 416 | 450 | 395 | 412 | 402 | 389 | 531 | 388 | 382 | 414 | 409 | 380 | 363 | 395 | 340 | 362 | 414 | 116 |
| M | 573 | 457 | 341 | 663 | 428 | 512 | 371 | 382 | 454 | 422 | 469 | 484 | 550 | 443 | 362 | 424 | 561 | 380 | 355 | 413 | 363 | 406 | 360 | 115 |
| N | 446 | 632 | 465 | 468 | 480 | 527 | 542 | 429 | 365 | 625 | 451 | 327 | 472 | 442 | 528 | 442 | 479 | 508 | 410 | 408 | 386 | 357 | 425 | 117 |
| O | 701 | 578 | 512 | 506 | 520 | 510 | 529 | 604 | 511 | 599 | 423 | 410 | 454 | 456 | 502 | 441 | 474 | 517 | 408 | 383 | 433 | 413 | 413 | 115 |
| P | 846 | 503 | 995 | 572 | 740 | 748 | 644 | 598 | 531 | 531 | 469 | 712 | 483 | 457 | 546 | 756 | 676 | 382 | 560 | 227 | 452 | 361 | 343 | 114 |

### Diverset Plate 15

**DAPI (Cell Conter)**

| Row | 01 | 02 | 03 | 04 | 05 | 06 | 07 | 08 | 09 | 10 | 11 | 12 | 13 | 14 | 15 | 16 | 17 | 18 | 19 | 20 | 21 | 22 | 23 | 24 |
|---|---|---|---|---|---|---|---|---|---|---|---|---|---|---|---|---|---|---|---|---|---|---|---|---|
| A | 287 | 360 | 572 | 723 | 897 | 1222 | 753 | 1015 | 865 | 36 | 489 | 678 | 636 | 320 | 643 | 916 | 1849 | 319 | 518 | 359 | 486 | 173 | 1354 | 683 |
| B | 297 | 399 | 270 | 57 | 603 | 508 | 103 | 506 | 655 | 175 | 326 | 356 | 869 | 1425 | 1034 | 594 | 1044 | 602 | 343 | 399 | 572 | 663 | 1250 | 575 |
| C | 225 | 177 | 184 | 436 | 454 | 1153 | 610 | 792 | 727 | 382 | 772 | 219 | 12 | 977 | 103 | 1340 | 617 | 102 | 792 | 1625 | 715 | 553 | 340 | 349 |
| D | 136 | 204 | 541 | 540 | 292 | 786 | 769 | 1036 | 708 | 959 | 377 | 1200 | 1057 | 595 | 137 | 914 | 673 | 1109 | 1347 | 1011 | 928 | 652 | 736 | 1090 |
| E | 297 | 752 | 370 | 722 | 963 | 1778 | 1512 | 1163 | 1020 | 2055 | 1392 | 989 | 2286 | 826 | 2266 | 1446 | 1702 | 701 | 1601 | 1223 | 2018 | 147 | 1153 | 197 |
| F | 269 | 329 | 1818 | 1940 | 2577 | 2864 | 2055 | 1469 | 1143 | 1814 | 1072 | 1651 | 2302 | 1128 | 1208 | 1834 | 1808 | 1306 | 2255 | 1531 | 1753 | 1434 | 2083 | 282 |
| G | 720 | 198 | 641 | 501 | 1072 | 524 | 829 | 912 | 939 | 1352 | 782 | 738 | 1530 | 139 | 1408 | 1097 | 1322 | 1099 | 923 | 1121 | 910 | 1357 | 1562 | 1685 |
| H | 57 | 239 | 668 | 585 | 682 | 986 | 285 | 332 | 770 | 1373 | 982 | 1351 | 631 | 520 | 588 | 925 | 1562 | 1085 | 964 | 84 | 698 | 1282 | 1456 | 366 |
| I | 1482 | 1788 | 767 | 1293 | 1183 | 1571 | 1928 | 1890 | 1829 | 1939 | 1340 | 1559 | 1190 | 1988 | 1559 | 600 | 1101 | 1799 | 1704 | 1485 | 479 | 2259 | 1820 | 3079 |
| J | 810 | 1378 | 1378 | 1055 | 935 | 1660 | 1600 | 1655 | 1867 | 1395 | 1839 | 2585 | 323 | 1724 | 1456 | 2372 | 2225 | 2302 | 1418 | 1997 | 567 | 1744 | 1910 | 1761 |
| K | 1026 | 583 | 233 | 1060 | 962 | 100 | 1183 | 1367 | 1073 | 1618 | 1852 | 229 | 1041 | 1411 | 1598 | 1403 | 976 | 1084 | 1469 | 2101 | 1596 | 206 | 1964 | 2537 |
| L | 1493 | 570 | 1212 | 1476 | 1149 | 1408 | 1341 | 2164 | 1920 | 1042 | 1058 | 2405 | 1860 | 2347 | 36 | 1342 | 1476 | 1849 | 1761 | 2556 | 683 | 2232 | 2047 | 2423 |
| M | 1706 | 672 | 517 | 747 | 2340 | 715 | 1175 | 1419 | 1245 | 1980 | 2076 | 2398 | 1297 | 1699 | 1378 | 722 | 1715 | 2187 | 2246 | 3076 | 2733 | 2945 | 2548 | 2913 |
| N | 2786 | 530 | 1017 | 1088 | 1725 | 1516 | 2035 | 1435 | 2623 | 761 | 1790 | 1854 | 2819 | 1890 | 1899 | 2111 | 1798 | 1645 | 2293 | 2472 | 182 | 2364 | 2866 | 3272 |
| O | 1943 | 1468 | 1172 | 1252 | 1739 | 1747 | 437 | 1451 | 455 | 1817 | 17 | 2159 | 2409 | 1626 | 2023 | 1906 | 1797 | 473 | 2602 | 2102 | 2960 | 2126 | 2937 | 830 |
| P | 2043 | 1858 | 1159 | 1313 | 1227 | 1305 | 1576 | 1340 | 1325 | 1494 | 826 | 1238 | 1193 | 3034 | 2534 | 2062 | 1220 | 1899 | 2466 | 1965 | 2710 | 2865 | 1920 | 1892 |

**TEXAS RED**

| Row | 01 | 02 | 03 | 04 | 05 | 06 | 07 | 08 | 09 | 10 | 11 | 12 | 13 | 14 | 15 | 16 | 17 | 18 | 19 | 20 | 21 | 22 | 23 | 24 |
|---|---|---|---|---|---|---|---|---|---|---|---|---|---|---|---|---|---|---|---|---|---|---|---|---|
| A | 405 | 551 | 412 | 428 | 464 | 360 | 610 | 466 | 506 | 646 | 390 | 637 | 481 | 580 | 509 | 465 | 304 | 578 | 360 | 588 | 438 | 568 | 319 | 132 |
| B | 635 | 504 | 645 | 924 | 421 | 487 | 746 | 472 | 519 | 720 | 592 | 405 | 563 | 371 | 470 | 523 | 409 | 481 | 595 | 520 | 444 | 448 | 458 | 118 |
| C | 637 | 676 | 564 | 536 | 459 | 344 | 428 | 429 | 462 | 556 | 440 | 658 | 429 | 327 | 844 | 262 | 409 | 775 | 373 | 306 | 430 | 553 | 496 | 120 |
| D | 798 | 725 | 525 | 502 | 591 | 432 | 429 | 413 | 537 | 386 | 531 | 359 | 363 | 388 | 685 | 403 | 412 | 354 | 327 | 390 | 361 | 448 | 397 | 117 |
| E | 413 | 339 | 404 | 369 | 293 | 277 | 264 | 318 | 333 | 241 | 270 | 319 | 238 | 346 | 245 | 259 | 259 | 333 | 271 | 332 | 239 | 369 | 329 | 86 |
| F | 580 | 537 | 313 | 300 | 267 | 261 | 279 | 349 | 350 | 310 | 387 | 368 | 287 | 453 | 354 | 309 | 265 | 326 | 250 | 287 | 270 | 334 | 279 | 115 |
| G | 437 | 744 | 470 | 480 | 377 | 509 | 439 | 356 | 367 | 334 | 439 | 444 | 345 | 759 | 343 | 401 | 368 | 401 | 356 | 343 | 451 | 321 | 330 | 118 |
| H | 815 | 587 | 452 | 507 | 504 | 421 | 597 | 559 | 464 | 364 | 453 | 339 | 437 | 477 | 512 | 401 | 362 | 386 | 375 | 756 | 459 | 352 | 319 | 117 |
| I | 322 | 334 | 431 | 382 | 408 | 347 | 327 | 289 | 333 | 342 | 339 | 303 | 343 | 308 | 352 | 416 | 381 | 318 | 289 | 343 | 539 | 298 | 344 | 117 |
| J | 397 | 365 | 388 | 408 | 371 | 324 | 359 | 365 | 330 | 344 | 325 | 298 | 647 | 355 | 358 | 298 | 290 | 257 | 366 | 310 | 522 | 333 | 318 | 118 |
| K | 406 | 517 | 624 | 446 | 420 | 685 | 363 | 341 | 380 | 352 | 339 | 742 | 397 | 365 | 355 | 352 | 390 | 434 | 365 | 326 | 346 | 661 | 321 | 115 |
| L | 373 | 580 | 387 | 379 | 401 | 362 | 387 | 312 | 286 | 417 | 379 | 273 | 311 | 299 | 645 | 371 | 365 | 297 | 351 | 295 | 426 | 277 | 337 | 119 |
| M | 367 | 476 | 462 | 513 | 290 | 449 | 401 | 358 | 407 | 307 | 302 | 283 | 425 | 332 | 380 | 466 | 362 | 321 | 304 | 270 | 277 | 269 | 292 | 118 |
| N | 313 | 641 | 481 | 418 | 317 | 378 | 312 | 405 | 280 | 474 | 326 | 323 | 274 | 334 | 313 | 337 | 340 | 382 | 296 | 331 | 766 | 308 | 281 | 118 |
| O | 326 | 374 | 345 | 391 | 276 | 341 | 580 | 339 | 531 | 309 | 298 | 304 | 304 | 317 | 335 | 353 | 329 | 578 | 268 | 298 | 276 | 279 | 265 | 117 |
| P | 342 | 348 | 360 | 400 | 447 | 343 | 317 | 379 | 374 | 380 | 450 | 374 | 437 | 257 | 260 | 321 | 364 | 303 | 272 | 310 | 244 | 251 | 299 | 118 |

### Diverset Plate 16

**DAPI (Cell Conter)**

| Row | 01 | 02 | 03 | 04 | 05 | 06 | 07 | 08 | 09 | 10 | 11 | 12 | 13 | 14 | 15 | 16 | 17 | 18 | 19 | 20 | 21 | 22 | 23 | 24 |
|---|---|---|---|---|---|---|---|---|---|---|---|---|---|---|---|---|---|---|---|---|---|---|---|---|
| A | 406 | 3 | 1102 | 898 | 958 | 360 | 801 | 927 | 551 | 266 | 1332 | 1344 | 973 | 1250 | 1172 | 623 | 192 | 1939 | 1865 | 1697 | 1465 | 2294 | 2730 | 1399 |
| B | 905 | 457 | 165 | 274 | 257 | 909 | 378 | 294 | 662 | 942 | 948 | 1158 | 627 | 581 | 766 | 1112 | 892 | 628 | 928 | 258 | 626 | 1163 | 1085 | 533 |
| C | 1041 | 347 | 263 | 600 | 568 | 325 | 310 | 457 | 26 | 2827 | 766 | 1010 | 785 | 1144 | 919 | 1136 | 1292 | 1018 | 1251 | 590 | 899 | 339 | 1226 | 1665 |
| D | 1226 | 347 | 243 | 539 | 459 | 470 | 468 | 460 | 888 | 742 | 220 | 872 | 932 | 947 | 816 | 854 | 1186 | 126 | 1110 | 119 | 588 | 921 | 833 | 268 |
| E | 1474 | 388 | 1537 | 983 | 1932 | 3034 | 1598 | 3246 | 1792 | 2620 | 2079 | 1820 | 1674 | 2548 | 3460 | 3162 | 2809 | 3060 | 3076 | 2153 | 2114 | 2485 | 1836 | 138 |
| F | 1013 | 1724 | 940 | 1342 | 2727 | 1928 | 1894 | 1832 | 1457 | 1975 | 976 | 3243 | 2930 | 534 | 553 | 2608 | 1347 | 1604 | 881 | 1393 | 1182 | 2630 | 1033 | 1098 |
| G | 748 | 257 | 524 | 509 | 377 | 357 | 651 | 117 | 147 | 528 | 301 | 1048 | 1012 | 687 | 1074 | 705 | 944 | 1540 | 1477 | 756 | 437 | 1229 | 1409 | 188 |
| H | 633 | 145 | 374 | 215 | 630 | 627 | 559 | 616 | 669 | 607 | 776 | 1130 | 844 | 12 | 1132 | 934 | 697 | 553 | 887 | 352 | 894 | 985 | 1290 | 1394 |
| I | 655 | 599 | 842 | 1306 | 1134 | 1632 | 2181 | 2182 | 750 | 2239 | 1584 | 1753 | 319 | 2510 | 1778 | 1117 | 2045 | 2154 | 1833 | 933 | 918 | 1430 | 2860 | 3374 |
| J | 1687 | 1140 | 1037 | 1056 | 1791 | 2328 | 441 | 1143 | 1952 | 1827 | 1797 | 2045 | 1793 | 2719 | 1716 | 3064 | 1987 | 2508 | 1403 | 2414 | 1673 | 2676 | 2755 | 2971 |
| K | 1776 | 1358 | 607 | 1160 | 1524 | 1398 | 932 | 1856 | 1463 | 1531 | 1363 | 2062 | 2875 | 956 | 1733 | 1313 | 385 | 1220 | 1354 | 1895 | 2611 | 1628 | 2493 | 675 |
| L | 602 | 925 | 1118 | 1913 | 1155 | 1247 | 1600 | 1239 | 1957 | 1467 | 1485 | 2070 | 2011 | 388 | 876 | 2275 | 1725 | 2289 | 1608 | 1460 | 1961 | 2163 | 950 | 1846 |
| M | 2067 | 1120 | 1132 | 1605 | 2401 | 2244 | 1491 | 1573 | 1078 | 1368 | 1804 | 2482 | 2422 | 2349 | 1724 | 1929 | 1914 | 944 | 2709 | 2679 | 2167 | 1306 | 2699 | 2086 |
| N | 1884 | 773 | 1623 | 1413 | 1967 | 2378 | 2373 | 2399 | 2217 | 2047 | 2541 | 1948 | 2376 | 1943 | 2585 | 1819 | 1979 | 2401 | 2360 | 596 | 2192 | 2052 | 2722 | 1443 |
| O | 1899 | 1316 | 711 | 1341 | 1935 | 2556 | 1210 | 1536 | 1123 | 1686 | 2048 | 2060 | 3328 | 1807 | 2716 | 2725 | 2085 | 2267 | 2439 | 2495 | 1165 | 2132 | 2091 | 3620 |
| P | 3160 | 1802 | 1565 | 1454 | 1652 | 2772 | 2511 | 599 | 2372 | 2199 | 2497 | 2931 | 2647 | 2616 | 3158 | 2811 | 793 | 2868 | 2626 | 1345 | 1944 | 1997 | 2690 | 1948 |

**TEXAS RED**

| Row | 01 | 02 | 03 | 04 | 05 | 06 | 07 | 08 | 09 | 10 | 11 | 12 | 13 | 14 | 15 | 16 | 17 | 18 | 19 | 20 | 21 | 22 | 23 | 24 |
|---|---|---|---|---|---|---|---|---|---|---|---|---|---|---|---|---|---|---|---|---|---|---|---|---|
| A | 479 | 329 | 407 | 493 | 443 | 580 | 438 | 402 | 570 | 659 | 395 | 401 | 440 | 453 | 408 | 664 | 788 | 331 | 327 | 395 | 383 | 323 | 275 | 117 |
| B | 517 | 396 | 845 | 588 | 452 | 444 | 599 | 654 | 509 | 408 | 351 | 379 | 461 | 511 | 501 | 397 | 428 | 457 | 463 | 574 | 481 | 450 | 356 | 127 |
| C | 415 | 603 | 602 | 473 | 481 | 582 | 579 | 506 | 594 | 238 | 391 | 385 | 386 | 346 | 355 | 347 | 333 | 355 | 308 | 490 | 365 | 497 | 339 | 135 |
| D | 393 | 651 | 649 | 496 | 466 | 518 | 492 | 511 | 394 | 386 | 605 | 405 | 376 | 358 | 337 | 389 | 316 | 669 | 315 | 657 | 432 | 336 | 342 | 117 |
| E | 333 | 415 | 275 | 316 | 237 | 226 | 269 | 219 | 227 | 213 | 228 | 239 | 201 | 238 | 220 | 229 | 218 | 225 | 231 | 276 | 244 | 239 | 242 | 84 |
| F | 294 | 361 | 392 | 366 | 265 | 331 | 305 | 296 | 396 | 340 | 428 | 236 | 257 | 373 | 388 | 242 | 308 | 328 | 403 | 330 | 327 | 271 | 399 | 116 |
| G | 536 | 589 | 491 | 425 | 504 | 559 | 503 | 630 | 621 | 512 | 566 | 366 | 338 | 459 | 340 | 408 | 422 | 312 | 314 | 435 | 496 | 359 | 346 | 120 |
| H | 608 | 800 | 633 | 641 | 507 | 393 | 510 | 445 | 498 | 473 | 432 | 353 | 381 | 372 | 354 | 474 | 420 | 475 | 399 | 543 | 391 | 411 | 341 | 117 |
| I | 479 | 484 | 464 | 355 | 366 | 337 | 297 | 262 | 457 | 272 | 349 | 283 | 665 | 311 | 327 | 392 | 291 | 293 | 331 | 428 | 400 | 344 | 280 | 116 |
| J | 376 | 428 | 401 | 401 | 350 | 334 | 651 | 412 | 332 | 350 | 323 | 308 | 310 | 287 | 292 | 262 | 313 | 273 | 359 | 303 | 296 | 280 | 278 | 117 |
| K | 327 | 359 | 431 | 375 | 368 | 385 | 444 | 309 | 363 | 315 | 350 | 310 | 259 | 436 | 327 | 352 | 612 | 401 | 337 | 305 | 293 | 313 | 306 | 117 |
| L | 618 | 443 | 401 | 332 | 346 | 401 | 367 | 406 | 314 | 359 | 330 | 297 | 325 | 655 | 477 | 297 | 305 | 307 | 347 | 334 | 315 | 287 | 408 | 116 |
| M | 373 | 396 | 402 | 343 | 311 | 328 | 399 | 381 | 373 | 420 | 372 | 287 | 295 | 299 | 357 | 351 | 325 | 496 | 258 | 309 | 295 | 419 | 282 | 116 |
| N | 397 | 530 | 359 | 409 | 348 | 312 | 295 | 300 | 295 | 310 | 286 | 370 | 317 | 359 | 297 | 332 | 330 | 308 | 303 | 560 | 330 | 340 | 310 | 117 |
| O | 284 | 335 | 521 | 367 | 385 | 328 | 411 | 343 | 398 | 374 | 320 | 349 | 260 | 339 | 296 | 293 | 381 | 296 | 283 | 303 | 368 | 311 | 334 | 117 |
| P | 283 | 353 | 336 | 394 | 421 | 257 | 275 | 515 | 299 | 301 | 283 | 283 | 260 | 320 | 289 | 257 | 457 | 267 | 294 | 379 | 401 | 294 | 266 | 120 |

### Diverset Plate 17

DAPI (Cell Conter)

| Row l | 01 | 02 | 03 | 04 | 05 | 06 | 07 | 08 | 09 | 10 | 11 | 12 | 13 | 14 | 15 | 16 | 17 | 18 | 19 | 20 | 21 | 22 | 23 | 24 |
|---|---|---|---|---|---|---|---|---|---|---|---|---|---|---|---|---|---|---|---|---|---|---|---|---|
| A | 746 | 757 | 619 | 1478 | 469 | 370 | 569 | 1830 | 891 | 1066 | 1441 | 1460 | 1892 | 1822 | 1495 | 1450 | 1970 | 2654 | 1354 | 1521 | 1394 | 1441 | 1017 | 3406 |
| B | 318 | 534 | 153 | 836 | 547 | 1358 | 1156 | 815 | 806 | 1095 | 1073 | 837 | 1852 | 524 | 599 | 1369 | 939 | 878 | 1794 | 845 | 311 | 745 | 1274 | 271 |
| C | 118 | 382 | 25 | 145 | 42 | 923 | 1446 | 250 | 294 | 310 | 176 | 549 | 1189 | 621 | 396 | 709 | 417 | 193 | 1146 | 455 | 1211 | 1615 | 901 | 936 |
| D | 600 | 757 | 722 | 644 | 1106 | 415 | 1079 | 1066 | 728 | 914 | 1009 | 984 | 1370 | 1461 | 839 | 1213 | 668 | 1196 | 1222 | 1377 | 1363 | 1260 | 1020 | 722 |
| E | 25 | 707 | 274 | 1592 | 1293 | 2095 | 1556 | 837 | 1425 | 1981 | 2340 | 2222 | 1591 | 696 | 2070 | 2266 | 1605 | 1022 | 2217 | 1620 | 2215 | 1761 | 1996 | 195 |
| F | 1042 | 728 | 1228 | 882 | 1251 | 1733 | 1947 | 1843 | 1035 | 2873 | 1733 | 2879 | 2416 | 2021 | 1224 | 1884 | 2648 | 2159 | 2183 | 1502 | 1932 | 2302 | 1908 | 562 |
| G | 470 | 222 | 81 | 458 | 304 | 432 | 417 | 658 | 514 | 527 | 434 | 1312 | 1840 | 101 | 1452 | 987 | 669 | 963 | 741 | 1149 | 1360 | 1996 | 2194 | 1578 |
| H | 1246 | 438 | 479 | 356 | 317 | 622 | 322 | 736 | 787 | 647 | 1136 | 562 | 747 | 1206 | 982 | 1151 | 1148 | 1182 | 575 | 1266 | 1250 | 1348 | 1287 | 1247 |
| I | 18 | 12 | 156 | 1153 | 967 | 1217 | 1565 | 1526 | 1597 | 2417 | 890 | 2593 | 2171 | 1958 | 1930 | 2054 | 2539 | 1632 | 1733 | 2430 | 1478 | 2100 | 2918 | 1680 |
| J | 1308 | 1186 | 1284 | 1782 | 1409 | 1818 | 1282 | 1663 | 1864 | 1774 | 356 | 2229 | 2754 | 1991 | 2149 | 1866 | 2171 | 2218 | 2879 | 3254 | 3288 | 2858 | 2120 | 1357 |
| K | 1381 | 728 | 275 | 712 | 386 | 1045 | 640 | 1042 | 939 | 1246 | 779 | 1981 | 2088 | 1664 | 1556 | 1114 | 38 | 520 | 2201 | 2483 | 1739 | 1742 | 1973 | 1294 |
| L | 986 | 508 | 623 | 1069 | 881 | 1499 | 1967 | 1538 | 1672 | 2200 | 1688 | 2126 | 2511 | 2116 | 875 | 973 | 1421 | 2385 | 1958 | 2124 | 2209 | 1766 | 2145 | 1047 |
| M | 724 | 700 | 224 | 984 | 126 | 1038 | 1356 | 1539 | 1138 | 1966 | 1804 | 2454 | 2848 | 1993 | 1633 | 1990 | 2209 | 772 | 2107 | 2058 | 2569 | 1948 | 2834 | 3330 |
| N | 133 | 1672 | 1672 | 2078 | 1405 | 2394 | 2121 | 1698 | 1389 | 1611 | 2277 | 1758 | 2126 | 1859 | 1725 | 1832 | 1866 | 1793 | 1663 | 2154 | 1685 | 2062 | 2469 | 2720 |
| O | 1105 | 915 | 381 | 967 | 123 | 1303 | 1209 | 1701 | 1727 | 1275 | 2281 | 1448 | 1761 | 2754 | 1349 | 1904 | 2411 | 562 | 1489 | 2185 | 1641 | 986 | 2142 | 80 |
| P | 1456 | 1209 | 597 | 2096 | 1552 | 658 | 1787 | 1197 | 1303 | 2398 | 1403 | 632 | 1973 | 2447 | 1534 | 3623 | 1523 | 1803 | 3064 | 1382 | 1386 | 2195 | 2591 | 2947 |

Average Column La

TEXAS RED

| Row l | 01 | 02 | 03 | 04 | 05 | 06 | 07 | 08 | 09 | 10 | 11 | 12 | 13 | 14 | 15 | 16 | 17 | 18 | 19 | 20 | 21 | 22 | 23 | 24 |
|---|---|---|---|---|---|---|---|---|---|---|---|---|---|---|---|---|---|---|---|---|---|---|---|---|
| A | 320 | 466 | 393 | 272 | 458 | 494 | 411 | 326 | 455 | 352 | 391 | 360 | 365 | 320 | 331 | 418 | 339 | 271 | 276 | 406 | 351 | 416 | 470 | 117 |
| B | 798 | 443 | 702 | 463 | 539 | 363 | 370 | 419 | 497 | 348 | 344 | 492 | 310 | 511 | 441 | 366 | 431 | 424 | 298 | 412 | 651 | 445 | 399 | 115 |
| C | 717 | 518 | 899 | 746 | 748 | 425 | 321 | 624 | 495 | 567 | 663 | 487 | 381 | 473 | 552 | 431 | 475 | 629 | 345 | 483 | 341 | 339 | 353 | 118 |
| D | 452 | 483 | 446 | 439 | 350 | 564 | 352 | 358 | 444 | 413 | 384 | 367 | 324 | 278 | 382 | 345 | 429 | 348 | 323 | 305 | 301 | 329 | 369 | 119 |
| E | 427 | 372 | 438 | 283 | 316 | 260 | 274 | 331 | 312 | 270 | 241 | 245 | 288 | 383 | 251 | 258 | 272 | 288 | 238 | 251 | 265 | 262 | 289 | 57 |
| F | 427 | 365 | 362 | 345 | 367 | 321 | 317 | 307 | 328 | 250 | 311 | 241 | 298 | 303 | 315 | 290 | 282 | 281 | 248 | 324 | 304 | 271 | 304 | 115 |
| G | 486 | 693 | 929 | 475 | 588 | 546 | 501 | 444 | 426 | 516 | 509 | 327 | 302 | 782 | 329 | 388 | 453 | 403 | 432 | 356 | 350 | 302 | 279 | 118 |
| H | 332 | 496 | 466 | 637 | 693 | 496 | 511 | 464 | 433 | 531 | 392 | 448 | 489 | 352 | 390 | 363 | 353 | 368 | 500 | 368 | 310 | 327 | 368 | 116 |
| I | 263 | 370 | 676 | 325 | 402 | 403 | 302 | 348 | 350 | 311 | 422 | 265 | 276 | 280 | 302 | 275 | 276 | 317 | 313 | 278 | 330 | 320 | 245 | 116 |
| J | 506 | 364 | 408 | 315 | 355 | 329 | 388 | 354 | 296 | 322 | 700 | 252 | 265 | 303 | 297 | 293 | 270 | 281 | 229 | 246 | 237 | 279 | 314 | 116 |
| K | 409 | 432 | 666 | 421 | 565 | 421 | 458 | 361 | 397 | 356 | 437 | 306 | 280 | 348 | 301 | 342 | 492 | 513 | 298 | 277 | 304 | 304 | 298 | 117 |
| L | 462 | 574 | 554 | 424 | 375 | 325 | 311 | 310 | 324 | 289 | 339 | 293 | 293 | 313 | 446 | 411 | 326 | 256 | 280 | 299 | 280 | 311 | 314 | 116 |
| M | 527 | 477 | 558 | 390 | 846 | 450 | 400 | 351 | 399 | 334 | 298 | 297 | 300 | 335 | 336 | 308 | 348 | 489 | 320 | 308 | 283 | 331 | 285 | 116 |
| N | 760 | 385 | 361 | 332 | 416 | 312 | 338 | 338 | 405 | 381 | 310 | 346 | 294 | 365 | 334 | 322 | 317 | 323 | 326 | 324 | 370 | 361 | 305 | 116 |
| O | 369 | 454 | 552 | 473 | 702 | 375 | 366 | 304 | 290 | 339 | 266 | 334 | 328 | 300 | 338 | 289 | 315 | 424 | 334 | 290 | 333 | 445 | 283 | 117 |
| P | 381 | 346 | 438 | 301 | 325 | 563 | 340 | 413 | 355 | 299 | 334 | 474 | 305 | 267 | 280 | 227 | 348 | 316 | 246 | 390 | 338 | 244 | 255 | 119 |

### Diverset Plate 18

DAPI (Cell Conter)

| Row | 01 | 02 | 03 | 04 | 05 | 06 | 07 | 08 | 09 | 10 | 11 | 12 | 13 | 14 | 15 | 16 | 17 | 18 | 19 | 20 | 21 | 22 | 23 | 24 |
|---|---|---|---|---|---|---|---|---|---|---|---|---|---|---|---|---|---|---|---|---|---|---|---|---|
| A | 546 | 338 | 150 | 191 | 733 | 1209 | 431 | 637 | 1175 | 1159 | 1631 | 855 | 1730 | 1150 | 2247 | 1063 | 1257 | 1641 | 678 | 1126 | 218 | 1281 | 1985 | 2100 |
| B | 209 | 10 | 155 | 363 | 183 | 241 | 800 | 253 | 550 | 300 | 461 | 218 | 782 | 482 | 799 | 671 | 629 | 349 | 840 | 550 | 1074 | 368 | 609 | 373 |
| C | 698 | 258 | 162 | 245 | 605 | 606 | 781 | 224 | 102 | 802 | 1184 | 255 | 814 | 641 | 1022 | 541 | 882 | 1261 | 808 | 1127 | 1458 | 1263 | 915 | 174 |
| D | 415 | 488 | 375 | 580 | 505 | 269 | 446 | 393 | 590 | 553 | 54 | 795 | 1033 | 737 | 500 | 962 | 821 | 849 | 1201 | 61 | 683 | 1205 | 813 | 335 |
| E | 308 | 519 | 690 | 1096 | 2286 | 717 | 815 | 1813 | 2078 | 1544 | 1170 | 2289 | 1762 | 1828 | 2005 | 1639 | 2279 | 1642 | 1326 | 2198 | 2286 | 1483 | 1281 | 22 |
| F | 194 | 808 | 838 | 529 | 839 | 1562 | 727 | 1293 | 844 | 936 | 1925 | 1522 | 1336 | 2324 | 1435 | 1258 | 1808 | 141 | 2462 | 1653 | 2147 | 2208 | 1190 | 189 |
| G | 954 | 147 | 415 | 471 | 786 | 624 | 268 | 242 | 890 | 384 | 404 | 521 | 1163 | 348 | 760 | 1704 | 947 | 921 | 1774 | 1265 | 1436 | 735 | 1694 | 59 |
| H | 672 | 137 | 277 | 340 | 563 | 92 | 387 | 796 | 48 | 645 | 847 | 288 | 1078 | 732 | 549 | 1006 | 878 | 999 | 992 | 1346 | 600 | 1364 | 1099 | 1645 |
| I | 1261 | 414 | 806 | 1164 | 658 | 1270 | 1220 | 1545 | 1643 | 1275 | 1222 | 1987 | 1719 | 1647 | 1140 | 1892 | 1763 | 1072 | 1674 | 1749 | 2575 | 705 | 2199 | 1229 |
| J | 1097 | 335 | 1149 | 918 | 1104 | 1342 | 1266 | 1577 | 657 | 1514 | 1747 | 1180 | 593 | 1902 | 1616 | 1932 | 2487 | 1353 | 2018 | 1765 | 1452 | 1522 | 1852 | 870 |
| K | 704 | 219 | 987 | 725 | 798 | 899 | 1158 | 1840 | 1667 | 1626 | 2159 | 2045 | 1552 | 1380 | 1184 | 601 | 1879 | 1538 | 1604 | 1645 | 1093 | 2689 | 1731 | 1607 |
| L | 1853 | 391 | 719 | 400 | 1447 | 1121 | 112 | 1821 | 1476 | 1445 | 2255 | 1613 | 1561 | 1268 | 1814 | 1399 | 1784 | 699 | 1620 | 1487 | 1757 | 1348 | 1500 | 638 |
| M | 1048 | 352 | 765 | 1394 | 1350 | 1690 | 1973 | 1616 | 1561 | 1459 | 2057 | 1633 | 2536 | 2057 | 2051 | 1065 | 1952 | 2105 | 2329 | 2215 | 2367 | 222 | 1476 | 1511 |
| N | 765 | 321 | 735 | 1130 | 1156 | 1701 | 1917 | 23 | 755 | 1823 | 2933 | 1061 | 2560 | 1954 | 1976 | 577 | 1591 | 156 | 2150 | 827 | 874 | 474 | 1415 | 178 |
| O | 1727 | 2109 | 1214 | 1203 | 2406 | 2288 | 1850 | 2252 | 1626 | 2708 | 1936 | 2500 | 652 | 2227 | 2409 | 1590 | 2281 | 2425 | 1921 | 2605 | 2558 | 2494 | 1743 | 2211 |
| P | 1698 | 2120 | 2177 | 828 | 2352 | 1294 | 1338 | 2145 | 2580 | 1853 | 1722 | 1506 | 1841 | 1316 | 2661 | 2340 | 2640 | 3153 | 2571 | 2881 | 3124 | 2691 | 2495 | 1308 |

TEXAS RED

| Row | 01 | 02 | 03 | 04 | 05 | 06 | 07 | 08 | 09 | 10 | 11 | 12 | 13 | 14 | 15 | 16 | 17 | 18 | 19 | 20 | 21 | 22 | 23 | 24 |
|---|---|---|---|---|---|---|---|---|---|---|---|---|---|---|---|---|---|---|---|---|---|---|---|---|
| A | 540 | 595 | 766 | 613 | 561 | 397 | 557 | 489 | 346 | 327 | 354 | 422 | 369 | 390 | 311 | 386 | 345 | 285 | 491 | 394 | 678 | 314 | 328 | 119 |
| B | 524 | 325 | 750 | 563 | 641 | 759 | 434 | 657 | 526 | 656 | 488 | 679 | 502 | 544 | 500 | 472 | 503 | 427 | 444 | 563 | 410 | 443 | 437 | 118 |
| C | 494 | 722 | 741 | 619 | 465 | 513 | 439 | 631 | 738 | 395 | 367 | 663 | 415 | 403 | 351 | 489 | 399 | 325 | 382 | 375 | 373 | 337 | 337 | 174 |
| D | 613 | 590 | 596 | 549 | 484 | 683 | 520 | 647 | 507 | 471 | 793 | 350 | 372 | 405 | 570 | 389 | 440 | 373 | 335 | 776 | 420 | 348 | 386 | 116 |
| E | 520 | 387 | 398 | 352 | 289 | 390 | 387 | 303 | 260 | 285 | 312 | 265 | 295 | 270 | 272 | 289 | 291 | 290 | 341 | 277 | 259 | 319 | 319 | 58 |
| F | 699 | 413 | 409 | 455 | 407 | 343 | 466 | 397 | 427 | 384 | 305 | 250 | 353 | 250 | 276 | 303 | 289 | 576 | 288 | 351 | 309 | 292 | 389 | 117 |
| G | 501 | 639 | 572 | 426 | 398 | 451 | 619 | 626 | 409 | 589 | 522 | 549 | 349 | 457 | 478 | 327 | 349 | 394 | 315 | 399 | 336 | 445 | 323 | 117 |
| H | 483 | 532 | 665 | 604 | 523 | 828 | 618 | 446 | 742 | 496 | 440 | 689 | 415 | 511 | 533 | 474 | 386 | 384 | 400 | 360 | 458 | 351 | 374 | 118 |
| I | 420 | 662 | 419 | 362 | 477 | 423 | 364 | 363 | 328 | 397 | 371 | 317 | 331 | 339 | 439 | 305 | 322 | 432 | 341 | 352 | 276 | 525 | 307 | 130 |
| J | 356 | 616 | 434 | 396 | 463 | 380 | 346 | 347 | 507 | 372 | 328 | 399 | 538 | 334 | 371 | 327 | 304 | 402 | 346 | 345 | 359 | 380 | 331 | 123 |
| K | 545 | 669 | 414 | 535 | 431 | 462 | 439 | 313 | 363 | 341 | 289 | 311 | 387 | 380 | 378 | 546 | 323 | 367 | 316 | 326 | 407 | 262 | 352 | 117 |
| L | 355 | 593 | 608 | 538 | 393 | 385 | 809 | 384 | 371 | 382 | 313 | 391 | 365 | 412 | 329 | 411 | 326 | 532 | 347 | 346 | 372 | 382 | 341 | 118 |
| M | 468 | 628 | 502 | 406 | 418 | 352 | 316 | 397 | 401 | 376 | 343 | 356 | 326 | 324 | 366 | 434 | 359 | 316 | 315 | 318 | 322 | 664 | 363 | 117 |
| N | 527 | 791 | 513 | 423 | 459 | 393 | 404 | 862 | 537 | 362 | 307 | 451 | 323 | 316 | 345 | 589 | 376 | 828 | 337 | 519 | 421 | 540 | 376 | 122 |
| O | 324 | 385 | 416 | 387 | 324 | 327 | 360 | 338 | 307 | 281 | 353 | 313 | 534 | 350 | 305 | 369 | 340 | 326 | 319 | 290 | 287 | 289 | 377 | 119 |
| P | 374 | 313 | 360 | 553 | 339 | 451 | 446 | 298 | 281 | 349 | 347 | 396 | 356 | 354 | 301 | 304 | 291 | 266 | 280 | 273 | 275 | 299 | 211 | 124 |

**Supplementary Figure1:** High Throughput Screening assay plate results, with the number of cells per well and quantification of Texas red intensity for the several tested compounds.

### Supplementary Figure 2

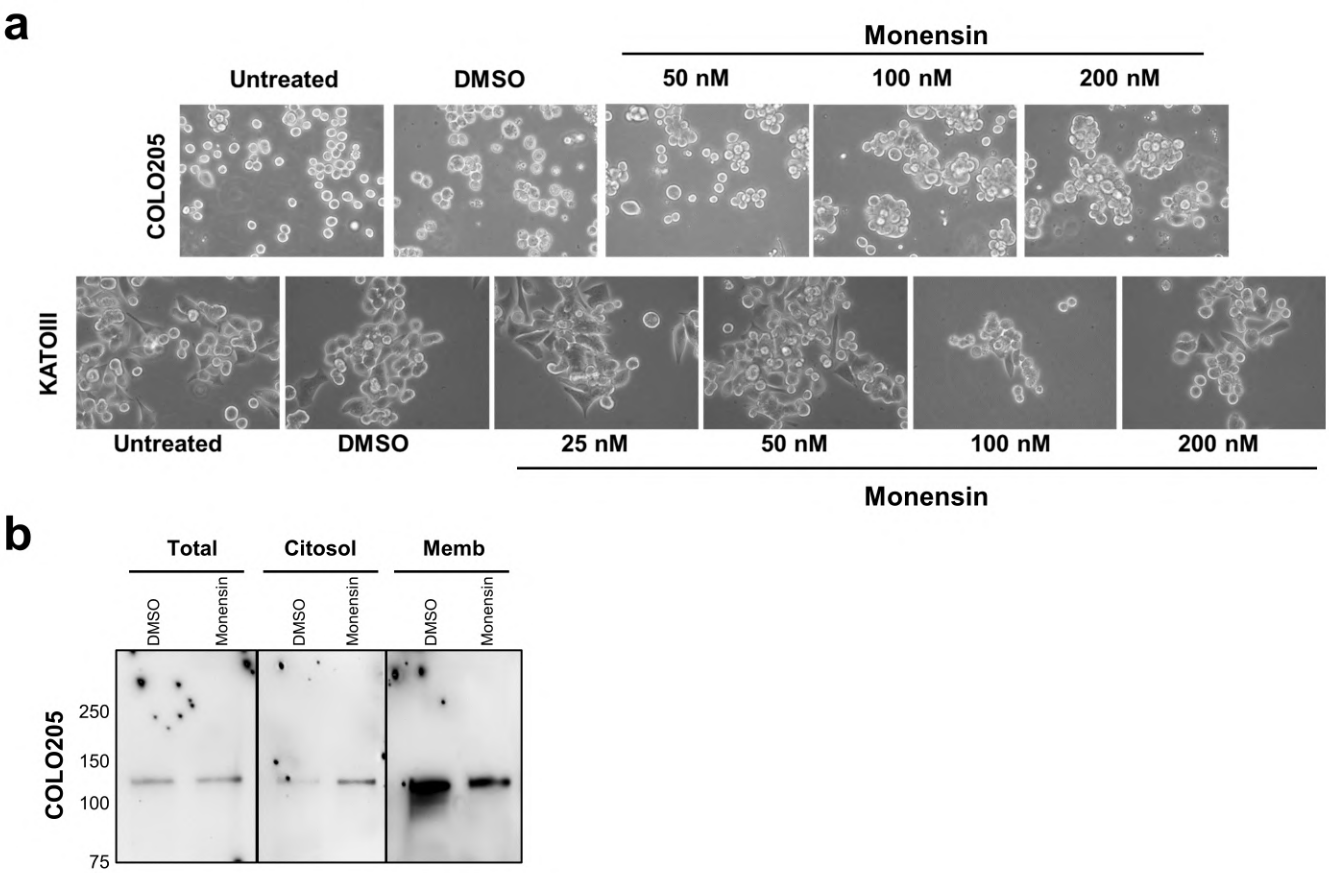


**Supplementary Figure 2: Effect of monensin treatment on CRC and GC cell morphology and in E-Cadherin expression in CRC cells. (a)** Brightfield microscopy images upon 72h of monensin treatment showing morphological alteration in both COLO205 and KATOIII treated cells compared to the untreated or DMSO treated cells. It is observed a larger vacuolar cell phenotype and an increase cells' aggregation upon Monensin treatment (Magnification 200x). **(b)** Western blot analysis of E-cadherin expression at different cellular compartments (total, cytosolic and membrane) in COLO205 cells. Monesisn treatment led to a reduction of E-cadherin expression at membrane fraction and an increase at the cytosolic fraction.

### Supplementary Figure 3

**a**

**KATOIII**
**DMSO**

**H&E**

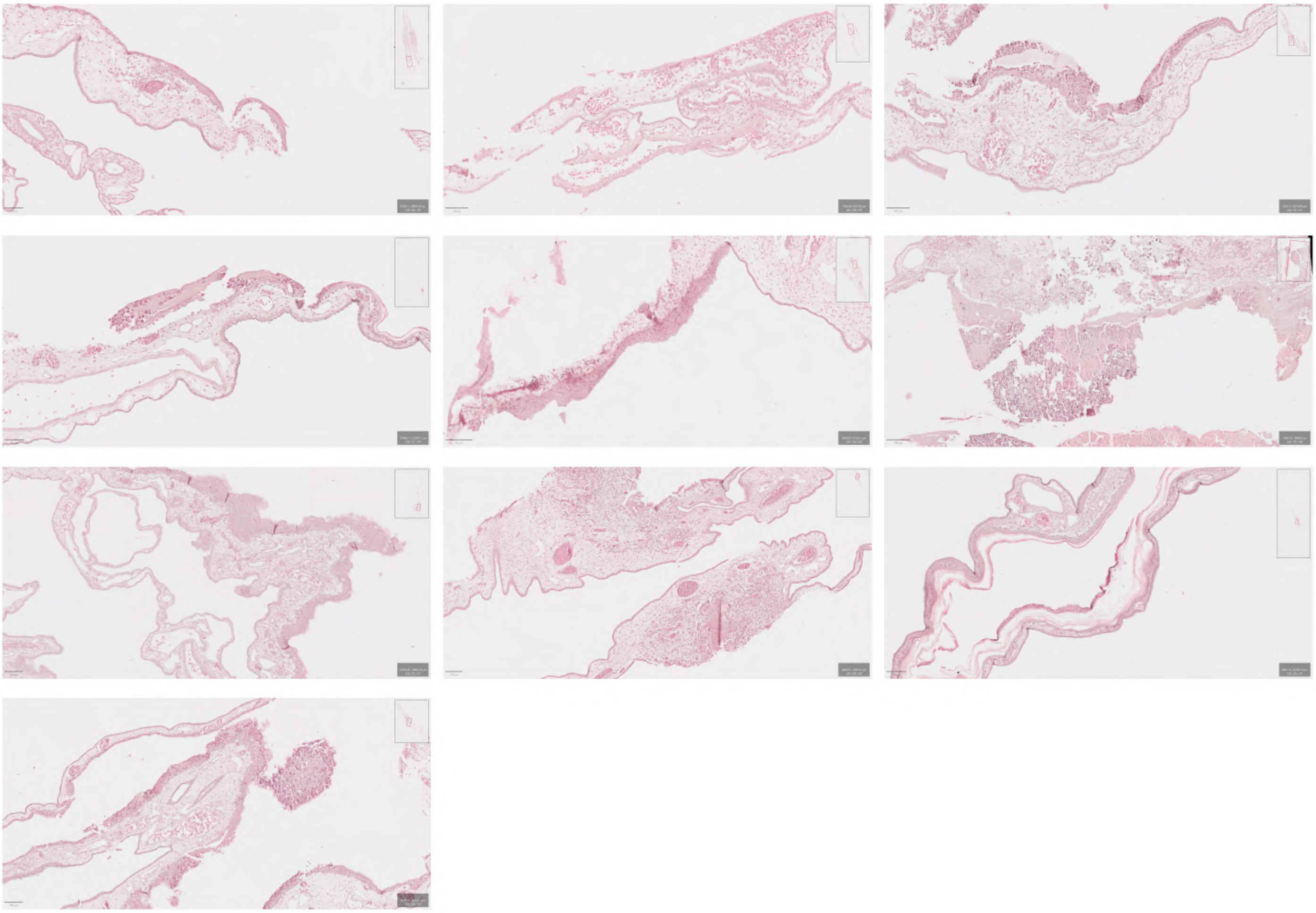

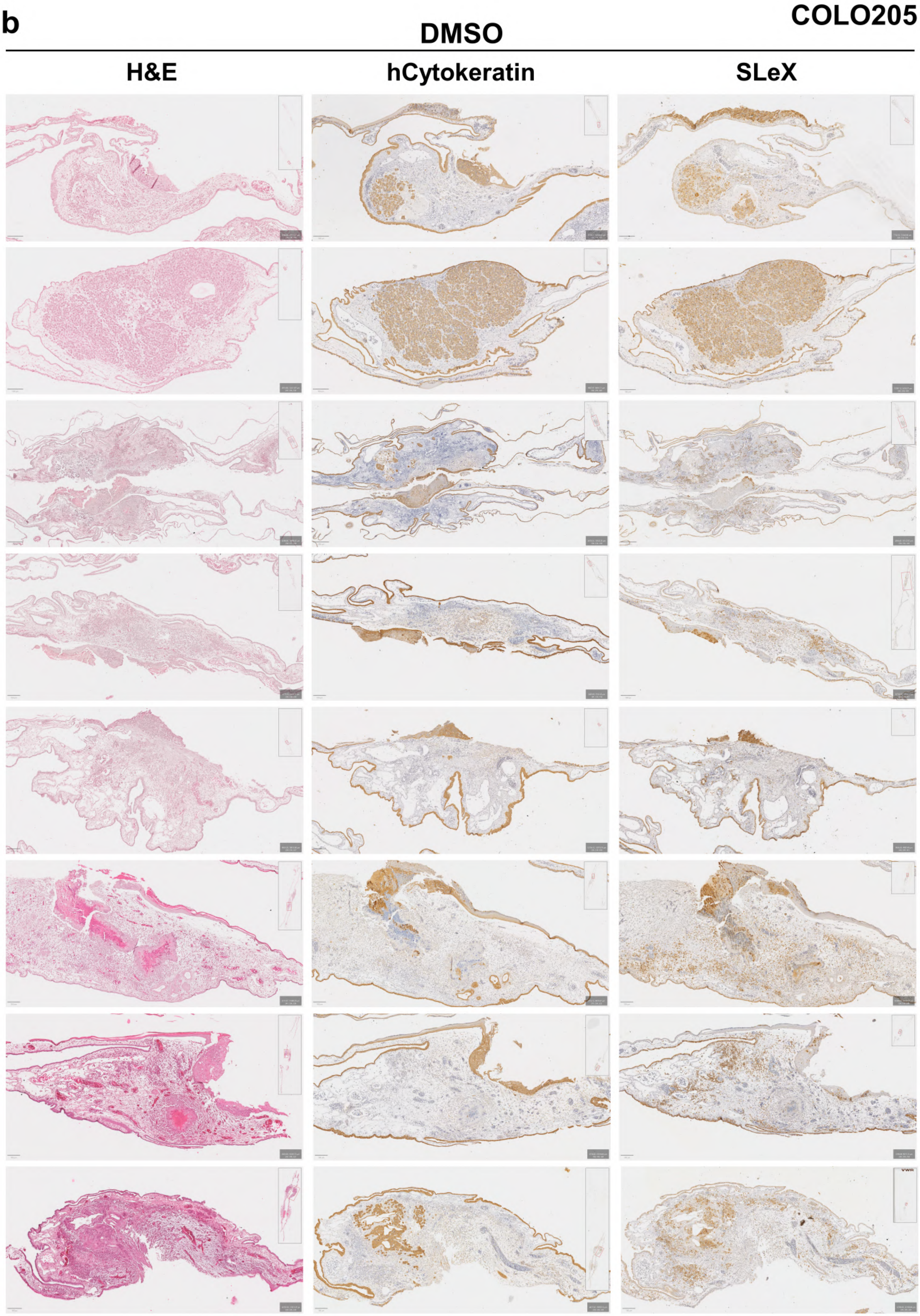
b
COLO205
DMSO
H&E
hCytokeratin
SLeX

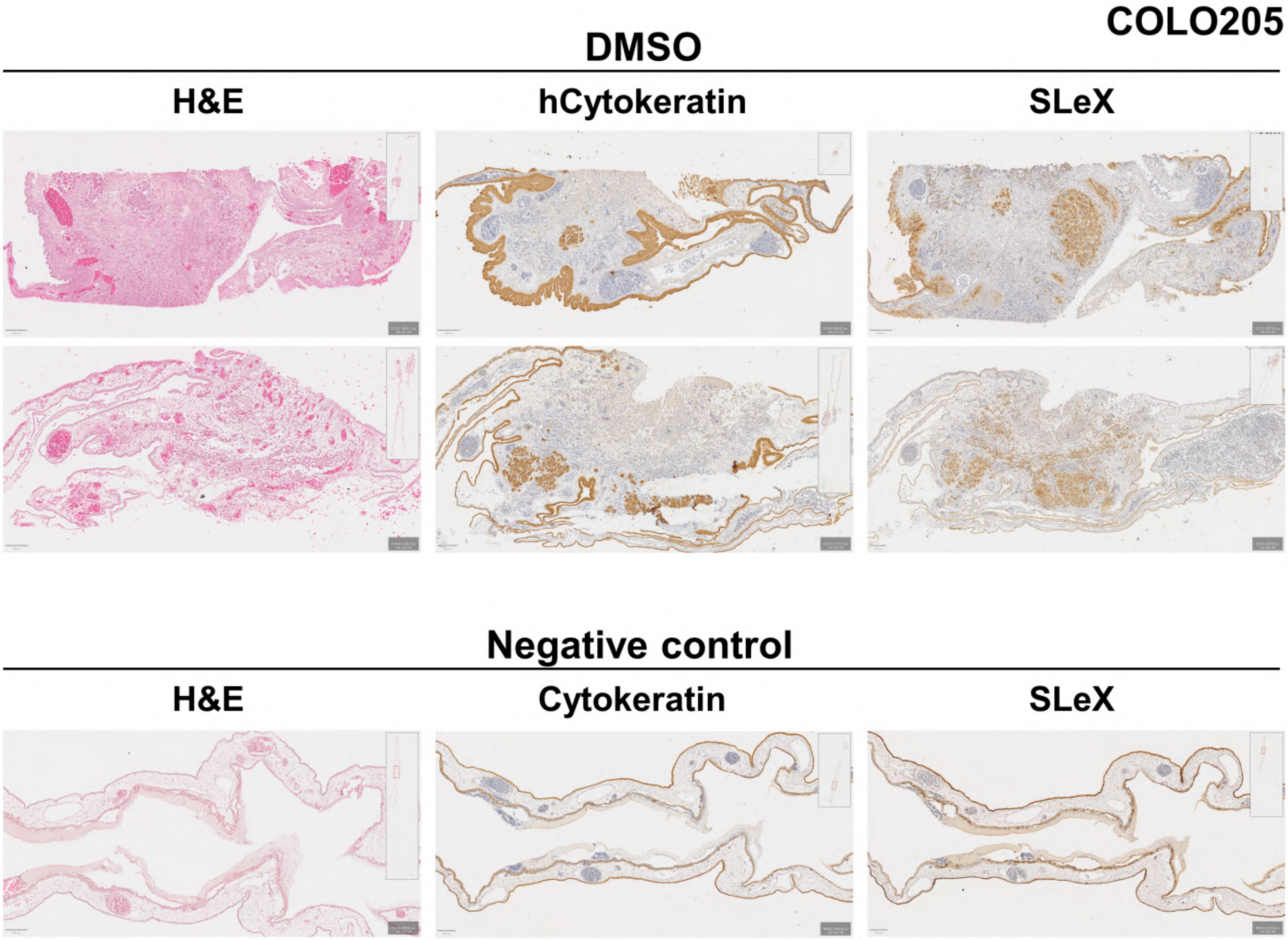
COLO205
DMSO
H&E
hCytokeratin
SLeX
Negative control
H&E
Cytokeratin
SLeX

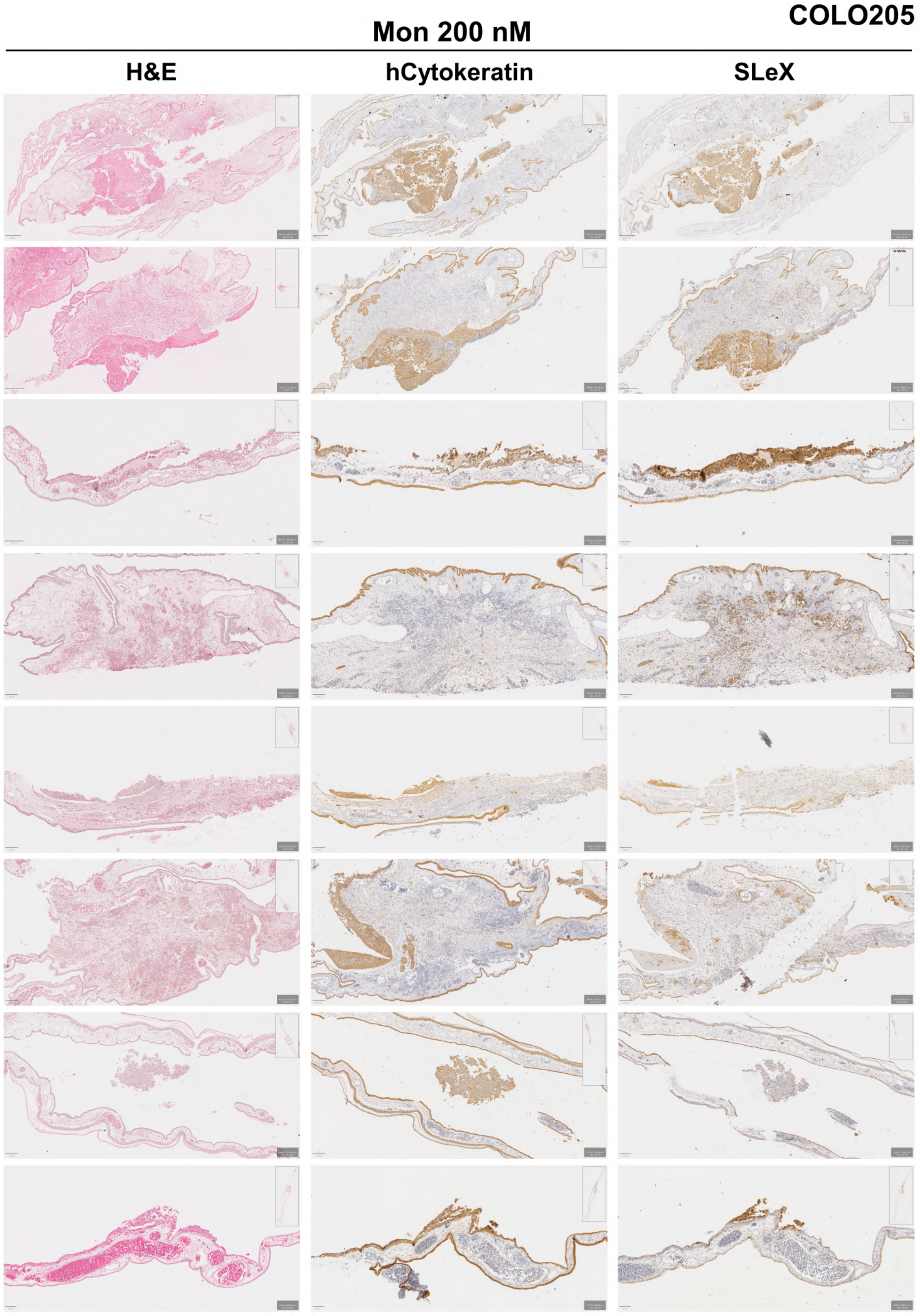
COLO205
Mon 200 nM
H&E
hCytokeratin
SLeX

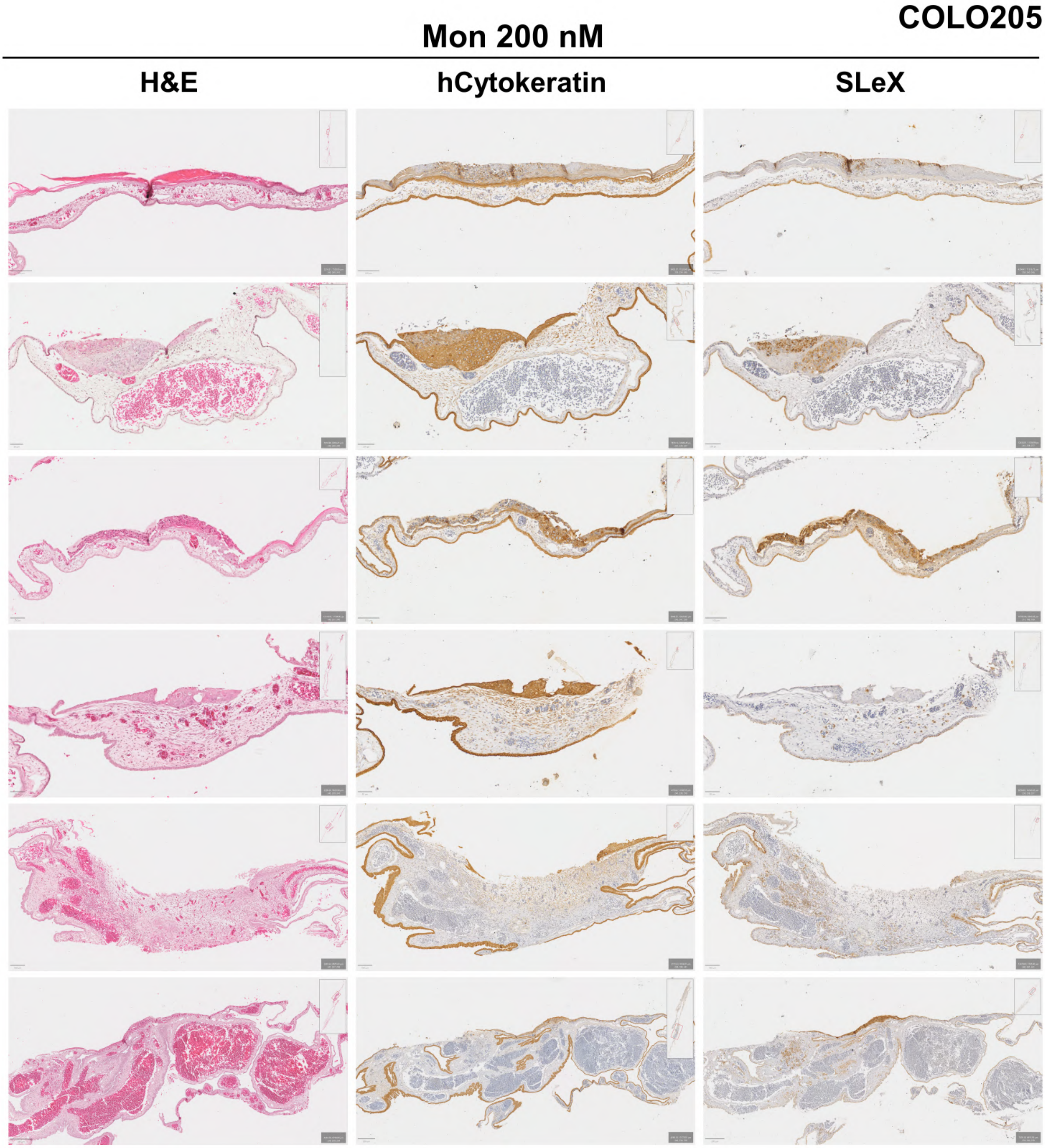


**Supplementary Figure 3: Histological analysis of chick chorioallantoic membrane (CAM) xenografted with KATOIII and COLO205 cell lines. (a)** Hematoxilin & Eosin (H&E) staining of KATOIII xenografted CAMs at day 6 showing no tumor formation. **(b)** Histological characterization (H&E, human cytokeratins and SLeX) of COLO205 xenografted CAMs showing tumor formation at day 6 in DMSO-treated, monensin-treated and Vitrogel control CAMs. Results show differences in tumor size and cell invasion throughout chicken membrane in monesin treated CAMs. Scale bars corresponds to 50-200 µm.

### Supplementary Figure 4

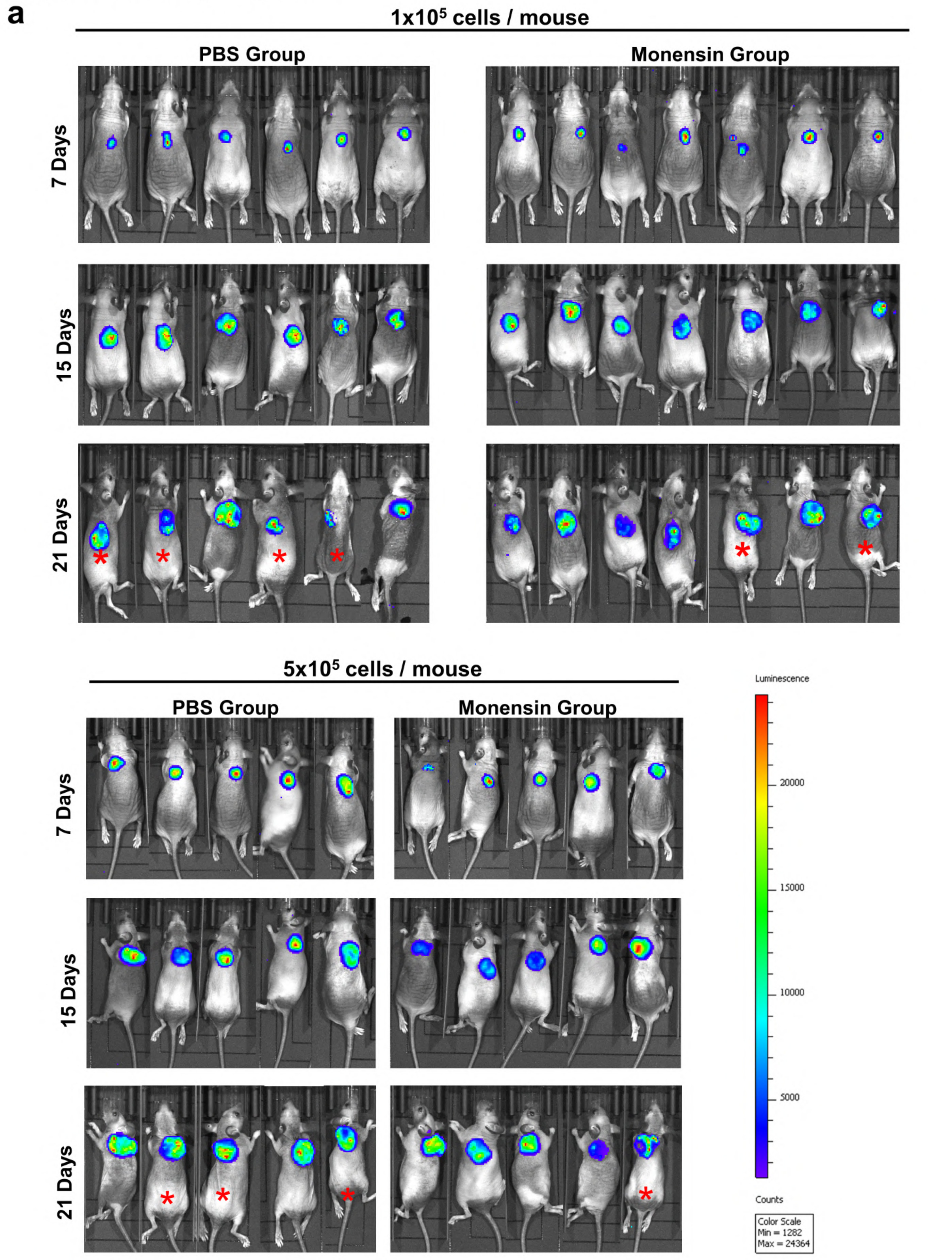


* The animal's tumor ulcerated shortly before the end of the experiment (1-2 days before), which led us, in some cases, to use the last measurement before the ulcer occurs.

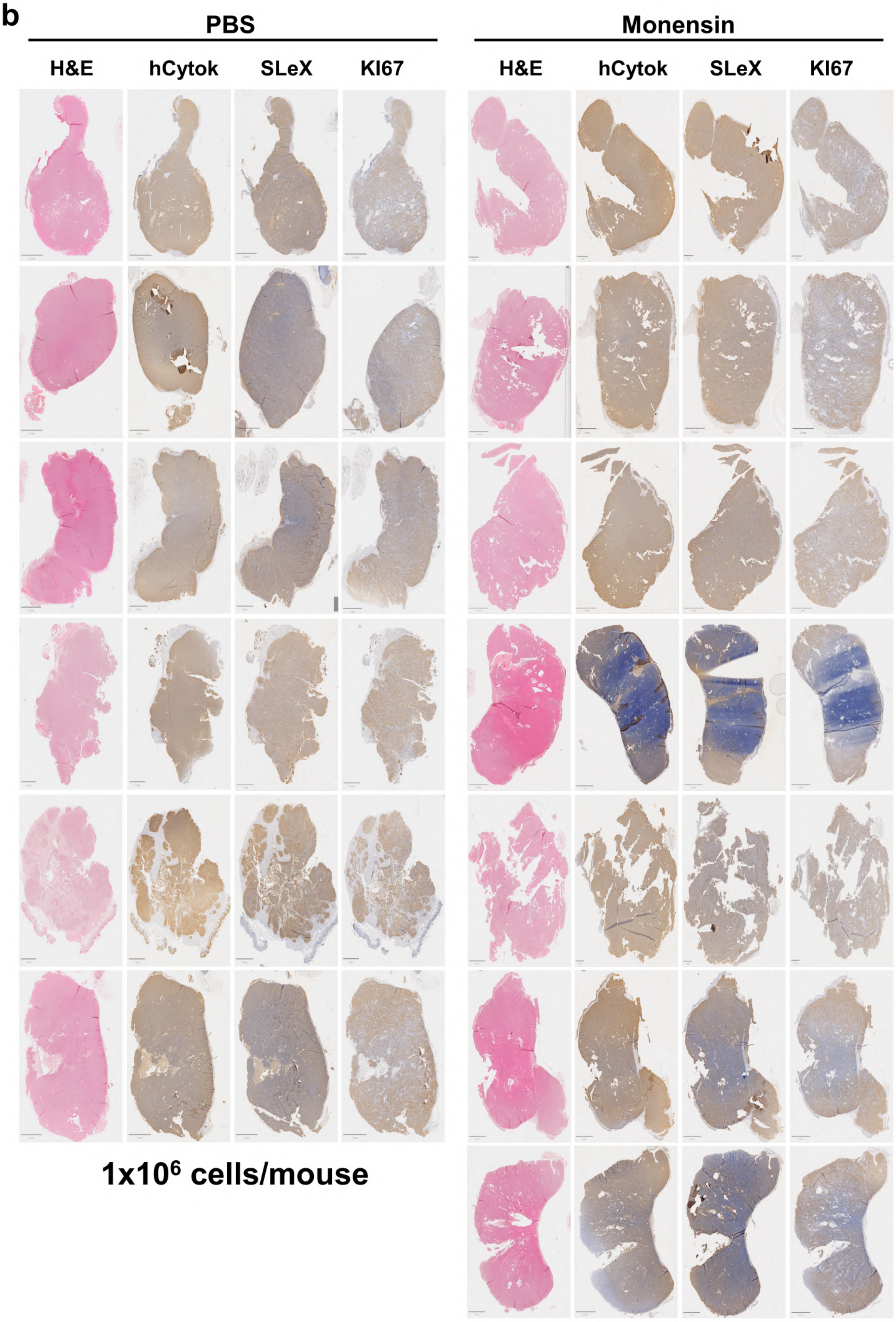
b
PBS
Monensin
H&E
hCytok
SLeX
KI67
H&E
hCytok
SLeX
KI67
1x10^6 cells/mouse

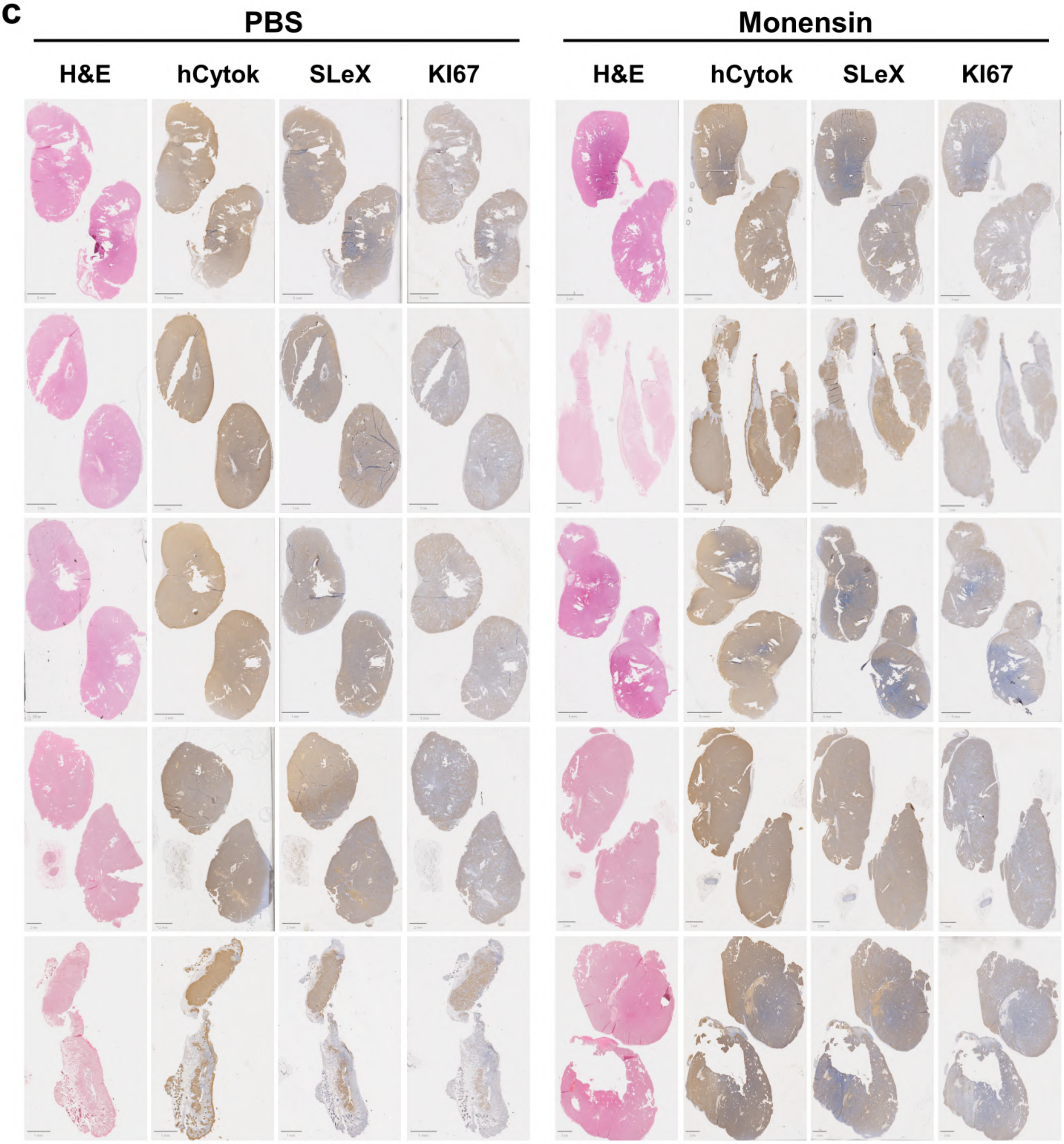


5x10$^6$ cells/mouse

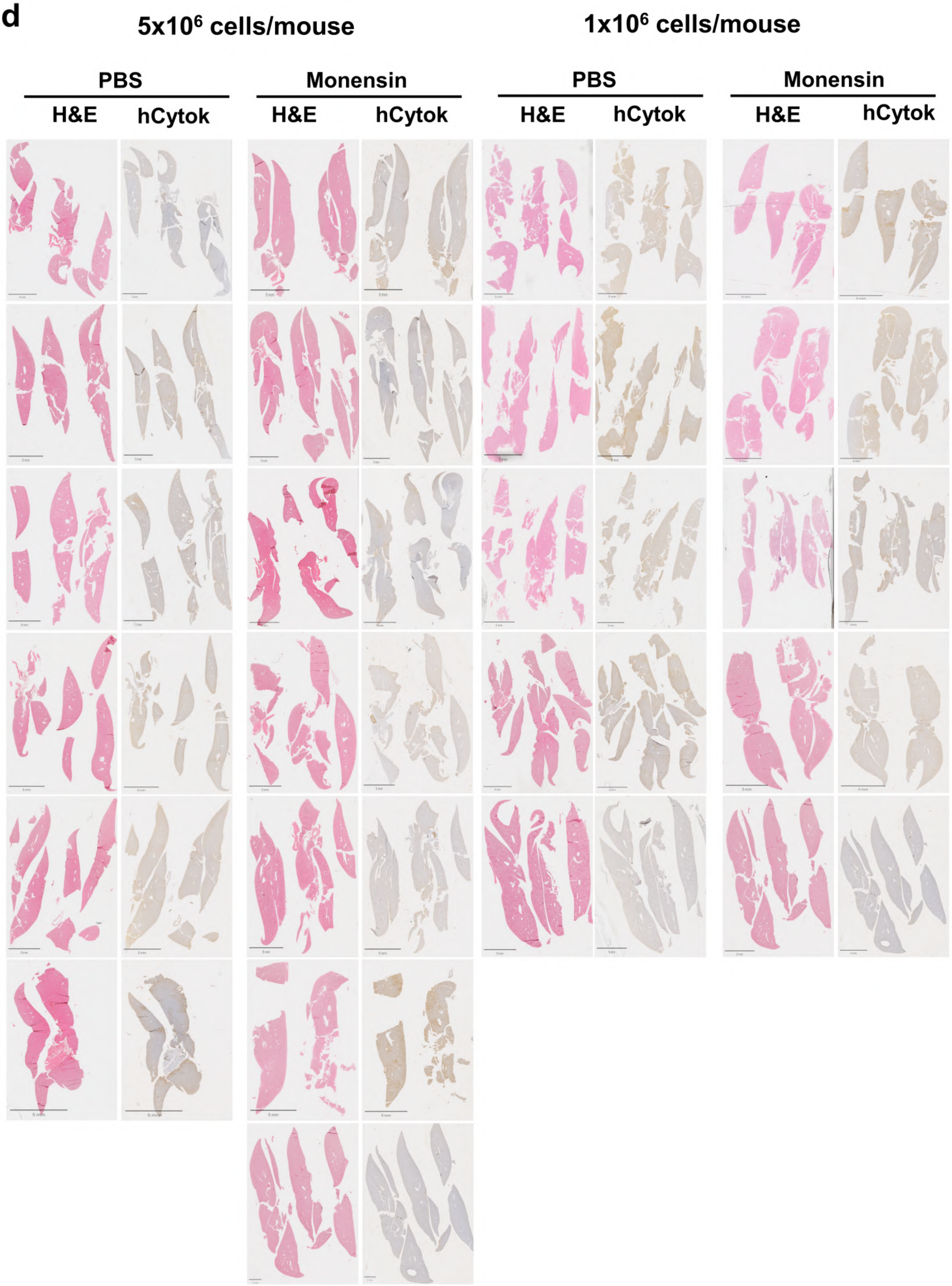

d
5x10⁶ cells/mouse
1x10⁶ cells/mouse
PBS
Monensin
PBS
Monensin
H&E
hCytok
H&E
hCytok
H&E
hCytok
H&E
hCytok

**Supplementary Figure 4: *In vivo* biological effect of monensin in Luciferase-positive COLO205 xenograft mouse model using 1 million and 5 million cell inoculum.** **(a)** *In vivo* tumor growth by measuring luciferase activity throughout time. **(b)** Tumor images of Hematoxilin & Eosin (H&E) staining and immunodetection of human cytokeratin, SLeX and KI67 in mice subcutaneously injected with 1 million cells. **(c)** Tumor images of Hematoxilin & Eosin (H&E) staining and immunodetection of human cytokeratin, SLeX and KI67 in mice subcutaneously injected with 5 million cells. **(d)** H&E and human cytokeratin staining of livers from mice injected with 1 or 5 million cells showing no metastatic lesions.

### Supplementary Figure 5

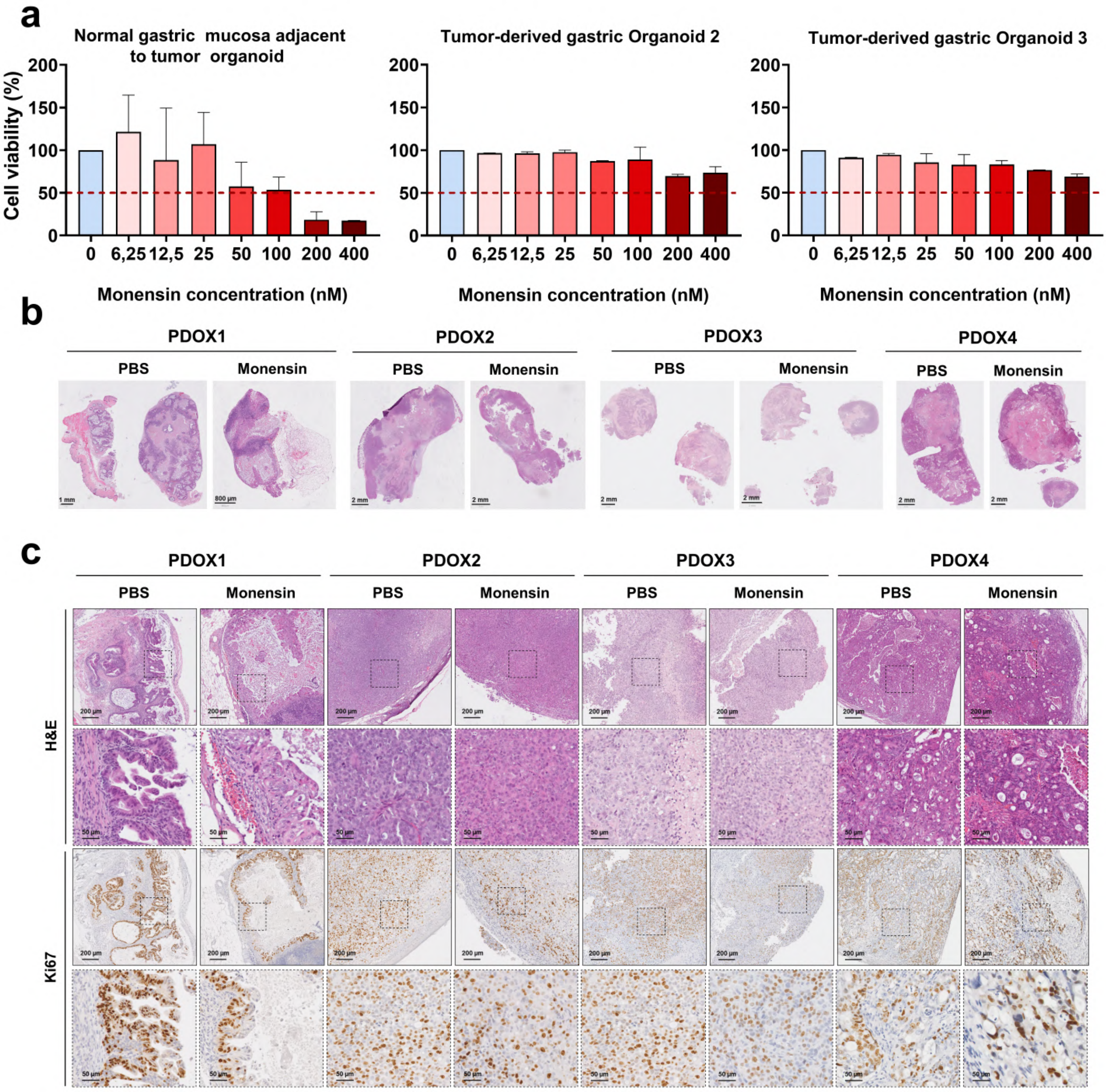


**Supplementary Figure 5: Monensin biological effect in a pre-clinical organoid model. (a)** Viability assay of healthy (left) and two tumor-patient (rigth) derived gastric organoids after monensin treatment. **(b)** Tumor´s images (H&E staining) of PDOX DMSO and Monensin-treated **(c)** Detailed histological characterization, H&E and Ki67 immunodetection, of PDOX tumors.
